# Proteomic Profiles of Obesity-Related Phenotypes and Incident Cardiovascular Events

**DOI:** 10.1101/2025.02.27.25323042

**Authors:** Chang Liu, Bojung Seo, Qin Hui, Peter WF Wilson, Arshed A Quyyumi, Yan V Sun

## Abstract

**Background:** Obesity is a modifiable cardiovascular risk factor. Proteomic profiling may improve the understanding of obesity and cardiovascular risk prediction. This study explores the use of protein-predicted scores for body mass index (PPS_BMI_), body fat percentage (PPS_BFP_), and waist-hip ratio (PPS_WHR_) in predicting major adverse cardiovascular events (MACE).

**Methods:** We used data from the UK Biobank with proteome profiling. PPS_BMI_, PPS_BFP_, and PPS_WHR_ were derived using LASSO algorithm. The association between these protein scores and incident MACE was evaluated using competing risk model. MACE prediction using the protein scores was compared with the PREVENT equation.

**Results:** Strong correlations were observed between protein-predicted obesity phenotypes and their measured counterparts (R^2^: BMI = 0.78, BFP = 0.85, WHR = 0.63). Per standard deviation of elevated PPS_BFP_ and PPS_WHR_, but not PPS_BMI_, was associated with increased risk of MACE (Hazard Ratio [HR] 1.25, 95% CI 1.14 - 1.38; HR 1.15, 95% CI 1.06 - 1.24, respectively), independent of traditional risk factors and outperformed the measured obesity-related phenotypes. For predicting MACE, compared with the PREVENT equation (C-statistic 0.694), the models adjusted for only age, sex, current smoking, and protein scores of obesity showed comparable performance (C-statistics 0.685 - 0.688).

**Conclusion:** Protein-predicted scores are better predictors of MACE than measured obesity phenotypes. Integration of protein-predicted scores provides a biologically relevant risk assessment using a single biochemical assay to potentially simplify and standardize the measure of obesity risk in clinical practice.

## INTRODUCTION

Over the past two decades, the prevalence of obesity has grown continuously, and the age-adjusted prevalence of obesity among U.S. adults has reached 42.4%.^1^ Obesity is a well-established modifiable risk factor for adverse cardiovascular events.^2^ While Body Mass Index (BMI) has been used as a simple and standard metric for measuring obesity, its limitations in capturing body composition nuances have prompted a search for more comprehensive measures of obesity risk.^3^ This has led to the emergence of body fat percentage (BFP) and waist-hip ratio (WHR), which can offer a more refined understanding of adiposity and its impact on health.^4,5^

Proteomics is an emerging field that holds promise in unraveling the complex molecular mechanisms underlying various phenotypes. Advancements in high-throughput technology of proteomics have enabled a comprehensive exploration of the molecular landscape underlying obesity.^6^ Using proteomic markers to understand obesity-related phenotypes and their potential role in the prediction of cardiovascular events remain relatively unexplored. Recent studies have shown that protein risk scores that incorporate proteomic profiles can enhance prediction of adverse cardiovascular events in both primary and secondary event populations, outperforming traditional risk factors.^7,8^ Leveraging obesity-associated proteomics improves our understanding of obesity at the molecular level, and thus may enable more precise estimates of adverse cardiovascular outcomes beyond directly measured phenotypes. In our study, we aimed to identify proteomic markers and scores associated with obesity-related phenotypes, such as BMI, BFP, and WHR, and assess the associations and predictive performance of these proteomic scores for incident major adverse cardiovascular events (MACE).

## METHODS

### UK Biobank Cohort

The UK Biobank (UKB) is a large-scale biomedical database, with the study design and cohort profile described previously.^9^ Established in 2006, the UKB recruited approximately half a million participants aged between 40 and 69 years old from United Kingdom. Participants completed standard questionnaires and provided detailed information about medical conditions, lifestyle, environment, physical measurements and biological measures. The UKB cohort was linked to Hospital Episode Statistics data for hospital admissions, primary care data, and a death registry included date of death, and both primary and secondary causes of death. All first occurrences of disease and cause of death were mapped to International Classification of Diseases, Tenth Revision (ICD-10) codes.^9^ Proteomic profiling on blood plasma samples was performed for 54,219 participants in the UKB using the antibody based Olink Explore 3072 PEA platform. A total of 2,923 distinct proteins were measured.^10^ The UKB study was approved by the North West Multi-center Research Ethics Committee, and consent was obtained from all participants.

Among the participants with proteomic profiling, a sub-cohort of 18,664 participants formed the healthy cohort without prevalent or incident diabetes (ICD-10 codes E10-E14), cardiovascular disease (ICD-10 codes I00-I13, I15, I20-I51, I60-I69), renal disease (ICD-10 codes N17-N23, N25-N29) or cancer. Additionally, a total of 20,683 participants without prevalent stroke (ICD-10 codes I60, I61, I63, I64) and coronary artery disease (ICD-10 codes I20-I25) were included for prediction of MACE, a composite event that included incident ischemic stroke (algorithmically-defined, Data-Field 42008), myocardial infarction (MI, ICD-10 codes I21-I23, I25) and cardiovascular death (ICD-10 codes I00-I13, I15, I20-I51, I60-I69).

At baseline, demographics and risk factors were collected at enrollment, including age, sex, race and ethnicity, total cholesterol levels, high-density lipoprotein cholesterol (HDL-C) levels, systolic blood pressure, estimated glomerular filtration rate (eGFR) calculated using the 2021 CKD-EPI equation,^11^ diabetes, smoking status, blood pressure lowering medication use, and cholesterol lowering medication use. Baseline obesity-related phenotypes including BMI, BFP, and WHR were obtained. BFP (%) was defined as the total mass of fat divided by total body mass, multiplied by 100. WHR was defined as the ratio of waist circumference to hip circumference. Time to MACE and the 3 subcomponents was defined as the duration from enrollment until the event, loss to follow-up, or the conclusion of follow-up in September 2023.

### Statistical Analysis

After excluding participants with > 20% missing rate across the 2,923 proteins, a total of 15,652 participants in the healthy cohort and additional 24,999 participants without prevalent stroke and coronary artery disease were included in the analysis. Four proteins with > 20% missing rate across all samples were excluded, and 2,919 proteins remained in the analysis. The missingness of the protein levels were imputed to the minimum value across the samples, assuming they were below the detectable limit. The relative protein abundance was normalized using rank-based inverse normalization.

Cohort characteristics were compared between the cohorts with and without MACE using a two-sample t-test for continuous variables, and Chi-squared test for categorical variables.

The protein scores for obesity-related phenotypes, including BMI, BFP, and WHR, were trained using data from the healthy cohort. To determine the adequate sample size for the training set of these protein scores, we partitioned the healthy cohort randomly into increments of 10%, ranging from 10% to 90%. Each partition was repeated 100 times, with participants randomly selected from the healthy cohort, and the remaining participants constituted the test set. Within the training set, we applied the least absolute shrinkage and selection operator (LASSO) algorithm with ten-fold cross-validation to identify the proteins that best predicted the measured obesity-related phenotype. For each obesity-related trait, the LASSO-selected proteins were used for the pathway enrichment for gene ontology using the topGO package.^12^ Multiple testing correction for the pathways were performed using a false discovery rate (FDR).^13^

Additionally, these selected proteins in the training set were used to compute a weighted protein score, namely, protein-predicted score of BMI (PPS_BMI_), BFP (PPS_BFP_), and WHR (PPS_WHR_). The scores were calculated by summing the protein levels weighted by the LASSO-derived beta coefficients, and subsequently transformed into z-scores with a mean of zero and a standard deviation of one. The performance of the PPS in the test set was evaluated using R^2^ in the linear regression of the PPS against the measured phenotype. Furthermore, we examined the association between each LASSO-selected protein and the obesity-related phenotype using linear regression, with the measured obesity-related phenotype regressed on the specific protein.

For risk prediction of MACE incidence and its subcomponents, ischemic stroke, myocardial infarction, and cardiovascular death, we utilized the healthy cohort excluding the training set, plus the participants without prevalent stroke and coronary artery disease at enrollment. In this cohort, the associations between PPS_BMI_, PPS_BFP_, and PPS_WHR_ and outcomes were estimated using Fine and Gray’s competing risk model, treating death as a competing risk.^14^ Three models with hierarchical adjustment were adopted: Model 1 adjusted for age, sex, and race (white vs. other); Model 2 adjusted for the measured obesity-related phenotype (BMI, BFP, or WHR) in addition to Model 1; Model 3 adjusted for additional risk factors in the PREVENT equation,^15^ including total cholesterol, HDL-C, systolic blood pressure, eGFR, diabetes, current smoking, blood pressure lowering medication use, and cholesterol lowering medication use in addition to Model 2. The analyses were conducted in the overall cohort, and sex-stratified groups. Sex interaction with the protein scores was tested using Model 3. Additionally, a sensitivity analysis was performed after excluding 3,736 individuals with prevalent cancer.

C statistics were calculated to evaluate the performance of the protein scores in predicting MACE. Each protein score was evaluated individually and together, in both an unadjusted model and a model adjusted for age, sex, and current smoking. C statistics were also calculated for the PREVENT equation model^15^ to compare the predictive performance.

The overall study workflow is shown in **Supplemental Figure 1**. All data analyses were conducted using R version 4.4.0. Statistical significance was based on P values < 0.05.

## RESULTS

Among 40,651 participants with proteomic data, a total of 4,071 (10.0%) developed incident MACE over a median follow-up of 14.5 (interquartile range 13.7 - 15.2) years, including 781 (19.2%) incident ischemic stroke, 3,096 (76.1%) MI, and 978 (24.0%) cardiovascular death events, **Table 1**. Compared to the cohort without incident MACE, the MACE cohort was older, and had a higher proportion of males, and more likely to have cardiovascular risk factors, except for lower total cholesterol levels, **Table 1**. In the analysis of obesity-related phenotypes stratified by sex, participants who developed MACE had higher BMI, BFP and WHR compared to participants without MACE among men and women, **Table 2**.

**Table 1.** Cohort Characteristics Among the UK Biobank Participants.

| **Variables** | **Overall**<br>**N=40651** | **No MACE**<br>**N=36580 (90.0%)** | **MACE**<br>**N=4071 (10.0%)** | **P Value**<br>**No MACE**<br>**vs.**<br>**MACE** |
| --- | --- | --- | --- | --- |
| **Age (years)** | 56.44 (8.19) | 55.98 (8.20) | 60.60 (6.89) | **<0.001** |
| **Male (%)** | 18026 (44.3) | 15525 (42.4) | 2501 (61.4) | **<0.001** |
| **White (%)** | 37997 (93.5) | 34171 (93.4) | 3826 (94.0) | 0.175 |
| **BMI (kg/m<sup>2</sup>)** | 27.32 (4.74) | 27.19 (4.69) | 28.46 (5.04) | **<0.001** |
| **Body Fat Percentage (%)** | 31.46 (8.57) | 31.51 (8.58) | 31.00 (8.42) | **<0.001** |
| **Waist-hip Ratio** | 0.87 (0.09) | 0.86 (0.09) | 0.91 (0.09) | **<0.001** |
| **Total Cholesterol (mg/dL)** | 221.97 (43.61) | 222.17 (43.22) | 220.19 (46.92) | **0.007** |
| **HDL-C (mg/dL)** | 56.48 (14.79) | 56.91 (14.75) | 52.64 (14.61) | **<0.001** |
| **Systolic Blood Pressure (mmHg)** | 139.63 (19.64) | 138.88 (19.49) | 146.35 (19.76) | **<0.001** |
| **eGFR (mL/min/1.73m<sup>2</sup>)** | 94.88 (13.07) | 95.30 (12.81) | 91.13 (14.66) | **<0.001** |
| **Diabetes Prevalence (%)** | 1845 (4.5) | 1419 (3.9) | 426 (10.5) | **<0.001** |
| **Current Smoking (%)** | 4276 (10.6) | 3615 (9.9) | 661 (16.3) | **<0.001** |
| **Blood Pressure Lowering Medication Use (%)** | 7530 (18.7) | 6220 (17.2) | 1310 (32.7) | **<0.001** |
| **Cholesterol Lowering Medication Use (%)** | 5702 (14.2) | 4638 (12.8) | 1064 (26.5) | **<0.001** |
| **Ischemic Stroke Incidence (%)** | 781(1.9) | 0 | 781(19.2) | - |
| **MI Incidence (%)** | 3096 (7.6) | 0 | 3096 (76.1) | - |
| **CV Death (%)** | 978 (2.4) | 0 | 978 (24.0) | - |

Mean (standard deviation) shown unless stated for continuous variables. Count (percentage) shown for categorical variables.
BMI, body mass index; HDL-C, high density lipoprotein cholesterol; eGFR, estimated glomerular filtration rate; MACE: major adverse cardiovascular events (ischemic stroke, myocardial infarction, and cardiovascular death); MI: myocardial infarction; CV: cardiovascular.

**Table 2.** Obesity-related Phenotypes Stratified by Sex Among the UK Biobank Participants.

| Obesity-related Phenotype | Sex | Overall | No MACE | MACE | P Value<br>No MACE<br>vs.<br>MACE |
| --- | --- | --- | --- | --- | --- |
| BMI (kg/m<sup>2</sup>) | Male | 27.68 (4.14) | 27.53 (4.06) | 28.58 (4.52) | <0.001 |
|  | Female | 27.03 (5.16) | 26.94 (5.10) | 28.28 (5.76) | <0.001 |
| Body Fat Percentage (%) | Male | 25.09 (5.77) | 24.85 (5.71) | 26.58 (5.87) | <0.001 |
|  | Female | 36.53 (6.88) | 36.42 (6.86) | 38.04 (6.94) | <0.001 |
| Waist-hip Ratio | Male | 0.93 (0.06) | 0.93 (0.06) | 0.95 (0.06) | <0.001 |
|  | Female | 0.82 (0.07) | 0.82 (0.07) | 0.84 (0.07) | <0.001 |

Mean (standard deviation) shown. BMI, body mass index; MACE: major adverse cardiovascular events (ischemic stroke, myocardial infarction, and cardiovascular death).

Based on the comparison of R^2^ values assessing the prediction performance of PPS_BMI_, PPS_BFP_, and PPS_WHR_ across various sample sizes within the training set, we observed a consistent improvement in R^2^ from 10% to 50% of the sample size in the healthy cohort. The performance remained stable from 50% to 90% of the sample size. Since there was no meaningful difference in performance between the 50% and 90% sample sizes among the healthy cohort, we chose to use 50% of the total healthy cohort (N=7,826) as the final training set for fitting LASSO to conduct protein selection for each obesity-related phenotype, **Supplemental Figure 2 and Supplemental Table 1**. Then, we randomly divided the healthy cohort into two subsets: a training set and a test set, each comprising 7,826 participants.

In the training set of the healthy cohort, LASSO models selected 389, 385, and 176 proteins for prediction of BMI, BFP and WHR, respectively. The associations between individual proteins and measured obesity-related phenotypes are shown in **Supplemental Tables 2A-2C.** For BMI, 219 proteins (56.3%) showed positive beta coefficients and 340 are statistically significant at p <0.05. For BFP, 219 proteins (56.9%) showed positive associations and 329 (85.5%) are significant. For WHR, 91 proteins (51.7) were positively associated and 168 (95.5%) were statistically significant. Across these LASSO-selected proteins, a total of 213, 226, and 76 distinct proteins were uniquely selected for BMI, BFP, and WHR prediction, respectively. Notably, 25 proteins were selected for prediction models across all three obesity traits, **Supplemental Tables 2D and Supplemental Figure 3**. Pathway enrichment for gene ontology using the proteins selected for BMI and WHR did not reveal significant pathways after FDR correction of multiple testing. The selected proteins for BFP resulted in two pathways with FDR corrected q value < 0.1, including cell adhesion and extracellular matrix organization, **Supplemental Tables 3A-3C**. In the test set of the healthy cohort plus the MACE prediction cohort, the PPS_BMI_, PPS_BFP_, and PPS_WHR_ scores were significantly correlated with the measured phenotypes, with R^2^ of 0.78, 0.85 and 0.63, respectively, **Supplemental Figure 4**.

The three protein scores were statistically associated with MACE and the subcomponents of MACE in Model 1 adjusting for age, sex and race. The associations remained statistically significant for MACE and MI in Model 2 additionally adjusting for the measured BMI, BFP, or WHR, **Table 3**. In the full Model 3, a standard deviation (SD) increment in PPS_BFP_ and PPS_WHR_ was significantly associated with higher risk for MACE (HR 1.25, 95% CI 1.14 - 1.38, p <0.0001; HR 1.15, 95% CI 1.06 - 1.24, p = 0.001, respectively), whereas PPS_BMI_ showed a nominal association (HR 1.08, 95% CI 1.00 – 1.17, p = 0.0524), **Table 3 and Figure 1**. In Model 3, all 3 scores remained significantly associated with MI, while PPS_BFP_ remained associated with cardiovascular death, **Table 3 and Supplemental Figure 5**. The sensitivity analysis after excluding prevalent cancer resulted in similar findings, **Supplemental Table 4**.

**Figure 1.**
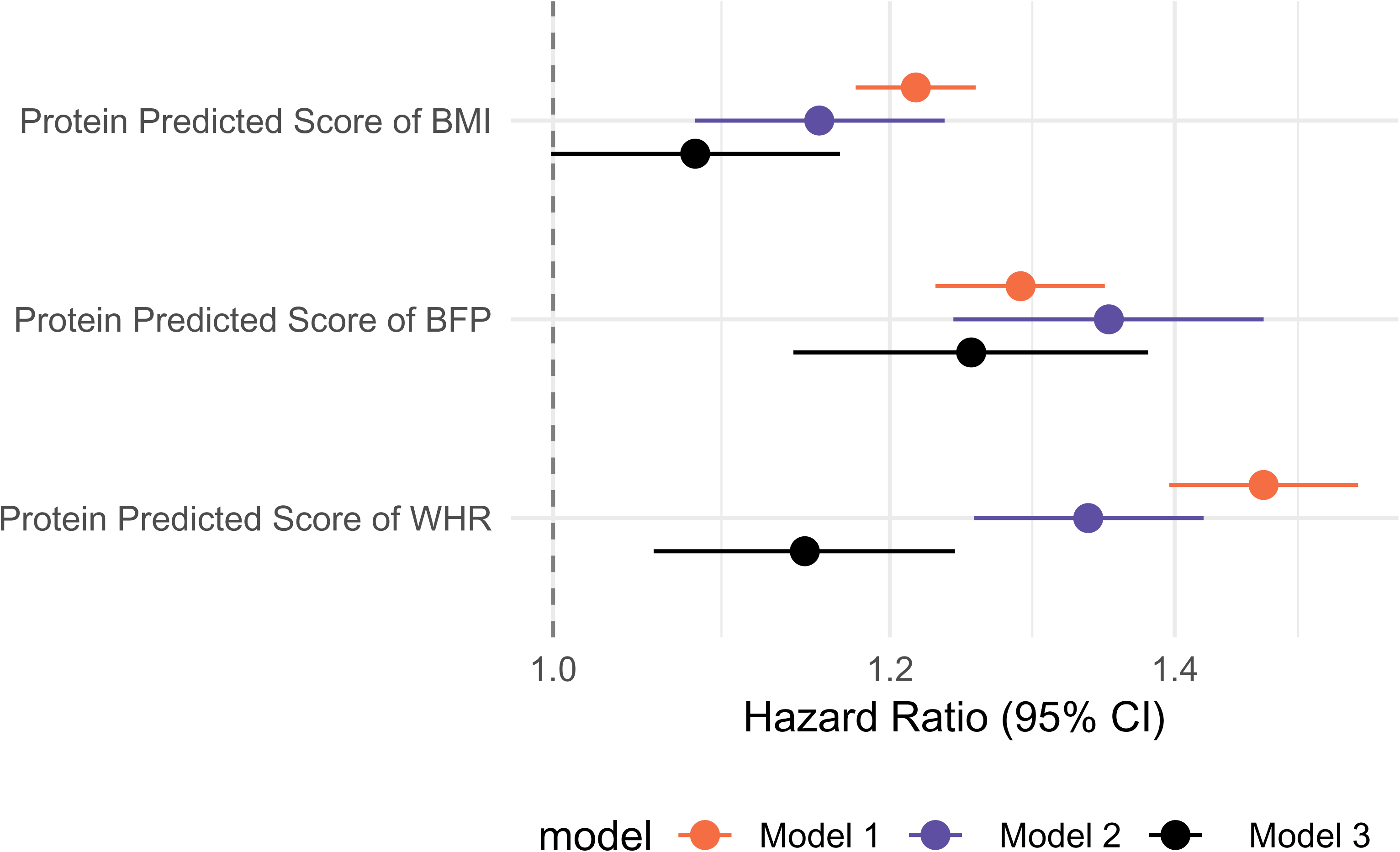
Forest Plot of the Associations Between Protein Predicted Scores of Obesity-related Phenotypes and MACE Outcomes. Model 1: adjusted for age, sex and race (white vs. other); Model 2: adjusted for the measured obesity-related phenotype (BMI, body fat percentage, or waist-hip ratio) in addition to Model 1; Model 3: adjusted for total cholesterol, high density lipoprotein cholesterol, systolic blood pressure, estimated glomerular filtration rate calculated using the 2021 CKD-EPI equation, diabetes, current smoking, blood pressure lowering medication use, cholesterol lowering medication use in addition to Model 2.

**Table 3.**
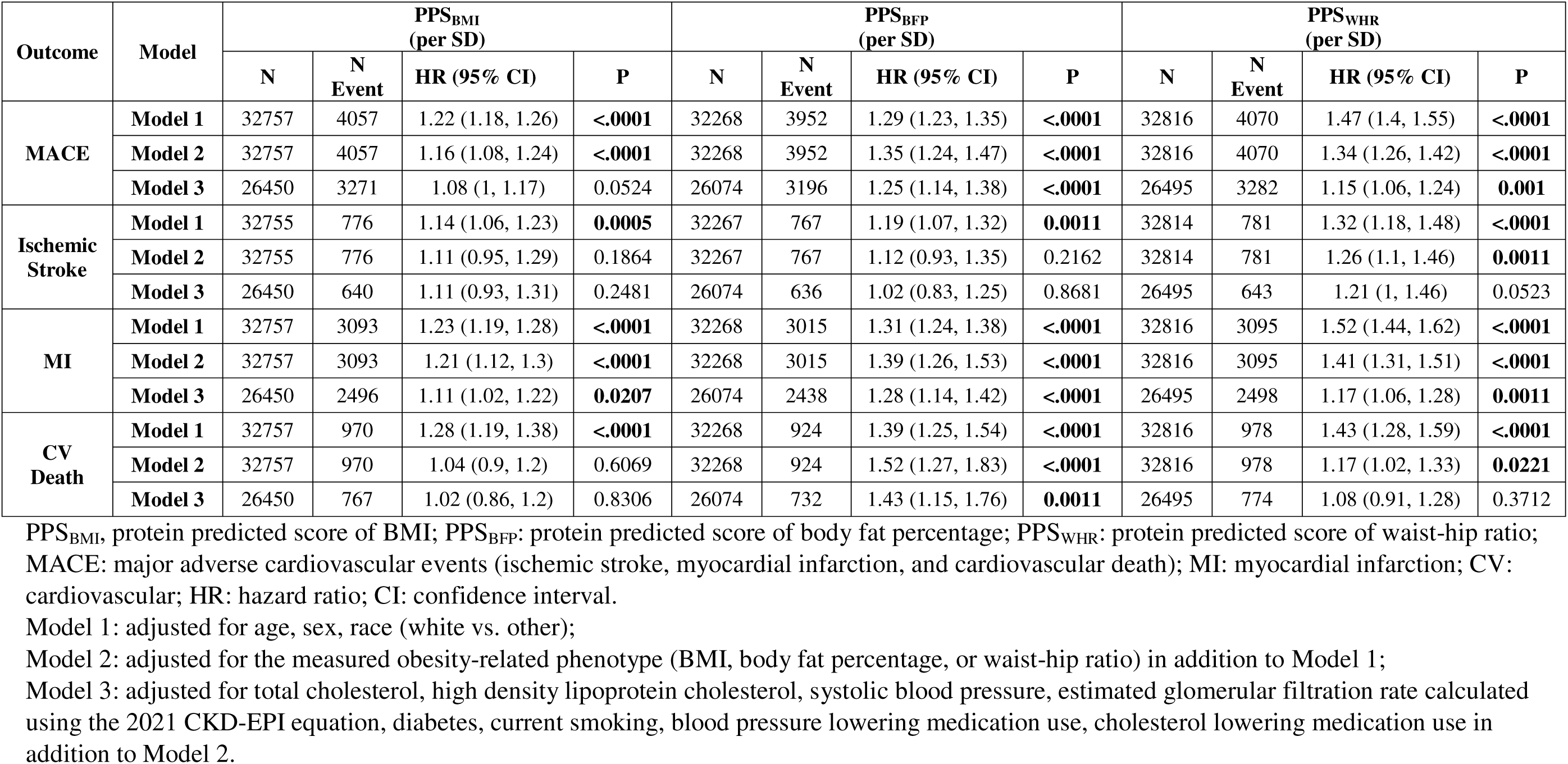
Associations Between Protein Predicted Scores and Outcomes.

Statistically significant sex interaction with the protein scores was not identified. In the sex-stratified analysis, the associations between PPS_BMI_ and MACE and MI, the association between PPS_BFP_ and cardiovascular death was significant only among males, while the association between PPS_WHR_ and ischemic stroke was significant only among females. Consistent associations were found among both sex groups between PPS_BFP_ and MACE and MI, and between PPS_WHR_ and MACE and MI, **Supplemental Table 5**. Sensitivity analyses that excluded individuals with a history of cancer, showed similar results. **Supplemental Table 6**.

The individual proteins scores PPS_BMI_, PPS_BFP_ and PPS_WHR_ had C statistics of 0.557, 0.529, and 0.626 for predicting MACE, respectively, **Table 4**. The combination of three protein scores showed a C statistic of 0.634. Compared with the fully adjusted PREVENT equation model^15^ with C statistic 0.694, the models adjusted for only age, sex, current smoking and individual proteins scores showed comparable performance (PPS_BMI_ 0.685, PPS_BFP_ 0.684, PPS_WHR_ 0.687). The model adjusted for age, sex, current smoking and all three protein scores showed a C statistic of 0.688, **Table 4**.

**Table 4.**
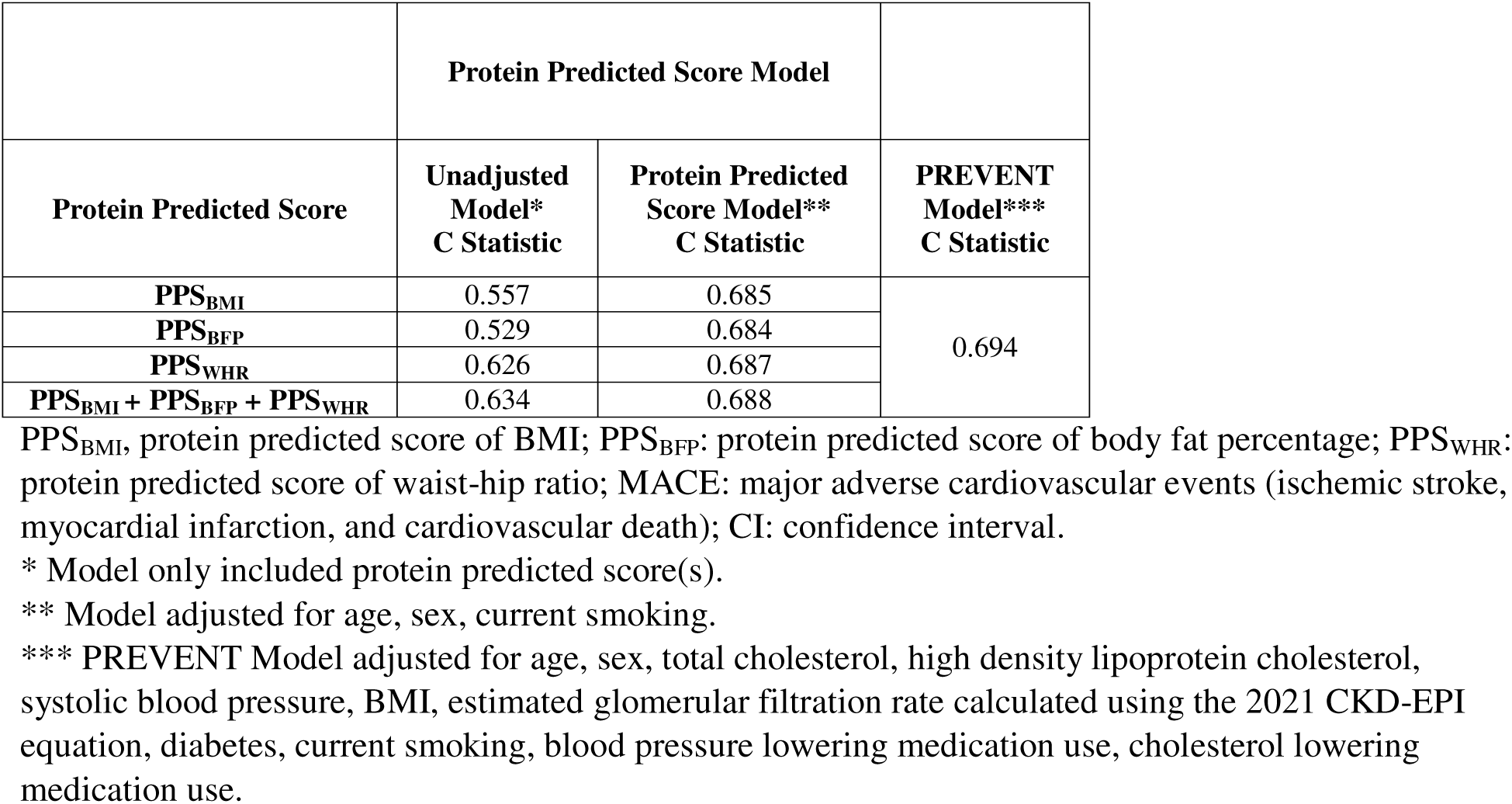
Prediction Performance for MACE.

## DISCUSSION

This study explored the capacity of proteomic profiles in estimating obesity-related phenotypes and assessed their associations with MACE. The protein-predicted obesity-related phenotypes - BMI, BFP, WHR were strongly correlated with their measured counterparts, suggesting that proteomic profiles capture the complex molecular underpinnings of obesity, potentially providing a more nuanced understanding beyond traditional metrics. Higher protein-predicted scores for BMI, BFP, and WHR were associated with an increased risk of MACE, even after adjusting for established cardiovascular risk factors. Our findings underscore the potential utility of proteomic data to help characterize the biological impact of adiposity in the prediction of cardiovascular events.

The protein predicted score of BFP had a strong correlation with the measured BFP (R^2^ =0.85), which is comparable to the findings in the Fenland study using aptamer technology and the SomaScan assay (R^2^ =0.92).^16^ This result may indicate that certain proteins could play more significant roles in adipogenesis, lipid metabolism, or energy homeostasis, impacting BFP more strongly.^17–19^ On the other hand, BMI depends on multiple tissue compositions such as muscle mass, bone density, and fat distribution. Consequently, the protein profile associated with BMI can be more complex, influencing multiple tissue types and physiological processes beyond adiposity alone.^20^ Additionally, WHR may also be affected by external factors such as measurement error.^21^ The complexities of WHR measurements, including variations in body shape and individual differences in skeletal structure, can introduce additional challenges when correlating proteomic data with this phenotype. The 25 proteins associated with all three obesity phenotypes highlight the multifaceted nature of obesity and emphasize the importance of diverse biological pathways in understanding and addressing this complex condition. For example, GHRL is crucial for regulating appetite and energy balance, thereby influencing body weight and fat distribution.^22^ CFH and AGER may intersect with obesity through their impact on insulin resistance.^23–25^ BAG3 plays a role in cell survival and responses to stress, making it relevant in various diseases, including metabolic disorders.^26^ LEP is shared among the proteins selected from LASSO for both BMI and BFP. Leptin resistance, marked by reduced satiety, often leads to obesity.^27^

In this prospective population-based study, even with the comprehensive adjustment for risk factors in the PREVENT equation,^15^ in addition to the measured obesity-related phenotypes, the protein predicted scores of BFP and WHR consistently demonstrated robust associations with MACE and MI. This finding underscores the potential utility of proteomic data to enhance risk prediction models for incident cardiovascular disease. Even though the PPS-sex interactions didn’t reach statistical significance, in the sex-stratified analysis, the associations of PPS_BMI_ with MACE and MI were only significant in males. A similar pattern was noted for PPS_BFP_ in relation to cardiovascular death. These findings are consistent with earlier studies that have reported sex differences in the impact of obesity on adverse cardiovascular outcomes.^28^ Additionally, a sex difference was observed in the association between PPS_WHR_ with ischemic stroke, with significant associations observed only among females. This result is consistent with prior research indicating that women demonstrate a greater excess risk of MI with increased waist circumference and WHR compared to men.^29^ Differences in body composition and fat distribution are influenced by sex hormones, with women typically exhibiting higher fat mass and subcutaneous fat, while men tend to have more lean mass and visceral fat.^29,30^ This underscores the importance of considering sex-specific differences in cardiovascular risk prediction and highlights the potential of proteomic data in elucidating sex-specific molecular mechanisms underlying cardiovascular disease.

Our study also demonstrates that models incorporating age, sex, smoking status, and obesity-related protein scores can predict MACE with performance comparable to the PREVENT equation model,^15^ a widely used tool for cardiovascular risk prediction. The advantage of using protein scores derived from obesity-related phenotypes is that they provide a direct biological assessment of obesity risk, which contributes to several cardiovascular risk factors in the PREVENT model, such as blood pressure, lipids, and diabetes. The ability of protein scores to achieve similar predictive performance to the PREVENT model,^15^ while requiring only basic demographic and smoking information, has significant practical implications. Age, sex, and smoking status are easily collected in clinical practice, and adding protein scores could offer a more personalized, biologically relevant cardiovascular risk assessment. This combination of simple clinical data with proteomic markers could make cardiovascular risk stratification easier, more cost-effective, and a valuable tool for clinical decision-making.

The study of the large biobank cohort with well-characterized phenotypes, comprehensive proteomic data, and long-term follow-up highlights the potential of proteomic profiles in predicting obesity-related phenotypes and their implications for cardiovascular risk prediction. However, several limitations warrant consideration. Our analysis focused on participants from the UK Biobank cohort, a population of predominantly European descent, which may limit the generalizability of our findings to other populations. While proteomic profiling offers valuable insights into molecular pathways associated with obesity and cardiovascular disease, future research should focus on validating these findings in diverse populations and elucidating the molecular mechanisms linking proteomic profiles with adverse cardiovascular outcomes.

Ultimately, integrating proteomic data into clinical practice may enhance risk stratification and facilitate personalized preventive strategies for individuals at high risk of cardiovascular disease.

## CONCLUSION

The protein-predicted scores for obesity-related phenotypes provide a potentially more refined approach to capturing the complexities of adiposity than directly measured metrics. By incorporating protein scores of obesity traits with easily obtained clinical variables, such as age, sex, and smoking status, we can achieve predictive performance for incident MACE comparable to the established PREVENT equation, offering a single-assay-based alternative and biologically relevant assessment of risk. This approach can potentially simplify the procedure of risk assessment, reduce measurement errors, and be easily integrated into routine practice, making it both cost-effective and accessible to a wide range of patient populations.

## Data Availability

The UK Biobank will make the data available to all bona fide researchers for all types of health-related research that is in the public interest, without preferential or exclusive access for any persons. All researchers will be subject to the same application process and approval criteria as specified by UK Biobank. For more details on the access procedure, see the UK Biobank website: www.ukbiobank.ac.uk.

## CONTRIBUTORS

CL: conceptualization, data curation, formal analysis, writing original draft. BS: data curation, formal analysis, review and editing. QH: data curation. PWFW: investigation, review and editing. AAQ: investigation, review and editing. YVS: resources, conceptualization, investigation, review and editing, supervision. All authors approved the manuscript before submission.

## ACKNOWLEDGEMENT

The research was conducted using data from the UK Biobank Resource under application number 34031. This research is supported by the National Heart, Lung, And Blood Institute, National Institutes of Health, Award Number P01HL154996.

---

**Supplemental Figure 1.** Description of the sample selection workflow from the UK Biobank cohort.

**Supplemental Figure 2.** R^2^ values assessing the prediction performance of protein-predicted scores of BMI (PPS_BMI_), BFP (PPS_BFP_), and WHR (PPS_WHR_) across various sample sizes. The median R^2^ from the LASSO models with 2.5% and 97.5% percentiles of the 100 iterations are shown.

**Supplemental Figure 3.** The LASSO Selected Proteins Shared across Obesity-related Phenotypes.

**Supplemental Figure 4.** Linear Associations Between Predicted Protein Scores of Obesity-related Phenotypes and Measured Phenotypes.

**Supplemental Figure 5.** Forest Plot of the Associations Between Protein Predicted Scores of Obesity-related Phenotypes and MACE Individual Components. Model 1: adjusted for age, sex and race (white vs. other); Model 2: adjusted for the measured obesity-related phenotype (BMI, body fat percentage, or waist-hip ratio) in addition to Model 1; Model 3: adjusted for total cholesterol, high density lipoprotein cholesterol, systolic blood pressure, estimated glomerular filtration rate calculated using the 2021 CKD-EPI equation, diabetes, current smoking, blood pressure lowering medication use, cholesterol lowering medication use in addition to Model 2.

## REFERENCES

1. Hales CM, Carroll MD, Fryar CD, Ogden CL. Prevalence of Obesity and Severe Obesity Among Adults: United States, 2017-2018. NCHS Data Brief 2020; (360): 1–8.

2. Powell-Wiley TM, Poirier P, Burke LE, et al. Obesity and Cardiovascular Disease: A Scientific Statement From the American Heart Association. Circulation 2021; 143(21): e984–e1010.

3. Frankenfield DC, Rowe WA, Cooney RN, Smith JS, Becker D. Limits of body mass index to detect obesity and predict body composition. Nutrition 2001; 17(1): 26–30.

4. Cheng CH, Ho CC, Yang CF, Huang YC, Lai CH, Liaw YP. Waist-to-hip ratio is a better anthropometric index than body mass index for predicting the risk of type 2 diabetes in Taiwanese population. Nutr Res 2010; 30(9): 585–93.

5. Goonasegaran AR, Nabila FN, Shuhada NS. Comparison of the effectiveness of body mass index and body fat percentage in defining body composition. Singapore Med J 2012; 53(6): 403–8.

6. Zaghlool SB, Sharma S, Molnar M, et al. Revealing the role of the human blood plasma proteome in obesity using genetic drivers. Nat Commun 2021; 12(1): 1279.

7. Helgason H, Eiriksdottir T, Ulfarsson MO, et al. Evaluation of Large-Scale Proteomics for Prediction of Cardiovascular Events. JAMA 2023; 330(8): 725–35.

8. Nurmohamed NS, Belo Pereira JP, Hoogeveen RM, et al. Targeted proteomics improves cardiovascular risk prediction in secondary prevention. Eur Heart J 2022; 43(16): 1569–77.

9. Sudlow C, Gallacher J, Allen N, et al. UK biobank: an open access resource for identifying the causes of a wide range of complex diseases of middle and old age. PLoS Med 2015; 12(3): e1001779.

10. Sun BB, Chiou J, Traylor M, et al. Plasma proteomic associations with genetics and health in the UK Biobank. Nature 2023; 622(7982): 329–38.

11. Inker LA, Eneanya ND, Coresh J, et al. New Creatinine- and Cystatin C-Based Equations to Estimate GFR without Race. N Engl J Med 2021; 385(19): 1737–49.

12. Alexa A, Rahnenfuhrer J, Lengauer T. Improved scoring of functional groups from gene expression data by decorrelating GO graph structure. Bioinformatics 2006; 22(13): 1600–7.

13. Benjamini Y, Hochberg Y. Controlling the False Discovery Rate: A Practical and Powerful Approach to Multiple Testing. Journal of the Royal Statistical Society: Series B (Methodological) 1995; 57(1): 289–300.

14. Fine JP, Gray RJ. A Proportional Hazards Model for the Subdistribution of a Competing Risk. Journal of the American Statistical Association 1999; 94(446): 496–509.

15. Khan SS, Matsushita K, Sang Y, et al. Development and Validation of the American Heart Association’s PREVENT Equations. Circulation 2024; 149(6): 430–49.

16. Williams SA, Kivimaki M, Langenberg C, et al. Plasma protein patterns as comprehensive indicators of health. Nat Med 2019; 25(12): 1851–7.

17. Konige M, Wang H, Sztalryd C. Role of adipose specific lipid droplet proteins in maintaining whole body energy homeostasis. Biochim Biophys Acta 2014; 1842(3): 393–401.

18. Huemer MT, Bauer A, Petrera A, et al. Proteomic profiling of low muscle and high fat mass: a machine learning approach in the KORA S4/FF4 study. J Cachexia Sarcopenia Muscle 2021; 12(4): 1011–23.

19. Liao J, Goodrich JA, Chen W, et al. Cardiometabolic profiles and proteomics associated with obesity phenotypes in a longitudinal cohort of young adults. Sci Rep 2024; 14(1): 7384.

20. Goudswaard LJ, Bell JA, Hughes DA, et al. Effects of adiposity on the human plasma proteome: observational and Mendelian randomisation estimates. Int J Obes (Lond) 2021; 45(10): 2221–9.

21. Sebo P, Herrmann FR, Haller DM. Accuracy of anthropometric measurements by general practitioners in overweight and obese patients. BMC Obes 2017; 4: 23.

22. Abizaid A, Horvath TL. Ghrelin and the central regulation of feeding and energy balance. Indian J Endocrinol Metab 2012; 16(Suppl 3): S617–26.

23. Moreno-Navarrete JM, Martinez-Barricarte R, Catalan V, et al. Complement factor H is expressed in adipose tissue in association with insulin resistance. Diabetes 2010; 59(1): 200–9.

24. Portero-Otin M, de la Maza MP, Uribarri J. Dietary Advanced Glycation End Products: Their Role in the Insulin Resistance of Aging. Cells 2023; 12(13).

25. Asadipooya K, Uy EM. Advanced Glycation End Products (AGEs), Receptor for AGEs, Diabetes, and Bone: Review of the Literature. J Endocr Soc 2019; 3(10): 1799–818.

26. Rosati A, Graziano V, De Laurenzi V, Pascale M, Turco MC. BAG3: a multifaceted protein that regulates major cell pathways. Cell Death Dis 2011; 2(4): e141.

27. Obradovic M, Sudar-Milovanovic E, Soskic S, et al. Leptin and Obesity: Role and Clinical Implication. Front Endocrinol (Lausanne) 2021; 12: 585887.

28. Global BMIMC, Di Angelantonio E, Bhupathiraju Sh N, et al. Body-mass index and all-cause mortality: individual-participant-data meta-analysis of 239 prospective studies in four continents. Lancet 2016; 388(10046): 776–86.

29. Peters SAE, Bots SH, Woodward M. Sex Differences in the Association Between Measures of General and Central Adiposity and the Risk of Myocardial Infarction: Results From the UK Biobank. J Am Heart Assoc 2018; 7(5).

30. Valencak TG, Osterrieder A, Schulz TJ. Sex matters: The effects of biological sex on adipose tissue biology and energy metabolism. Redox Biol 2017; 12: 806–13.

