## Supplemental Material for "Proteomic Profiles of Obesity-Related Phenotypes and Incident Cardiovascular Events"

**Supplemental Table 3C. Pathway Enrichment for Gene Ontology Using the 176 LASSO-selected Proteins for Waist-hip Ratio. Top 10 Pathways out of Total 57 Pathways with $p < 0.05$ are shown.** 33

**Supplemental Figure 5. Forest Plot of the Associations Between Protein Predicted Scores of Obesity-related Phenotypes and MACE Individual Components. Model 1: adjusted for age, sex and race (white vs. other); Model 2: adjusted for the measured obesity-related phenotype (BMI, body fat percentage, or waist-hip ratio) in addition to Model 1; Model 3: adjusted for total cholesterol, high density lipoprotein cholesterol, systolic blood pressure, estimated glomerular filtration rate**

**calculated using the 2021 CKD-EPI equation, diabetes, current smoking, blood pressure lowering medication use, cholesterol lowering medication use in addition to Model 2.** ..................................... 43

**Supplemental Table 1. Performance of the Protein Predicted Scores by Different Sample Sizes of Training set.**

| Protein Predicted Score | Training Set Sample Size (%) | Median $R^2$ | Percentile 2.5% | Percentile 97.5% |
|---|---|---|---|---|
| **$PPS_{BMI}$** | 1564 (10%) | 0.7110 | 0.6969 | 0.7231 |
| | 3127 (20%) | 0.7399 | 0.7309 | 0.7474 |
| | 4690 (30%) | 0.7513 | 0.7433 | 0.7587 |
| | 6254 (40%) | 0.7588 | 0.7503 | 0.7672 |
| | 7817 (50%) | 0.7646 | 0.7558 | 0.7720 |
| | 9380 (60%) | 0.7677 | 0.7593 | 0.7762 |
| | 10944 (70%) | 0.7708 | 0.7610 | 0.7809 |
| | 12507 (80%) | 0.7733 | 0.7618 | 0.7857 |
| | 14070 (90%) | 0.7763 | 0.7568 | 0.7937 |
| **$PPS_{BFP}$** | 1548 (10%) | 0.8236 | 0.8148 | 0.8308 |
| | 3096 (20%) | 0.8358 | 0.8310 | 0.8400 |
| | 4644 (30%) | 0.8418 | 0.8378 | 0.8460 |
| | 6191 (40%) | 0.8466 | 0.8408 | 0.8509 |
| | 7739 (50%) | 0.8496 | 0.8442 | 0.8549 |
| | 9287 (60%) | 0.8523 | 0.8471 | 0.8575 |
| | 10834 (70%) | 0.8535 | 0.8465 | 0.8600 |
| | 12382 (80%) | 0.8553 | 0.8465 | 0.8647 |
| | 13930 (90%) | 0.8577 | 0.8411 | 0.8691 |
| **$PPS_{WHR}$** | 1566 (10%) | 0.5771 | 0.5624 | 0.5884 |
| | 3131 (20%) | 0.5952 | 0.5868 | 0.6016 |
| | 4696 (30%) | 0.6041 | 0.5961 | 0.6116 |
| | 6261 (40%) | 0.6097 | 0.5992 | 0.6162 |
| | 7826 (50%) | 0.6125 | 0.6034 | 0.6237 |
| | 9391 (60%) | 0.6158 | 0.6035 | 0.6264 |
| | 10956 (70%) | 0.6181 | 0.6017 | 0.6318 |
| | 12521 (80%) | 0.6202 | 0.6027 | 0.6394 |
| | 14086 (90%) | 0.6209 | 0.5906 | 0.6507 |

$PPS_{BMI}$, protein predicted score of BMI; $PPS_{BFP}$: protein predicted score of body fat percentage; $PPS_{WHR}$: protein predicted score of waist-hip ratio.

**Supplemental Table 2A. Association between the 389 LASSO Selected Proteins and BMI in the Training Set of the Healthy Cohort.**

| Protein | Name | LASSO | Linear Regression* | |
|---|---|---|---|---|
| | | Beta | Beta (95% CI) | P |
| ACAN | Aggrecan core protein | -0.0077 | -0.49 (-0.59, -0.4) | 1.30E-24 |
| ACE2 | Angiotensin-converting enzyme 2 | 0.1393 | 0.86 (0.76, 0.96) | 1.20E-65 |
| ACRV1 | Acrosomal protein SP-10 | 0.0604 | 0.36 (0.27, 0.46) | 3.20E-13 |
| ADA | Adenosine deaminase | 0.0088 | 0.55 (0.46, 0.64) | 2.00E-32 |
| ADAM12 | Disintegrin and metalloproteinase domain-containing protein 12 | 0.003 | 0.89 (0.79, 0.98) | 6.20E-75 |
| ADAM15 | Disintegrin and metalloproteinase domain-containing protein 15 | -0.0319 | -0.29 (-0.38, -0.19) | 1.90E-09 |
| ADAMTS15 | A disintegrin and metalloproteinase with thrombospondin motifs 15 | 0.1426 | 1.67 (1.58, 1.76) | 2.10E-259 |
| ADAMTS16 | A disintegrin and metalloproteinase with thrombospondin motifs 16 | 0.0014 | 0.21 (0.12, 0.31) | 1.10E-05 |
| ADAMTS8 | A disintegrin and metalloproteinase with thrombospondin motifs 8 | -0.0136 | -0.53 (-0.63, -0.44) | 1.50E-29 |
| ADGRB3 | Adhesion G protein-coupled receptor B3 | -0.0003 | -0.57 (-0.67, -0.48) | 7.60E-33 |
| ADGRG2 | Adhesion G-protein coupled receptor G2 | -0.1181 | -0.97 (-1.06, -0.88) | 3.60E-92 |
| ADM | Pro-adrenomedullin | 0.2999 | 1.79 (1.7, 1.88) | 7.00E-289 |
| AFP | Alpha-fetoprotein | -0.0106 | -0.12 (-0.22, -0.03) | 0.0088 |
| AGER | Advanced glycosylation end product-specific receptor | -0.1281 | -0.84 (-0.93, -0.74) | 1.60E-68 |
| AGT | Angiotensinogen | -0.0175 | -0.19 (-0.28, -0.1) | 0.0001 |
| AKR1C4 | Aldo-keto reductase family 1 member C4 | 0.01 | 0.32 (0.22, 0.41) | 2.70E-11 |
| AMBP | Protein AMBP | 0.0082 | 1.1 (1.01, 1.2) | 5.10E-111 |
| ANGPTL1 | Angiopoietin-related protein 1 | -0.0601 | -0.44 (-0.53, -0.35) | 3.70E-20 |
| ANGPTL2 | Angiopoietin-related protein 2 | 0.0292 | 1.12 (1.03, 1.22) | 7.10E-120 |
| ANGPTL7 | Angiopoietin-related protein 7 | -0.1052 | -0.14 (-0.23, -0.04) | 0.0052 |
| ANXA2 | Annexin A2 | 0.0023 | 0.72 (0.63, 0.81) | 1.40E-51 |
| APCS | Serum amyloid P-component | 0.0067 | 1.78 (1.7, 1.86) | 0.00E+00 |
| APOA1 | Apolipoprotein A-I | -0.0865 | -0.76 (-0.86, -0.67) | 8.30E-60 |
| APOA4 | Apolipoprotein A-IV | -0.0019 | -0.23 (-0.32, -0.14) | 1.60E-06 |
| APOF | Apolipoprotein F | -0.0009 | -1.4 (-1.49, -1.31) | 6.90E-190 |
| APOL1 | Apolipoprotein L1 | 0.0119 | 0.6 (0.5, 0.69) | 6.10E-36 |
| APOM | Apolipoprotein M | -0.0177 | -0.52 (-0.61, -0.42) | 7.70E-27 |
| AREG | Amphiregulin | -0.0525 | -0.22 (-0.32, -0.13) | 1.90E-06 |
| ARG2 | Arginase-2, mitochondrial | 0.0334 | 0.09 (0, 0.18) | 0.0457 |
| ARSA | Arylsulfatase A | -0.0121 | 0.72 (0.63, 0.82) | 1.40E-51 |
| ART3 | Ecto-ADP-ribosyltransferase 3 | -0.1188 | -0.46 (-0.56, -0.37) | 2.20E-21 |
| B4GAT1 | Beta-1,4-glucuronyltransferase 1 | -0.1476 | -0.8 (-0.89, -0.71) | 1.50E-64 |
| BAG3 | BAG family molecular chaperone regulator 3 | 0.0124 | 1.1 (1, 1.19) | 1.40E-115 |
| BCHE | Cholinesterase | 0.0211 | 1.04 (0.95, 1.14) | 1.40E-106 |
| BCL2 | Apoptosis regulator Bcl-2 | -0.0023 | 0.68 (0.59, 0.77) | 3.20E-46 |
| BCL2L11 | Bcl-2-like protein 11, Isoform BimL | -0.0144 | 0 (-0.09, 0.1) | 0.9608 |
| BOC | Brother of CDO | -0.0044 | -0.35 (-0.44, -0.26) | 3.80E-14 |
| BPIFB2 | BPI fold-containing family B member 2 | 0.0141 | 1.52 (1.43, 1.61) | 6.40E-226 |
| BRK1 | Protein BRICK1 | 0.0015 | 0.42 (0.32, 0.51) | 4.50E-18 |

| | | | | |
|---|---|---|---|---|
| BSG | Basigin | 0.0491 | 0.94 (0.85, 1.04) | 2.00E-81 |
| C1QTNF6 | Complement C1q tumor necrosis factor-related protein 6 | 0.012 | 0.26 (0.17, 0.36) | 2.50E-08 |
| C7 | Complement component C7 | -0.0478 | -0.78 (-0.88, -0.68) | 2.30E-55 |
| CA14 | Carbonic anhydrase 14 | 0.032 | -0.92 (-1.02, -0.83) | 1.90E-82 |
| CA4 | Carbonic anhydrase 4 | 0.1754 | 0.3 (0.2, 0.39) | 7.20E-10 |
| CA6 | Carbonic anhydrase 6 | 0.0288 | -0.31 (-0.4, -0.21) | 6.90E-11 |
| CA9 | Carbonic anhydrase 9 | -0.0419 | -1.1 (-1.19, -1) | 7.40E-115 |
| CALB1 | Calbindin | -0.0566 | -0.7 (-0.8, -0.61) | 3.70E-46 |
| CALB2 | Calretinin | 0.0639 | 1.24 (1.15, 1.33) | 1.30E-150 |
| CALCA | Calcitonin | 0.0111 | 0.7 (0.61, 0.79) | 5.60E-48 |
| CALCB | Calcitonin gene-related peptide 2 | -0.0155 | -0.49 (-0.58, -0.4) | 1.70E-24 |
| CAPS | Calcyphosin | 0.0015 | 0.31 (0.22, 0.4) | 4.10E-11 |
| CCL15 | C-C motif chemokine 15 | -0.0479 | 0.22 (0.13, 0.32) | 2.20E-06 |
| CCL19 | C-C motif chemokine 19 | 0.0101 | 0.69 (0.6, 0.78) | 3.50E-50 |
| CCL23 | C-C motif chemokine 23 | -0.0072 | 0.35 (0.26, 0.45) | 3.50E-13 |
| CCL27 | C-C motif chemokine 27 | -0.1521 | 0.01 (-0.08, 0.11) | 0.776 |
| CCL28 | C-C motif chemokine 28 | -0.0381 | -0.53 (-0.63, -0.44) | 2.50E-29 |
| CCN5 | CCN family member 5 | 0.2143 | 1.2 (1.1, 1.3) | 7.80E-130 |
| CD14 | Monocyte differentiation antigen CD14 | -0.0924 | -0.04 (-0.14, 0.06) | 0.4207 |
| CD22 | B-cell receptor CD22 | 0.053 | 0.97 (0.88, 1.06) | 5.80E-89 |
| CD300LG | CMRF35-like molecule 9 | 0.1124 | -0.01 (-0.11, 0.08) | 0.7624 |
| CD36 | Platelet glycoprotein 4 | 0.0336 | 0.49 (0.4, 0.59) | 5.70E-25 |
| CD59 | CD59 glycoprotein | 0.0228 | 1.18 (1.08, 1.27) | 1.10E-116 |
| CD84 | SLAM family member 5 | -0.0025 | -0.01 (-0.1, 0.08) | 0.8093 |
| CD99 | CD99 antigen | 0.0552 | 0.66 (0.56, 0.75) | 8.90E-42 |
| CDA | Cytidine deaminase | 0.0039 | 0.87 (0.78, 0.96) | 5.30E-78 |
| CDCP1 | CUB domain-containing protein 1 | -0.0589 | 0.41 (0.31, 0.5) | 1.30E-15 |
| CDH15 | Cadherin-15 | 0.0118 | 0.56 (0.46, 0.65) | 3.60E-29 |
| CDH6 | Cadherin-6 | -0.0044 | -0.23 (-0.33, -0.14) | 2.20E-06 |
| CDHR1 | Cadherin-related family member 1 | 0.0603 | 0.68 (0.59, 0.77) | 2.30E-46 |
| CDSN | Corneodesmosin | -0.0102 | 0.15 (0.05, 0.24) | 0.002 |
| CEACAM5 | Carcinoembryonic antigen-related cell adhesion molecule 5 | 0.0379 | -0.12 (-0.22, -0.02) | 0.0144 |
| CELSR2 | Cadherin EGF LAG seven-pass G-type receptor 2 | -0.0338 | 0.13 (0.03, 0.22) | 0.0088 |
| CES1 | Liver carboxylesterase 1 | -0.0395 | 1.3 (1.2, 1.39) | 5.30E-153 |
| CES2 | Cocaine esterase | 0.0154 | 0.71 (0.62, 0.8) | 5.30E-51 |
| CES3 | Carboxylesterase 3 | 0.001 | 0.7 (0.61, 0.79) | 4.40E-50 |
| CFD | Complement factor D | 0.0981 | 1.38 (1.29, 1.48) | 2.30E-177 |
| CFH | Complement factor H | 0.1082 | 1.84 (1.75, 1.93) | 9.9e-324 |
| CFHR2 | Complement factor H-related protein 2 | 0.0071 | 0.49 (0.39, 0.58) | 6.60E-23 |
| CFHR4 | Complement factor H-related protein 4 | 0.0024 | 0.4 (0.31, 0.49) | 5.20E-17 |
| CFHR5 | Complement factor H-related protein 5 | 0.0028 | 0.78 (0.68, 0.87) | 9.10E-58 |
| CHCHD10 | Coiled-coil-helix-coiled-coil-helix domain-containing protein 10, mitochondrial | 0.0375 | 1.05 (0.96, 1.14) | 9.90E-104 |

| | | | | |
|---|---|---|---|---|
| CHGB | Secretogranin-1 | -0.0551 | -0.97 (-1.06, -0.87) | 2.00E-87 |
| CHRDL2 | Chordin-like protein 2 | -0.0067 | 0.14 (0.05, 0.23) | 0.0028 |
| CKB | Creatine kinase B-type | -0.1086 | -1.79 (-1.87, -1.7) | 9.9e-324 |
| CKMT1A_CKMT1B | Creatine kinase U-type, mitochondrial | -0.0165 | -0.21 (-0.3, -0.12) | 1.00E-05 |
| CLEC3B | Tetranectin | 0.0303 | 0.47 (0.38, 0.56) | 2.80E-23 |
| CLEC5A | C-type lectin domain family 5 member A | -0.0716 | -0.34 (-0.44, -0.25) | 3.30E-12 |
| CLEC6A | C-type lectin domain family 6 member A | -0.0193 | 0.35 (0.25, 0.44) | 1.40E-12 |
| CLMP | CXADR-like membrane protein | 0.2649 | 1.45 (1.36, 1.54) | 3.10E-196 |
| CLSTN2 | Calsyntenin-2 | -0.0001 | -0.44 (-0.54, -0.35) | 2.00E-20 |
| CLSTN3 | Calsyntenin-3 | 0.0041 | 0.64 (0.55, 0.73) | 4.80E-40 |
| CLUL1 | Clusterin-like protein 1 | -0.0096 | -0.84 (-0.93, -0.74) | 2.70E-70 |
| CNTN3 | Contactin-3 | 0.1141 | 1.16 (1.07, 1.25) | 4.30E-131 |
| CNTN5 | Contactin-5 | -0.0442 | -0.6 (-0.69, -0.5) | 1.20E-35 |
| COL15A1 | Collagen alpha-1(XV) chain | 0.3179 | 1.46 (1.37, 1.55) | 8.70E-202 |
| COL28A1 | Collagen alpha-1(XXVIII) chain | 0.02 | 0.25 (0.16, 0.35) | 1.70E-07 |
| COL4A1 | Collagen alpha-1(IV) chain | -0.1903 | -1.27 (-1.37, -1.18) | 1.10E-157 |
| COL9A1 | Collagen alpha-1(IX) chain | 0.0013 | 0.03 (-0.07, 0.12) | 0.5889 |
| COMP | Cartilage oligomeric matrix protein | 0.086 | 0.75 (0.66, 0.85) | 2.10E-54 |
| COMT | Catechol O-methyltransferase | -0.0136 | 0.29 (0.2, 0.38) | 8.60E-10 |
| CPVL | Probable serine carboxypeptidase CPVL | 0.0209 | 0.24 (0.15, 0.34) | 4.60E-07 |
| CRIM1 | Cysteine-rich motor neuron 1 protein | 0.1755 | 0.02 (-0.08, 0.12) | 0.6784 |
| CRLF1 | Cytokine receptor-like factor 1 | 0.0147 | 0.79 (0.7, 0.88) | 7.30E-64 |
| CRTAC1 | Cartilage acidic protein 1 | 0.0304 | 0.04 (-0.06, 0.13) | 0.4468 |
| CRYBB2 | Beta-crystallin B2 | -0.0576 | -0.48 (-0.57, -0.38) | 2.10E-22 |
| CSF3R | Granulocyte colony-stimulating factor receptor | 0.0027 | 0.55 (0.46, 0.65) | 1.40E-30 |
| CST1 | Cystatin-SN | -0.0121 | -0.33 (-0.43, -0.23) | 2.50E-11 |
| CST5 | Cystatin-D | -0.089 | -0.33 (-0.42, -0.23) | 1.10E-11 |
| CST6 | Cystatin-M | -0.0831 | -0.17 (-0.26, -0.07) | 0.0006 |
| CSTB | Cystatin-B | 0.0409 | 0.95 (0.86, 1.04) | 6.00E-86 |
| CTBS | Di-N-acetylchitobiase | -0.002 | 0.68 (0.58, 0.77) | 2.10E-44 |
| CTHRC1 | Collagen triple helix repeat-containing protein 1 | 0.1693 | 1.2 (1.11, 1.29) | 7.00E-134 |
| CTNNA1 | Catenin alpha-1 | 0.0214 | -0.07 (-0.16, 0.03) | 0.1585 |
| CXCL13 | C-X-C motif chemokine 13 | -0.0335 | 0.51 (0.42, 0.61) | 7.70E-27 |
| CXCL17 | C-X-C motif chemokine 17 | -0.2006 | -0.15 (-0.25, -0.06) | 0.0017 |
| CXCL9 | C-X-C motif chemokine 9 | -0.0048 | 0.06 (-0.04, 0.15) | 0.2409 |
| CYTL1 | Cytokine-like protein 1 | -0.2013 | -0.14 (-0.24, -0.05) | 0.0034 |
| DCLRE1C | Protein artemis | -0.0125 | -0.12 (-0.21, -0.02) | 0.0136 |
| DCUN1D1 | DCN1-like protein 1 | 0.0431 | -0.07 (-0.17, 0.02) | 0.1175 |
| DEFB103A_DEFB103B | Beta-defensin 103 | 0.024 | 0.3 (0.2, 0.39) | 4.70E-10 |
| DEFB4A_DEFB4B | Beta-defensin 4A | 0.0502 | 0.6 (0.51, 0.7) | 1.70E-36 |
| DKK3 | Dickkopf-related protein 3 | -0.0759 | -0.84 (-0.93, -0.74) | 3.70E-69 |
| DLL1 | Delta-like protein 1 | 0.0581 | 0.69 (0.59, 0.79) | 1.40E-42 |

| | | | | |
|---|---|---|---|---|
| DMP1 | Dentin matrix acidic phosphoprotein 1 | -0.0295 | -0.66 (-0.75, -0.56) | 6.20E-42 |
| DPEP2 | Dipeptidase 2 | 0.0018 | 0.01 (-0.09, 0.1) | 0.9016 |
| DPT | Dermatopontin | 0.0935 | 1.35 (1.26, 1.44) | 1.40E-172 |
| DSG3 | Desmoglein-3 | 0.0637 | 0.32 (0.22, 0.41) | 4.60E-11 |
| DSG4 | Desmoglein-4 | -0.0339 | -0.46 (-0.55, -0.37) | 4.00E-22 |
| EDIL3 | EGF-like repeat and discoidin I-like domain-containing protein 3 | -0.0505 | -0.61 (-0.71, -0.52) | 2.00E-38 |
| EDN1 | Endothelin-1 | 0.0334 | 0.45 (0.35, 0.55) | 9.50E-19 |
| EFEMP1 | EGF-containing fibulin-like extracellular matrix protein 1 | 0.175 | 0.7 (0.6, 0.8) | 5.50E-45 |
| EGFLAM | Pikachurin | -0.014 | -0.27 (-0.36, -0.17) | 1.80E-08 |
| EIF4EBP1 | Eukaryotic translation initiation factor 4E-binding protein 1 | -0.036 | 0.39 (0.29, 0.48) | 4.70E-16 |
| ENG | Endoglin | -0.1619 | -0.58 (-0.68, -0.49) | 1.90E-34 |
| ENO3 | Beta-enolase | 0.0344 | 0.91 (0.81, 1) | 1.50E-83 |
| ENPP6 | Glycerophosphocholine cholinephosphodiesterase ENPP6 | -0.047 | -1.15 (-1.24, -1.07) | 1.40E-136 |
| ENTPD2 | Ectonucleoside triphosphate diphosphohydrolase 2 | -0.0153 | -0.02 (-0.12, 0.07) | 0.6209 |
| EPHA1 | Ephrin type-A receptor 1 | 0.0021 | 1.17 (1.07, 1.26) | 2.40E-126 |
| ERCC1 | DNA excision repair protein ERCC-1 | -0.0022 | -0.07 (-0.16, 0.03) | 0.1561 |
| ESM1 | Endothelial cell-specific molecule 1 | -0.0334 | -0.93 (-1.02, -0.83) | 8.70E-86 |
| EXTL1 | Exostosin-like 1 | -0.0501 | -0.26 (-0.35, -0.17) | 5.40E-08 |
| EZR | Ezrin | 0.0604 | 0.98 (0.89, 1.07) | 8.60E-98 |
| FABP4 | Fatty acid-binding protein, adipocyte | 0.34 | 2.15 (2.07, 2.24) | 0.00E+00 |
| FABP9 | Fatty acid-binding protein 9 | -0.0259 | -0.25 (-0.34, -0.16) | 1.80E-07 |
| FAM13A | Protein FAM13A | -0.0004 | 0.31 (0.22, 0.4) | 5.70E-11 |
| FAM3D | Protein FAM3D | -0.0405 | -0.28 (-0.37, -0.19) | 2.70E-09 |
| FAP | Prolyl endopeptidase FAP | -0.0165 | 0.53 (0.43, 0.62) | 1.10E-27 |
| FASLG | Tumor necrosis factor ligand superfamily member 6 | -0.0084 | -0.33 (-0.42, -0.24) | 5.90E-12 |
| FCER2 | Low affinity immunoglobulin epsilon Fc receptor | 0.0041 | 0.96 (0.86, 1.05) | 1.50E-83 |
| FCRL6 | Fc receptor-like protein 6 | -0.0033 | -0.06 (-0.15, 0.04) | 0.2368 |
| FDX1 | Adrenodoxin, mitochondrial | 0.0033 | 0.44 (0.35, 0.54) | 5.20E-21 |
| FGFBP1 | Fibroblast growth factor-binding protein 1 | 0.0174 | -0.14 (-0.23, -0.04) | 0.0053 |
| FGFBP3 | Fibroblast growth factor-binding protein 3 | 0.0206 | 0.45 (0.35, 0.54) | 9.10E-21 |
| FGL1 | Fibrinogen-like protein 1 | -0.0523 | -0.63 (-0.73, -0.54) | 3.00E-40 |
| FGR | Tyrosine-protein kinase Fgr | -0.0013 | 0.5 (0.41, 0.59) | 9.30E-27 |
| FLT4 | Vascular endothelial growth factor receptor 3 | 0.0268 | 0.9 (0.81, 1) | 5.30E-79 |
| FN1 | Fibronectin | 0.0546 | 1.1 (1.01, 1.19) | 1.10E-122 |
| FOLR1 | Folate receptor alpha | -0.1915 | -0.81 (-0.91, -0.72) | 1.90E-61 |
| FSHB | Follitropin subunit beta | -0.0035 | -0.38 (-0.48, -0.29) | 1.60E-15 |
| FSTL1 | Follistatin-related protein 1 | 0.0037 | 0.21 (0.12, 0.31) | 1.40E-05 |
| FUCA1 | Tissue alpha-L-fucosidase | -0.0118 | 0.27 (0.18, 0.37) | 2.00E-08 |
| FURIN | Furin | 0.0313 | 1.74 (1.65, 1.82) | 2.80E-298 |
| FUT3_FUT5 | 3-galactosyl-N-acetylglucosaminide 4-alpha-L-fucosyltransferase FUT3_4-galactosyl-N-acetylglucosaminide 3-alpha-L-fucosyltransferase FUT5 | 0.016 | 0.26 (0.17, 0.36) | 1.40E-07 |
| FZD10 | Frizzled-10 | -0.0128 | -0.07 (-0.16, 0.02) | 0.1392 |
| GABARAP | Gamma-aminobutyric acid receptor-associated protein | -0.0017 | 0 (-0.09, 0.09) | 0.9951 |

| | | | | |
|---|---|---|---|---|
| GAD1 | Glutamate decarboxylase 1 | -0.0063 | -0.18 (-0.27, -0.08) | 0.0002 |
| GAGE2A | G antigen 2A | -0.0055 | -0.07 (-0.17, 0.02) | 0.1178 |
| GAL | Galanin peptides | -0.064 | -0.59 (-0.68, -0.5) | 9.60E-36 |
| GALNT5 | Polypeptide N-acetylgalactosaminyltransferase 5 | -0.0011 | -0.21 (-0.31, -0.12) | 8.50E-06 |
| GASK1A | Golgi-associated kinase 1A | 0.0131 | 0.88 (0.79, 0.98) | 7.40E-77 |
| GC | Vitamin D-binding protein | -0.0229 | -0.09 (-0.18, 0.01) | 0.0712 |
| GDF2 | Growth/differentiation factor 2 | -0.0688 | -0.75 (-0.85, -0.66) | 4.50E-57 |
| GFRAL | GDNF family receptor alpha-like | 0.0511 | 0.39 (0.29, 0.48) | 7.70E-16 |
| GHR | Growth hormone receptor | 0.0172 | 1.61 (1.53, 1.7) | 1.00E-263 |
| GHRL | Appetite-regulating hormone | -0.0668 | -1.02 (-1.11, -0.93) | 6.90E-101 |
| GIP | Gastric inhibitory polypeptide | -0.027 | -0.12 (-0.22, -0.03) | 0.0136 |
| GOLM2 | Protein GOLM2 | 0.0425 | 1.09 (1, 1.18) | 4.50E-118 |
| GPD1 | Glycerol-3-phosphate dehydrogenase | 0.0752 | 1.67 (1.58, 1.75) | 4.70E-284 |
| GPHA2 | Glycoprotein hormone alpha-2 | -0.0352 | -0.56 (-0.65, -0.47) | 3.90E-33 |
| GPIHBP1 | Glycosylphosphatidylinositol-anchored high density lipoprotein-binding protein 1 | 0.0068 | 0.18 (0.08, 0.27) | 0.0002 |
| GPR158 | Probable G-protein coupled receptor 158 | -0.0359 | -0.75 (-0.84, -0.66) | 1.90E-55 |
| GRIN2B | Glutamate receptor ionotropic, NMDA 2B | 0.0054 | 0 (-0.1, 0.09) | 0.9562 |
| GRP | Gastrin-releasing peptide | 0.0116 | -0.1 (-0.19, 0) | 0.0428 |
| GUCA2A | Guanylin | -0.0039 | -0.71 (-0.81, -0.62) | 3.80E-48 |
| HDAC8 | Histone deacetylase 8 | -0.0244 | -0.09 (-0.18, 0.01) | 0.0681 |
| HDGF | Hepatoma-derived growth factor | -0.007 | 0.16 (0.07, 0.26) | 0.0005 |
| HEPACAM2 | HEPACAM family member 2 | -0.0012 | -0.48 (-0.58, -0.39) | 4.90E-25 |
| HGFAC | Hepatocyte growth factor activator | 0.0073 | 0.2 (0.1, 0.29) | 0.0001 |
| HJV | Hemojuvelin | 0.0092 | 1.07 (0.98, 1.16) | 4.60E-118 |
| HRAS | GTPase Hras | 0.0237 | 0.12 (0.03, 0.21) | 0.0117 |
| HRC | Sarcoplasmic reticulum histidine-rich calcium-binding protein | 0.0093 | -0.36 (-0.45, -0.26) | 8.90E-14 |
| HRG | Histidine-rich glycoprotein | 0.0231 | 0.33 (0.24, 0.43) | 2.50E-12 |
| HS3ST3B1 | Heparan sulfate glucosamine 3-O-sulfotransferase 3B1 | -0.018 | 0.01 (-0.08, 0.11) | 0.7916 |
| HS6ST2 | Heparan-sulfate 6-O-sulfotransferase 2 | -0.0455 | -0.27 (-0.37, -0.18) | 2.40E-08 |
| HSPG2 | Basement membrane-specific heparan sulfate proteoglycan core protein | 0.0257 | 1.43 (1.33, 1.53) | 2.20E-176 |
| ICAM5 | Intercellular adhesion molecule 5 | -0.0139 | -0.54 (-0.63, -0.45) | 2.10E-29 |
| ICOSLG | ICOS ligand | -0.0945 | -0.68 (-0.77, -0.59) | 8.40E-47 |
| ID4 | DNA-binding protein inhibitor ID-4 | -0.0122 | -0.14 (-0.24, -0.05) | 0.0019 |
| IFIT3 | Interferon-induced protein with tetratricopeptide repeats 3 | -0.0413 | 0.1 (0, 0.19) | 0.039 |
| IFNL2 | Interferon lambda-2 | 0.0199 | 0.02 (-0.07, 0.12) | 0.6022 |
| IGFBP1 | Insulin-like growth factor-binding protein 1 | -0.047 | -1.48 (-1.56, -1.39) | 1.30E-224 |
| IGSF3 | Immunoglobulin superfamily member 3 | 0.0366 | 1.19 (1.1, 1.29) | 4.20E-128 |
| IL17C | Interleukin-17C | 0.0653 | 0.26 (0.16, 0.35) | 1.60E-07 |
| IL1B | Interleukin-1 beta | -0.0231 | 0.27 (0.17, 0.36) | 3.10E-08 |
| IL1R1 | Interleukin-1 receptor type 1 | -0.1056 | -0.43 (-0.52, -0.33) | 1.20E-17 |
| IL1RN | Interleukin-1 receptor antagonist protein | 0.0168 | 1.79 (1.71, 1.88) | 1.1e-319 |
| IL21R | Interleukin-21 receptor | 0.0241 | 0.04 (-0.05, 0.14) | 0.3456 |

| | | | | |
|---|---|---|---|---|
| IL32 | Interleukin-32 | -0.0189 | -0.53 (-0.63, -0.44) | 1.10E-27 |
| IL33 | Interleukin-33 | -0.0189 | -0.03 (-0.12, 0.07) | 0.5461 |
| IL36G | Interleukin-36 gamma | 0.0026 | 0.35 (0.26, 0.45) | 1.60E-13 |
| INSL3 | Insulin-like 3 | 0.0592 | 0.35 (0.26, 0.44) | 3.90E-14 |
| INSL4 | Early placenta insulin-like peptide | 0.0358 | 0.2 (0.11, 0.29) | 1.70E-05 |
| INSL5 | Insulin-like peptide INSL5 | 0.0099 | 0.56 (0.47, 0.65) | 1.80E-32 |
| ISLR2 | Immunoglobulin superfamily containing leucine-rich repeat protein 2 | 0.0204 | -0.14 (-0.23, -0.05) | 0.0026 |
| ITGAV | Integrin alpha-V | -0.007 | -0.54 (-0.63, -0.45) | 6.50E-30 |
| ITGB5 | Integrin beta-5 | -0.0066 | -0.13 (-0.23, -0.04) | 0.0044 |
| ITIH4 | Inter-alpha-trypsin inhibitor heavy chain H4 | 0.0242 | 0.65 (0.56, 0.74) | 2.90E-43 |
| IVD | Isovaleryl-CoA dehydrogenase, mitochondrial | 1.70E-05 | 0.19 (0.1, 0.28) | 4.70E-05 |
| KIAA0319 | Dyslexia-associated protein KIAA0319 | -0.0104 | -0.89 (-0.98, -0.8) | 1.60E-77 |
| KIRREL2 | Kin of IRRE-like protein 2 | -0.0063 | -0.49 (-0.58, -0.39) | 8.70E-25 |
| KLK11 | Kallikrein-11 | -0.0437 | -0.2 (-0.29, -0.1) | 0.0001 |
| KLK12 | Kallikrein-12 | 0.0004 | 0.01 (-0.08, 0.1) | 0.816 |
| KLK14 | Kallikrein-14 | -0.0468 | -0.41 (-0.5, -0.31) | 2.30E-17 |
| KLK15 | Kallikrein-15 | -0.0122 | -0.13 (-0.22, -0.03) | 0.0087 |
| KLK4 | Kallikrein-4 | -0.021 | -0.34 (-0.43, -0.25) | 3.90E-13 |
| KRT8 | Keratin, type II cytoskeletal 8 | 0.0027 | 0.23 (0.13, 0.33) | 5.90E-06 |
| L1CAM | Neural cell adhesion molecule L1 | 0.0508 | 0.16 (0.07, 0.26) | 0.0006 |
| LACRT | Extracellular glycoprotein lacritin | -0.0479 | -0.61 (-0.7, -0.52) | 7.60E-39 |
| LAMB1 | Laminin subunit beta-1 | 0.0166 | 0 (-0.1, 0.09) | 0.9418 |
| LAMP3 | Lysosome-associated membrane glycoprotein 3 | 0.0639 | 0.38 (0.28, 0.47) | 9.50E-15 |
| LAYN | Layilin | -0.0479 | -0.35 (-0.45, -0.25) | 4.50E-12 |
| LDLR | Low-density lipoprotein receptor | 0.0189 | 1.44 (1.36, 1.53) | 2.20E-218 |
| LEP | Leptin | 1.0981 | 2.3 (2.22, 2.38) | 0.00E+00 |
| LEPR | Leptin receptor | -0.0112 | -1.16 (-1.25, -1.07) | 1.10E-134 |
| LGALS3 | Galectin-3 | 0.0303 | 0.58 (0.48, 0.67) | 1.30E-31 |
| LGALS3BP | Galectin-3-binding protein | 0.05 | 1.15 (1.06, 1.24) | 2.80E-128 |
| LILRA5 | Leukocyte immunoglobulin-like receptor subfamily A member 5 | 0.0333 | 1.27 (1.17, 1.36) | 7.30E-149 |
| LMNB2 | Lamin-B2 | 0.0243 | 1.07 (0.98, 1.17) | 8.90E-106 |
| LMOD1 | Leiomodin-1 | -0.034 | -0.07 (-0.17, 0.02) | 0.144 |
| LRPAP1 | Alpha-2-macroglobulin receptor-associated protein | 0.0341 | 0.65 (0.56, 0.74) | 1.30E-45 |
| LRRC38 | Leucine-rich repeat-containing protein 38 | -0.002 | 0.15 (0.06, 0.24) | 0.0016 |
| LRRN1 | Leucine-rich repeat neuronal protein 1 | -0.0685 | -0.24 (-0.33, -0.15) | 4.60E-07 |
| LRTM2 | Leucine-rich repeat and transmembrane domain-containing protein 2 | -0.0069 | -1.09 (-1.18, -0.99) | 2.50E-115 |
| LTBP2 | Latent-transforming growth factor beta-binding protein 2 | -0.0565 | -0.68 (-0.77, -0.58) | 1.70E-44 |
| LTBP3 | Latent-transforming growth factor beta-binding protein 3 | 0.0182 | 0.84 (0.74, 0.93) | 3.30E-68 |
| MBL2 | Mannose-binding protein C | -0.0223 | -0.2 (-0.29, -0.11) | 3.20E-05 |
| MEGF10 | Multiple epidermal growth factor-like domains protein 10 | 0.0028 | -0.19 (-0.29, -0.1) | 3.00E-05 |
| MELTF | Melanotransferrin | 0.0032 | -0.14 (-0.23, -0.04) | 0.0037 |
| MEPE | Matrix extracellular phosphoglycoprotein | -0.058 | 0.28 (0.18, 0.37) | 5.90E-09 |
| MERTK | Tyrosine-protein kinase Mer | 0.036 | 0.37 (0.28, 0.47) | 1.10E-14 |

| | | | | |
|---|---|---|---|---|
| MFGE8 | Lactadherin | -0.0389 | 0.24 (0.15, 0.34) | 7.20E-07 |
| MIA | Melanoma-derived growth regulatory protein | -0.1294 | -0.13 (-0.23, -0.04) | 0.0059 |
| MMP12 | Macrophage metalloelastase | -0.0652 | 0 (-0.1, 0.1) | 0.9445 |
| MNDA | Myeloid cell nuclear differentiation antigen | -0.0285 | 0.33 (0.24, 0.42) | 2.30E-12 |
| MOG | Myelin-oligodendrocyte glycoprotein | -0.1972 | -0.8 (-0.89, -0.71) | 2.80E-61 |
| MRC1 | Macrophage mannose receptor 1 | 0.06 | 1.09 (1, 1.18) | 3.40E-112 |
| MSTN | Growth/differentiation factor 8 | 0.0272 | 0.45 (0.36, 0.55) | 4.30E-21 |
| MUC16 | Mucin-16 | 0.0074 | 0.08 (-0.01, 0.17) | 0.0878 |
| MYBPC2 | Myosin-binding protein C, fast-type | 0.0041 | 0.34 (0.24, 0.43) | 1.60E-12 |
| MYL3 | Myosin light chain 3 | 0.0691 | 0.55 (0.46, 0.65) | 3.70E-30 |
| MYOM3 | Myomesin-3 | 0.0859 | 0.34 (0.24, 0.43) | 3.10E-12 |
| NADK | NAD kinase | -0.0429 | 0.71 (0.62, 0.8) | 7.80E-52 |
| NAGPA | N-acetylglucosamine-1-phosphodiester alpha-N-acetylglucosaminidase | 0.0078 | 0.57 (0.48, 0.67) | 1.90E-31 |
| NAP1L4 | Nucleosome assembly protein 1-like 4 | -0.0256 | 0.07 (-0.03, 0.16) | 0.1607 |
| NCAM2 | Neural cell adhesion molecule 2 | -0.0577 | -1.06 (-1.15, -0.97) | 2.60E-112 |
| NCAN | Neurocan core protein | -0.1099 | -1.09 (-1.19, -1) | 1.00E-115 |
| NEFL | Neurofilament light polypeptide | -0.1485 | -0.52 (-0.61, -0.42) | 1.50E-24 |
| NELL1 | Protein kinase C-binding protein NELL1 | 0.063 | -0.18 (-0.27, -0.08) | 0.0004 |
| NFATC3 | Nuclear factor of activated T-cells, cytoplasmic 3 | -0.0367 | 0.28 (0.19, 0.38) | 7.50E-09 |
| NHLRC3 | NHL repeat-containing protein 3 | 0.0242 | 1.02 (0.92, 1.12) | 1.80E-90 |
| NPHS2 | Podocin | 0.0025 | 0.06 (-0.04, 0.15) | 0.2247 |
| NPPC | C-type natriuretic peptide | -0.0571 | 0.53 (0.44, 0.63) | 4.80E-27 |
| NPTX1 | Neuronal pentraxin-1 | -0.0416 | -0.56 (-0.65, -0.46) | 5.30E-31 |
| NPTX2 | Neuronal pentraxin-2 | -0.0024 | -0.2 (-0.29, -0.1) | 3.40E-05 |
| NRCAM | Neuronal cell adhesion molecule | 0.0104 | 0.33 (0.23, 0.42) | 2.90E-11 |
| NTRK2 | BDNF/NT-3 growth factors receptor | 0.1347 | 0.57 (0.48, 0.66) | 2.50E-32 |
| NTRK3 | NT-3 growth factor receptor | -0.0245 | -0.99 (-1.08, -0.9) | 2.00E-97 |
| NXPE4 | NXPE family member 4 | 0.0067 | 0 (-0.09, 0.09) | 0.9967 |
| ODAM | Odontogenic ameloblast-associated protein | -0.0087 | -0.33 (-0.42, -0.24) | 3.90E-12 |
| OGT | UDP-N-acetylglucosamine--peptide N-acetylglucosaminyltransferase 110 kDa subunit | -0.0048 | -0.09 (-0.18, 0) | 0.0573 |
| OMG | Oligodendrocyte-myelin glycoprotein | 0.2207 | -0.49 (-0.58, -0.39) | 5.20E-24 |
| OPTC | Opticin | -0.2481 | -0.91 (-1, -0.82) | 2.00E-81 |
| PAEP | Glycodelin | -0.0462 | -0.14 (-0.22, -0.05) | 0.0016 |
| PALM | Paralemmin-1 | 0.1206 | 1.47 (1.38, 1.56) | 2.40E-202 |
| PAM | Peptidyl-glycine alpha-amidating monooxygenase | 0.0642 | 0.46 (0.36, 0.55) | 1.80E-20 |
| PAMR1 | Inactive serine protease PAMR1 | 0.0458 | 1.32 (1.23, 1.41) | 1.30E-170 |
| PCDH12 | Protocadherin-12 | -0.0382 | -0.01 (-0.1, 0.08) | 0.8189 |
| PCDH17 | Protocadherin-17 | -0.0321 | 0.02 (-0.08, 0.11) | 0.7218 |
| PDCD6 | Programmed cell death protein 6 | -0.0052 | -0.05 (-0.14, 0.05) | 0.3432 |
| PENK | Proenkephalin-A | -0.0239 | 0.15 (0.06, 0.25) | 0.0022 |
| PFDN6 | Prefoldin subunit 6 | 0.0054 | 0.01 (-0.08, 0.1) | 0.8279 |
| PINLYP | phospholipase A2 inhibitor and Ly6/PLAUR domain-containing protein | -0.0241 | 0.1 (0.01, 0.19) | 0.0373 |

| | | | | |
|---|---|---|---|---|
| PLA2G1B | Phospholipase A2 | -0.0326 | -0.98 (-1.07, -0.89) | 1.20E-90 |
| PLA2G2A | Phospholipase A2, membrane associated | 0.022 | 0.49 (0.39, 0.58) | 4.60E-25 |
| PM20D1 | N-fatty-acyl-amino acid synthase/hydrolase PM20D1 | -0.0189 | -0.02 (-0.11, 0.08) | 0.7378 |
| PODXL2 | Podocalyxin-like protein 2 | -0.0154 | -0.64 (-0.74, -0.55) | 7.00E-40 |
| POMC | Pro-opiomelanocortin | 0.0494 | 0.56 (0.47, 0.65) | 1.50E-31 |
| PRDX1 | Peroxiredoxin-1 | -0.0055 | -0.06 (-0.15, 0.04) | 0.2474 |
| PRRT3 | Proline-rich transmembrane protein 3 | -0.2645 | -0.91 (-1, -0.81) | 5.90E-78 |
| PRSS2 | Trypsin-2 | -0.1027 | -0.16 (-0.26, -0.07) | 0.001 |
| PRSS27 | Serine protease 27 | -0.0432 | -0.24 (-0.33, -0.14) | 6.70E-07 |
| PRSS53 | Serine protease 53 | -0.0063 | -0.05 (-0.14, 0.05) | 0.3269 |
| PSPN | Persephin | 0.1437 | 0.78 (0.69, 0.88) | 9.70E-57 |
| PTPRR | Receptor-type tyrosine-protein phosphatase R | -0.0494 | -0.68 (-0.78, -0.59) | 2.70E-45 |
| PTPRS | Receptor-type tyrosine-protein phosphatase S | 0.0105 | 0.27 (0.17, 0.36) | 4.20E-08 |
| PYDC1 | Pyrin domain-containing protein 1 | 0.0137 | 0.25 (0.15, 0.34) | 2.30E-07 |
| REG1A | Lithostathine-1-alpha | -0.0299 | -0.47 (-0.56, -0.37) | 4.80E-22 |
| REG3A | Regenerating islet-derived protein 3-alpha | -0.0381 | -0.38 (-0.47, -0.28) | 3.20E-15 |
| RNASE1 | Ribonuclease pancreatic | 0.0347 | 1.23 (1.13, 1.32) | 1.00E-130 |
| ROBO1 | Roundabout homolog 1 | 0.0663 | 0.48 (0.39, 0.58) | 1.90E-22 |
| ROBO2 | Roundabout homolog 2 | 0.0192 | 0.02 (-0.08, 0.12) | 0.6877 |
| RPL14 | 60S ribosomal protein L14 | 0.0291 | 0.66 (0.56, 0.75) | 7.00E-45 |
| RSPO1 | R-spondin-1 | -0.0048 | -0.23 (-0.33, -0.14) | 1.20E-06 |
| RSPO3 | R-spondin-3 | -0.077 | 0.02 (-0.08, 0.11) | 0.7475 |
| RTN4R | Reticulon-4 receptor | 0.0365 | 1.57 (1.48, 1.66) | 1.50E-231 |
| S100G | Protein S100-G | 0.0184 | 0.01 (-0.08, 0.11) | 0.7705 |
| SCARF2 | Scavenger receptor class F member 2 | -0.1717 | -0.52 (-0.61, -0.42) | 8.40E-27 |
| SCG2 | Secretogranin-2 | -0.0931 | -0.51 (-0.61, -0.42) | 2.30E-25 |
| SCGB1A1 | Uteroglobin | -0.01 | -0.37 (-0.46, -0.27) | 8.20E-14 |
| SCGB3A1 | Secretoglobin family 3A member 1 | -0.0528 | -0.78 (-0.87, -0.69) | 2.70E-60 |
| SCGB3A2 | Secretoglobin family 3A member 2 | -0.073 | -1.22 (-1.31, -1.13) | 1.80E-146 |
| SCN4B | Sodium channel subunit beta-4 | 0.004 | 0.56 (0.47, 0.66) | 8.60E-31 |
| SCPEP1 | Retinoid-inducible serine carboxypeptidase | 0.0342 | 0.63 (0.54, 0.73) | 8.40E-41 |
| SELE | E-selectin | 0.0638 | 1.24 (1.14, 1.33) | 2.90E-141 |
| SEPTIN3 | Neuronal-specific septin-3 | 0.0045 | 0.19 (0.1, 0.29) | 0.0001 |
| SERPINA4 | Kallistatin | -0.0471 | 0.31 (0.21, 0.4) | 2.10E-10 |
| SERPINA7 | Thyroxine-binding globulin | 0.01 | 0.59 (0.49, 0.68) | 1.30E-34 |
| SERPINC1 | Antithrombin-III | -0.0688 | -0.36 (-0.45, -0.27) | 4.70E-14 |
| SETMAR | Histone-lysine N-methyltransferase SETMAR | 0.0049 | 1.29 (1.21, 1.38) | 8.70E-170 |
| SEZ6 | Seizure protein 6 homolog | -0.0486 | -0.81 (-0.91, -0.72) | 1.40E-65 |
| SEZ6L | Seizure 6-like protein | -0.0715 | -1.04 (-1.13, -0.95) | 3.80E-105 |
| SEZ6L2 | Seizure 6-like protein 2 | 0.0306 | -0.53 (-0.62, -0.44) | 7.10E-30 |
| SFRP1 | Secreted frizzled-related protein 1 | 0.0856 | 0.9 (0.8, 0.99) | 6.10E-75 |
| SFRP4 | Secreted frizzled-related protein 4 | -0.0213 | 0.72 (0.63, 0.81) | 2.90E-54 |
| SFTPD | Pulmonary surfactant-associated protein D | -0.0953 | -0.36 (-0.45, -0.26) | 3.00E-13 |

| | | | | |
|---|---|---|---|---|
| SHBG | Sex hormone-binding globulin | -0.0832 | -1.45 (-1.53, -1.36) | 2.30E-229 |
| SIGLEC7 | Sialic acid-binding Ig-like lectin 7 | 0.0119 | 0.76 (0.66, 0.86) | 3.90E-53 |
| SIL1 | Nucleotide exchange factor SIL1 | 0.003 | 0.46 (0.37, 0.55) | 9.40E-23 |
| SLITRK1 | SLIT and NTRK-like protein 1 | -0.142 | -1.16 (-1.25, -1.07) | 7.50E-134 |
| SLITRK2 | SLIT and NTRK-like protein 2 | 0.0182 | 0.67 (0.58, 0.77) | 2.00E-43 |
| SMAD5 | Mothers against decapentaplegic homolog 5 | -0.0039 | 0.77 (0.68, 0.87) | 1.20E-56 |
| SNCG | Gamma-synuclein | 0.0603 | 1.21 (1.11, 1.3) | 1.20E-128 |
| SORCS2 | VPS10 domain-containing receptor SorCS2 | -0.0298 | 0.12 (0.02, 0.21) | 0.0233 |
| SPACA5_SPACA5B | Sperm acrosome-associated protein 5 | 0.0143 | 0 (-0.09, 0.1) | 0.9522 |
| SPINK2 | Serine protease inhibitor Kazal-type 2 | 0.0352 | 0.66 (0.57, 0.75) | 1.30E-42 |
| SPINK6 | Serine protease inhibitor Kazal-type 6 | 0.006 | 0.79 (0.7, 0.88) | 3.90E-62 |
| SPON2 | Spondin-2 | -0.0028 | 0.54 (0.44, 0.64) | 9.90E-27 |
| SPP1 | Osteopontin | -0.016 | -0.35 (-0.45, -0.26) | 5.50E-13 |
| SSC4D | Scavenger receptor cysteine-rich domain-containing group B protein | 0.0317 | 1.75 (1.66, 1.83) | 5.9e-312 |
| STX1B | Syntaxin-1B | -0.0181 | -0.26 (-0.35, -0.17) | 5.10E-08 |
| SUSD2 | Sushi domain-containing protein 2 | -0.0528 | -0.82 (-0.91, -0.73) | 1.40E-65 |
| SUSD4 | Sushi domain-containing protein 4 | 0.0076 | -0.15 (-0.25, -0.06) | 0.0014 |
| SYT1 | Synaptotagmin-1 | -0.0775 | -0.63 (-0.72, -0.53) | 3.90E-39 |
| TAGLN3 | Transgelin-3 | -0.011 | -0.08 (-0.18, 0.01) | 0.0777 |
| TARM1 | T-cell-interacting, activating receptor on myeloid cells protein 1 | -0.0008 | -0.13 (-0.22, -0.03) | 0.0071 |
| TCTN3 | Tectonic-3 | 0.0031 | 0.92 (0.83, 1.02) | 2.30E-81 |
| TFPI | Tissue factor pathway inhibitor | 0.0085 | 0.72 (0.62, 0.81) | 4.80E-52 |
| TGFBR2 | TGF-beta receptor type-2 | 0.2394 | 1.17 (1.08, 1.27) | 3.90E-122 |
| THOP1 | Thimet oligopeptidase | -0.0118 | 0.85 (0.76, 0.94) | 1.60E-70 |
| THRAP3 | Thyroid hormone receptor-associated protein 3 | 0.0349 | 0.05 (-0.04, 0.14) | 0.2673 |
| THY1 | Thy-1 membrane glycoprotein | 0.277 | 1.1 (1, 1.19) | 2.80E-109 |
| TIMD4 | T-cell immunoglobulin and mucin domain-containing protein 4 | 0.0424 | 0.74 (0.65, 0.84) | 3.50E-53 |
| TMPRSS11D | Transmembrane protease serine 11D | -0.0891 | -0.18 (-0.28, -0.09) | 0.0002 |
| TMPRSS5 | Transmembrane protease serine 5 | -0.0439 | -0.74 (-0.84, -0.65) | 2.10E-55 |
| TNC | Tenascin | -0.0022 | -0.12 (-0.22, -0.03) | 0.0112 |
| TNFAIP8 | Tumor necrosis factor alpha-induced protein 8 | -0.0012 | 0.07 (-0.02, 0.16) | 0.1399 |
| TNFRSF21 | Tumor necrosis factor receptor superfamily member 21 | 0.0103 | 0.24 (0.15, 0.34) | 8.00E-07 |
| TNFSF10 | Tumor necrosis factor ligand superfamily member 10 | 0.065 | 0.78 (0.69, 0.87) | 4.50E-61 |
| TNNI3 | Troponin I, cardiac muscle | 0.0155 | 0.14 (0.05, 0.24) | 0.0028 |
| TPK1 | Thiamin pyrophosphokinase 1 | -0.0071 | 0.24 (0.14, 0.33) | 8.10E-07 |
| TPSD1 | Tryptase delta | 0.006 | 0.1 (0.01, 0.19) | 0.0361 |
| TRAF3 | TNF receptor-associated factor 3, Isoform 2 | 0.0104 | 0.29 (0.2, 0.38) | 5.60E-10 |
| TREH | Trehalase | -0.0089 | 0.55 (0.45, 0.64) | 2.20E-30 |
| TRIM25 | E3 ubiquitin/ISG15 ligase TRIM25 | -0.0222 | 0.21 (0.12, 0.3) | 9.30E-06 |
| TSC22D1 | TSC22 domain family protein 1 | 0.0105 | 0.28 (0.19, 0.37) | 1.30E-09 |
| TSHB | Thyrotropin subunit beta | 0.0066 | 0.2 (0.11, 0.3) | 1.80E-05 |
| TYRP1 | 5,6-dihydroxyindole-2-carboxylic acid oxidase | -0.0143 | -0.07 (-0.16, 0.03) | 0.1608 |

| | | | | |
|---|---|---|---|---|
| UMOD | Uromodulin | -0.0261 | -0.74 (-0.83, -0.65) | 1.20E-56 |
| VASN | Vasorin | 0.0226 | 0.56 (0.47, 0.66) | 2.40E-31 |
| VAT1 | Synaptic vesicle membrane protein VAT-1 homolog | 0.0211 | 0.34 (0.25, 0.44) | 4.80E-13 |
| VEGFD | Vascular endothelial growth factor D | -0.0107 | -0.99 (-1.08, -0.9) | 1.30E-95 |
| VSIG2 | V-set and immunoglobulin domain-containing protein 2 | -0.0188 | -0.29 (-0.39, -0.19) | 1.30E-08 |
| VWA1 | von Willebrand factor A domain-containing protein 1 | -0.0115 | 0.89 (0.8, 0.98) | 6.60E-78 |
| VWC2 | Brorin | 0.0129 | 0.59 (0.5, 0.69) | 5.20E-33 |
| VWC2L | von Willebrand factor C domain-containing protein 2-like | -0.1231 | -1.03 (-1.13, -0.94) | 4.90E-100 |
| WFDC12 | WAP four-disulfide core domain protein 12 | 0.0288 | 0.72 (0.63, 0.82) | 2.20E-51 |
| WFIKKN1 | WAP, Kazal, immunoglobulin, Kunitz and NTR domain-containing protein 1 | 0.0608 | 0.74 (0.64, 0.83) | 2.70E-53 |
| WFIKKN2 | WAP, Kazal, immunoglobulin, Kunitz and NTR domain-containing protein 2 | -0.0492 | -1.22 (-1.31, -1.13) | 1.70E-144 |
| WNT9A | Protein Wnt-9a | -0.028 | -0.45 (-0.55, -0.35) | 4.20E-19 |

BMI, body mass index; CI: confidence interval.
*Linear regressions of the measured BMI regressed on each of the proteins (per standard deviation).

**Supplemental Table 2B. Association between the 385 LASSO Selected Proteins and Body Fat Percentage in the Training Set of the Healthy Cohort.**

| Protein | Name | LASSO | Linear Regression* | |
|---|---|---|---|---|
| | | Beta | Beta (95% CI) | P |
| ACAN | Aggrecan core protein | -0.0781 | -1.53 (-1.71, -1.34) | 4.70E-57 |
| ACHE | Acetylcholinesterase | -0.0112 | -2.21 (-2.4, -2.03) | 1.90E-118 |
| ACTN4 | Alpha-actinin-4 | 0.0047 | 0.16 (-0.03, 0.34) | 0.1019 |
| ADAM12 | Disintegrin and metalloproteinase domain-containing protein 12 | 0.0455 | 2.19 (2.01, 2.38) | 7.60E-115 |
| ADAMTS15 | A disintegrin and metalloproteinase with thrombospondin motifs 15 | 0.2694 | 2.71 (2.52, 2.9) | 9.40E-168 |
| ADAMTS16 | A disintegrin and metalloproteinase with thrombospondin motifs 16 | 0.0188 | 1.87 (1.69, 2.06) | 5.10E-85 |
| ADAMTSL2 | ADAMTS-like protein 2 | 0.0309 | 1.89 (1.7, 2.08) | 1.50E-79 |
| ADAMTSL4 | ADAMTS-like protein 4 | -0.052 | 0.97 (0.79, 1.16) | 4.40E-24 |
| ADGRD1 | Adhesion G-protein coupled receptor D1 | -0.0535 | -1.36 (-1.55, -1.17) | 9.90E-45 |
| ADGRG2 | Adhesion G-protein coupled receptor G2 | -0.0482 | 0.69 (0.5, 0.88) | 5.40E-13 |
| ADM | Pro-adrenomedullin | 0.3844 | 3.05 (2.86, 3.24) | 1.30E-205 |
| AGER | Advanced glycosylation end product-specific receptor | -0.0398 | -0.24 (-0.43, -0.05) | 0.0136 |
| AGRP | Agouti-related protein | -0.0443 | -1.03 (-1.22, -0.85) | 1.10E-28 |
| AGXT | Serine--pyruvate aminotransferase | -0.0232 | 0.02 (-0.17, 0.21) | 0.8375 |
| AHNAK2 | Protein AHNAK2 | -0.0242 | -0.18 (-0.36, 0.01) | 0.0602 |
| AKR1C4 | Aldo-keto reductase family 1 member C4 | 0.008 | 0.22 (0.03, 0.4) | 0.0238 |
| AMOT | Angiomotin | 0.1023 | 2.96 (2.79, 3.14) | 1.30E-226 |
| ANGPTL2 | Angiopoietin-related protein 2 | 0.0225 | 1.2 (1.01, 1.39) | 1.00E-34 |
| ANGPTL7 | Angiopoietin-related protein 7 | -0.3151 | -1.24 (-1.43, -1.05) | 1.30E-37 |
| APLP1 | Amyloid-like protein 1 | -0.0628 | -0.28 (-0.47, -0.09) | 0.0039 |
| APOA4 | Apolipoprotein A-IV | -0.2446 | -0.87 (-1.06, -0.68) | 8.50E-20 |
| APOL1 | Apolipoprotein L1 | 0.0405 | 1.37 (1.19, 1.56) | 1.90E-47 |
| ARG2 | Arginase-2, mitochondrial | 0.0504 | 0.57 (0.39, 0.75) | 6.30E-10 |
| ARHGAP30 | Rho GTPase-activating protein 30 | -0.0098 | -0.2 (-0.39, -0.02) | 0.0292 |
| ART3 | Ecto-ADP-ribosyltransferase 3 | -0.178 | -3.38 (-3.55, -3.2) | 2.00E-289 |
| ASPN | Asporin | 0.0793 | 1.22 (1.04, 1.4) | 4.30E-38 |
| ATP1B1 | Sodium/potassium-transporting ATPase subunit beta-1 | -0.0262 | -0.39 (-0.57, -0.21) | 3.30E-05 |
| ATP6V1G2 | V-type proton ATPase subunit G 2 | 0.0088 | 0.06 (-0.12, 0.25) | 0.4961 |
| B4GALT1 | Beta-1,4-galactosyltransferase 1 | -0.048 | -0.08 (-0.28, 0.11) | 0.3957 |
| BAG3 | BAG family molecular chaperone regulator 3 | -0.0041 | 0.49 (0.3, 0.68) | 5.60E-07 |
| BCAT2 | Branched-chain-amino-acid aminotransferase, mitochondrial | 0.0099 | 0.25 (0.06, 0.43) | 0.01 |
| BGLAP | Osteocalcin | 0.0793 | 1.04 (0.85, 1.23) | 7.80E-27 |
| BLNK | B-cell linker protein | 0.018 | 0.57 (0.39, 0.76) | 1.20E-09 |
| BMP6 | Bone morphogenetic protein 6 | 0.0159 | -0.12 (-0.31, 0.07) | 0.1999 |
| BPIFB2 | BPI fold-containing family B member 2 | 0.0643 | 3.3 (3.13, 3.48) | 2.70E-269 |
| BRME1 | Break repair meiotic recombinase recruitment factor 1 | 0.0073 | 0.14 (-0.05, 0.32) | 0.1405 |
| C7 | Complement component C7 | -0.0507 | -0.12 (-0.31, 0.08) | 0.2393 |
| CA14 | Carbonic anhydrase 14 | -0.1317 | -3.61 (-3.79, -3.44) | 0.00E+00 |
| CALB2 | Calretinin | 0.1153 | 3 (2.82, 3.18) | 5.50E-225 |

| | | | | |
|---|---|---|---|---|
| CAPS | Calcyphosin | 0.0384 | 1.03 (0.85, 1.21) | 9.40E-29 |
| CARHSP1 | Calcium-regulated heat-stable protein 1 | 0.0224 | -0.6 (-0.79, -0.42) | 1.10E-10 |
| CBLN4 | Cerebellin-4 | -0.019 | 0.32 (0.13, 0.51) | 0.0009 |
| CCL15 | C-C motif chemokine 15 | -0.0306 | 0.16 (-0.03, 0.34) | 0.095 |
| CCL16 | C-C motif chemokine 16 | -0.0176 | 0.01 (-0.18, 0.21) | 0.8845 |
| CCL20 | C-C motif chemokine 20 | 0.0073 | 1.27 (1.09, 1.45) | 1.40E-42 |
| CCL27 | C-C motif chemokine 27 | -0.1968 | 0.08 (-0.11, 0.27) | 0.4258 |
| CCL7 | C-C motif chemokine 7 | 0.065 | 1.81 (1.63, 1.99) | 1.30E-81 |
| CCNE1 | G1/S-specific cyclin-E1 | -0.0105 | 0.08 (-0.1, 0.27) | 0.3643 |
| CD160 | CD160 antigen | 0.0115 | 0.75 (0.56, 0.94) | 1.40E-14 |
| CD1C | T-cell surface glycoprotein CD1c | -0.0201 | 0.08 (-0.11, 0.26) | 0.4219 |
| CD22 | B-cell receptor CD22 | 0.152 | 2.68 (2.5, 2.86) | 2.20E-173 |
| CD276 | CD276 antigen | -0.0617 | 0.43 (0.23, 0.62) | 2.10E-05 |
| CD300LG | CMRF35-like molecule 9 | 0.1262 | 1.46 (1.27, 1.65) | 2.30E-51 |
| CD38 | ADP-ribosyl cyclase/cyclic ADP-ribose hydrolase 1 | -0.3636 | -2.67 (-2.85, -2.49) | 1.20E-169 |
| CD70 | CD70 antigen | 0.031 | 1.96 (1.77, 2.14) | 1.80E-94 |
| CD83 | CD83 antigen | 0.0379 | 1.81 (1.62, 2) | 7.00E-75 |
| CD86 | T-lymphocyte activation antigen CD86 | 0.0033 | 0.86 (0.68, 1.05) | 1.80E-19 |
| CD99 | CD99 antigen | -0.1061 | -2 (-2.19, -1.82) | 4.90E-97 |
| CD99L2 | CD99 antigen-like protein 2 | 0.0451 | 1.9 (1.72, 2.09) | 2.30E-87 |
| CDH2 | Cadherin-2 | -0.0524 | 0.24 (0.04, 0.44) | 0.0207 |
| CDH5 | Cadherin-5 | -0.0121 | -0.42 (-0.61, -0.24) | 7.90E-06 |
| CDHR1 | Cadherin-related family member 1 | 0.0573 | 0.71 (0.53, 0.9) | 7.40E-14 |
| CDHR5 | Cadherin-related family member 5 | -0.0041 | 1.73 (1.55, 1.92) | 1.70E-73 |
| CEACAM21 | Carcinoembryonic antigen-related cell adhesion molecule 21 | 0.0075 | 0.33 (0.14, 0.51) | 0.0005 |
| CEP164 | Centrosomal protein of 164 kDa | 0.0147 | 0.33 (0.14, 0.52) | 0.0006 |
| CFB | Complement factor B | 0.1215 | 2.95 (2.77, 3.12) | 2.40E-217 |
| CFH | Complement factor H | 0.0384 | 2.4 (2.22, 2.59) | 2.30E-132 |
| CFHR2 | Complement factor H-related protein 2 | 0.0188 | 1.09 (0.9, 1.28) | 1.60E-28 |
| CFI | Complement factor I | 0.0412 | 3.14 (2.96, 3.31) | 3.50E-253 |
| CGB3_CGB5_CGB8 | Choriogonadotropin subunit beta 3 | 0.0754 | 1.75 (1.56, 1.93) | 2.80E-76 |
| CGREF1 | Cell growth regulator with EF hand domain protein 1 | -0.0003 | 0.59 (0.39, 0.78) | 2.10E-09 |
| CHGB | Secretogranin-1 | -0.0457 | -0.98 (-1.17, -0.79) | 2.90E-23 |
| CHRDL1 | Chordin-like protein 1 | 0.054 | 2.1 (1.91, 2.29) | 1.90E-101 |
| CHRDL2 | Chordin-like protein 2 | -0.0585 | -0.19 (-0.37, -0.01) | 0.0392 |
| CLC | Galectin-10 | -0.0207 | 0.12 (-0.06, 0.3) | 0.1907 |
| CLEC4A | C-type lectin domain family 4 member A | 0.0228 | 0.03 (-0.15, 0.21) | 0.7517 |
| CLMP | CXADR-like membrane protein | 0.9422 | 4.52 (4.35, 4.68) | 0.00E+00 |
| CLSTN2 | Calsyntenin-2 | -0.052 | -0.75 (-0.94, -0.57) | 2.80E-15 |
| CLU | Clusterin | 0.0304 | 1.22 (1.04, 1.41) | 1.40E-38 |
| CNGB3 | Cyclic nucleotide-gated cation channel beta-3 | 0.005 | 0.1 (-0.08, 0.28) | 0.2809 |
| CNST | Consortin | -0.0045 | 0.27 (0.08, 0.45) | 0.005 |

| | | | | |
|---|---|---|---|---|
| COL15A1 | Collagen alpha-1(XV) chain | 0.0778 | 0.21 (0.02, 0.41) | 0.0326 |
| COL1A1 | Collagen alpha-1(I) chain | -0.0419 | -0.25 (-0.44, -0.06) | 0.0084 |
| COL3A1 | Collagen alpha-1(III) chain | -0.0495 | -0.31 (-0.5, -0.13) | 0.001 |
| COL4A1 | Collagen alpha-1(IV) chain | -0.2023 | -1.46 (-1.65, -1.27) | 1.30E-51 |
| COL4A4 | Collagen alpha-4(IV) chain | -0.0193 | -0.15 (-0.33, 0.04) | 0.1138 |
| COL6A3 | Collagen alpha-3(VI) chain | -0.0423 | 1.82 (1.62, 2.01) | 3.70E-71 |
| COQ7 | 5-demethoxyubiquinone hydroxylase, mitochondrial | 0.1004 | 1.05 (0.86, 1.23) | 5.70E-29 |
| CPE | Carboxypeptidase E | -0.0569 | -1.07 (-1.25, -0.89) | 9.70E-30 |
| CPOX | Oxygen-dependent coproporphyrinogen-III oxidase, mitochondrial | -0.0024 | -0.79 (-0.98, -0.6) | 1.30E-16 |
| CPQ | Carboxypeptidase Q | 0.0674 | 1.12 (0.94, 1.31) | 1.60E-31 |
| CPVL | Probable serine carboxypeptidase CPVL | 0.0054 | 0.67 (0.48, 0.85) | 4.70E-12 |
| CRH | Corticoliberin | -0.0135 | -0.9 (-1.09, -0.72) | 4.70E-22 |
| CRIM1 | Cysteine-rich motor neuron 1 protein | 0.1255 | 0.56 (0.37, 0.75) | 1.10E-08 |
| CRISP2 | Cysteine-rich secretory protein 2 | -0.053 | -3.23 (-3.4, -3.05) | 2.50E-267 |
| CRYBB2 | Beta-crystallin B2 | -0.0412 | 0.14 (-0.05, 0.33) | 0.1562 |
| CSF3R | Granulocyte colony-stimulating factor receptor | 0.0548 | 1.06 (0.88, 1.25) | 2.30E-28 |
| CST5 | Cystatin-D | -0.0445 | -0.45 (-0.63, -0.26) | 3.80E-06 |
| CST6 | Cystatin-M | -0.0753 | -1.57 (-1.75, -1.38) | 1.90E-60 |
| CTBS | Di-N-acetylchitobiase | -0.083 | -0.05 (-0.25, 0.14) | 0.5753 |
| CTHRC1 | Collagen triple helix repeat-containing protein 1 | -0.0993 | -0.83 (-1.02, -0.64) | 4.40E-17 |
| CTSH | Pro-cathepsin H | 0.0532 | 0.18 (-0.01, 0.36) | 0.0662 |
| CTSO | Cathepsin O | 0.0015 | 0.65 (0.46, 0.84) | 4.30E-11 |
| CTSV | Cathepsin L2 | 0.0031 | -0.41 (-0.6, -0.22) | 2.10E-05 |
| CTSZ | Cathepsin Z | -0.0049 | 0.41 (0.22, 0.61) | 4.00E-05 |
| CXCL1 | Growth-regulated alpha protein | 0.0732 | 0.5 (0.31, 0.68) | 1.00E-07 |
| CXCL11 | C-X-C motif chemokine 11 | 0.0114 | 1.44 (1.25, 1.62) | 1.60E-52 |
| CXCL16 | C-X-C motif chemokine 16 | -0.0027 | 1.8 (1.62, 1.99) | 6.60E-78 |
| CXCL6 | C-X-C motif chemokine 6 | 0.0015 | 0.57 (0.38, 0.75) | 1.80E-09 |
| CYB5A | Cytochrome b5 | -0.0001 | -0.03 (-0.22, 0.15) | 0.7298 |
| CYTL1 | Cytokine-like protein 1 | -0.2192 | -1.94 (-2.12, -1.75) | 1.20E-89 |
| DCBLD2 | Discoidin, CUB and LCCL domain-containing protein 2 | 0.0013 | 0.68 (0.49, 0.88) | 5.60E-12 |
| DCLRE1C | Protein artemis | -0.0462 | -0.19 (-0.37, 0) | 0.048 |
| DDC | Aromatic-L-amino-acid decarboxylase | -0.0322 | -0.84 (-1.02, -0.65) | 1.00E-18 |
| DDR1 | Epithelial discoidin domain-containing receptor 1 | -0.0545 | -0.87 (-1.06, -0.68) | 3.80E-19 |
| DDT | D-dopachrome decarboxylase | 0.0199 | 0.02 (-0.16, 0.21) | 0.7925 |
| DDX1 | ATP-dependent RNA helicase DDX1 | 0.0197 | 0.03 (-0.16, 0.21) | 0.7876 |
| DDX53 | Probable ATP-dependent RNA helicase DDX53 | -0.056 | -0.39 (-0.58, -0.2) | 3.90E-05 |
| DEFB104A_DEFB104B | Beta-defensin 104 | -0.0028 | -1.83 (-2.01, -1.65) | 1.50E-85 |
| DENND2B | DENN domain-containing protein 2B | 0.0104 | 0.15 (-0.04, 0.33) | 0.1176 |
| DIPK2B | Divergent protein kinase domain 2B | 0.1487 | 1.48 (1.3, 1.67) | 6.20E-54 |
| DKK3 | Dickkopf-related protein 3 | -0.0643 | -0.75 (-0.94, -0.57) | 3.20E-15 |
| DMP1 | Dentin matrix acidic phosphoprotein 1 | 0.0545 | -0.03 (-0.22, 0.16) | 0.7496 |

| | | | | |
|---|---|---|---|---|
| DPEP1 | Dipeptidase 1 | -0.0294 | -1.7 (-1.88, -1.52) | 1.60E-72 |
| DPEP2 | Dipeptidase 2 | 0.01 | 0.68 (0.49, 0.87) | 1.40E-12 |
| DPP7 | Dipeptidyl peptidase 2 | -0.0479 | -0.25 (-0.44, -0.06) | 0.0096 |
| DPT | Dermatopontin | 0.1813 | 2.08 (1.89, 2.27) | 5.50E-102 |
| DSG2 | Desmoglein-2 | 0.0059 | 0.26 (0.08, 0.45) | 0.006 |
| DTX3 | Probable E3 ubiquitin-protein ligase DTX3 | -0.0241 | -0.31 (-0.51, -0.12) | 0.0015 |
| ECHDC3 | Enoyl-CoA hydratase domain-containing protein 3, mitochondrial | 0.0188 | 0.47 (0.28, 0.65) | 9.10E-07 |
| EFEMP1 | EGF-containing fibulin-like extracellular matrix protein 1 | 0.2057 | 2.28 (2.09, 2.47) | 3.70E-119 |
| EFNA1 | Ephrin-A1 | 0.0645 | 1.77 (1.57, 1.96) | 3.40E-70 |
| ELN | Elastin | 0.1144 | 1.96 (1.77, 2.14) | 1.20E-90 |
| ENDOU | Poly(U)-specific endoribonuclease | -0.0106 | -2.21 (-2.39, -2.02) | 1.50E-114 |
| ENG | Endoglin | -0.1451 | -1.29 (-1.47, -1.1) | 6.40E-42 |
| ENPP2 | Ectonucleotide pyrophosphatase/phosphodiesterase family member 2 | 0.0237 | 3.63 (3.45, 3.8) | 0.00E+00 |
| ENPP5 | Ectonucleotide pyrophosphatase/phosphodiesterase family member 5 | -0.0489 | -2.76 (-2.93, -2.59) | 2.30E-212 |
| ENPP6 | Glycerophosphocholine cholinephosphodiesterase ENPP6 | -0.1154 | -0.74 (-0.92, -0.55) | 5.60E-15 |
| EPN1 | Epsin-1 | -0.0013 | -0.02 (-0.2, 0.16) | 0.8208 |
| EPO | Erythropoietin | -0.0395 | 0.62 (0.44, 0.8) | 3.70E-11 |
| EPS8L2 | Epidermal growth factor receptor kinase substrate 8-like protein 2 | -0.0427 | 0.27 (0.08, 0.47) | 0.0049 |
| EXTL1 | Exostosin-like 1 | -0.0131 | -0.7 (-0.88, -0.51) | 1.70E-13 |
| F11 | Coagulation factor XI | 0.0233 | 1.85 (1.67, 2.04) | 7.80E-85 |
| FABP2 | Fatty acid-binding protein, intestinal | -0.1286 | -0.47 (-0.66, -0.28) | 1.10E-06 |
| FABP4 | Fatty acid-binding protein, adipocyte | 1.2469 | 5.65 (5.5, 5.8) | 0.00E+00 |
| FAM171A2 | Protein FAM171A2 | -0.0109 | -0.01 (-0.19, 0.18) | 0.9333 |
| FAM3B | Protein FAM3B | -0.0434 | -0.71 (-0.9, -0.52) | 3.00E-13 |
| FAM3C | Protein FAM3C | -0.0936 | -0.24 (-0.44, -0.04) | 0.0173 |
| FCGR3B | Low affinity immunoglobulin gamma Fc region receptor III-B | 0.0043 | 0.91 (0.73, 1.1) | 9.20E-23 |
| FCRLB | Fc receptor-like B | -0.0054 | -0.63 (-0.82, -0.44) | 1.00E-10 |
| FGF5 | Fibroblast growth factor 5 | 0.0052 | 0.27 (0.08, 0.46) | 0.0047 |
| FGFBP3 | Fibroblast growth factor-binding protein 3 | 0.0036 | 0.95 (0.76, 1.13) | 3.70E-23 |
| FGL1 | Fibrinogen-like protein 1 | -0.0181 | 1.53 (1.35, 1.72) | 2.20E-58 |
| FH | Fumarate hydratase, mitochondrial | 0.0289 | 0.31 (0.12, 0.49) | 0.0012 |
| FLRT2 | Leucine-rich repeat transmembrane protein FLRT2 | -0.0046 | 0.61 (0.42, 0.8) | 3.60E-10 |
| FLT4 | Vascular endothelial growth factor receptor 3 | 0.0631 | 1.38 (1.19, 1.56) | 3.00E-46 |
| FNDC1 | Fibronectin type III domain-containing protein 1 | 0.0096 | 0.99 (0.8, 1.18) | 5.00E-24 |
| FOLR2 | Folate receptor beta | 0.0336 | 2.62 (2.44, 2.8) | 1.10E-169 |
| FOLR3 | Folate receptor gamma | 0.0286 | 1.43 (1.25, 1.61) | 9.00E-54 |
| FURIN | Furin | 0.0208 | 2.66 (2.47, 2.84) | 8.00E-170 |
| GALNT5 | Polypeptide N-acetylgalactosaminyltransferase 5 | -0.0098 | -0.71 (-0.89, -0.52) | 1.40E-13 |
| GASK1A | Golgi-associated kinase 1A | 0.0077 | 0.9 (0.72, 1.09) | 4.20E-21 |
| GBP6 | Guanylate-binding protein 6 | 0.0258 | 0.01 (-0.18, 0.19) | 0.9298 |
| GC | Vitamin D-binding protein | -0.0235 | 0.46 (0.27, 0.64) | 1.30E-06 |
| GH1 | Somatotropin | -0.0382 | 0.86 (0.68, 1.04) | 3.60E-21 |
| GH2 | Growth hormone variant | 0.001 | 0.35 (0.16, 0.53) | 0.0002 |

| | | | | |
|---|---|---|---|---|
| GHRL | Appetite-regulating hormone | -0.0548 | 0.34 (0.15, 0.53) | 0.0004 |
| GMPR | GMP reductase 1 | -0.0036 | -0.97 (-1.15, -0.78) | 3.10E-25 |
| GNLY | Granulysin | 0.0074 | 1.2 (1.02, 1.39) | 1.70E-35 |
| GNPDA2 | Glucosamine-6-phosphate isomerase 2 | -0.0402 | -0.02 (-0.21, 0.17) | 0.8247 |
| GP2 | Pancreatic secretory granule membrane major glycoprotein GP2 | -0.0028 | -1.52 (-1.71, -1.33) | 1.00E-55 |
| GPD1 | Glycerol-3-phosphate dehydrogenase | 0.0394 | 2.27 (2.08, 2.45) | 2.90E-127 |
| GRAP2 | GRB2-related adapter protein 2 | -0.1711 | 0.28 (0.1, 0.47) | 0.003 |
| GSTA1 | Glutathione S-transferase A1 | -0.0712 | 0.83 (0.64, 1.02) | 8.20E-18 |
| GZMA | Granzyme A | 0.0769 | 1.62 (1.44, 1.81) | 2.10E-64 |
| GZMB | Granzyme B | -0.0511 | -1.04 (-1.23, -0.86) | 4.10E-29 |
| HDAC8 | Histone deacetylase 8 | -0.0301 | -0.04 (-0.23, 0.14) | 0.6507 |
| HGFAC | Hepatocyte growth factor activator | 0.0324 | 1.21 (1.02, 1.39) | 3.00E-36 |
| HK2 | Hexokinase-2 | 0.004 | 0.2 (0.02, 0.39) | 0.0278 |
| HMOX1 | Heme oxygenase 1 | -0.0255 | -0.85 (-1.04, -0.66) | 1.10E-18 |
| HRG | Histidine-rich glycoprotein | 0.0295 | 0.98 (0.8, 1.17) | 6.10E-25 |
| HS6ST2 | Heparan-sulfate 6-O-sulfotransferase 2 | -0.1868 | -2.24 (-2.43, -2.05) | 5.30E-119 |
| HSPG2 | Basement membrane-specific heparan sulfate proteoglycan core protein | 0.0023 | 1.2 (1, 1.4) | 1.50E-31 |
| ICAM4 | Intercellular adhesion molecule 4 | 0.0213 | -0.27 (-0.45, -0.08) | 0.0052 |
| ICOSLG | ICOS ligand | -0.1129 | -0.83 (-1.01, -0.64) | 2.80E-18 |
| IDUA | Alpha-L-iduronidase | -0.0005 | 1.13 (0.94, 1.32) | 8.00E-31 |
| IFNAR1 | Interferon alpha/beta receptor 1 | 0.03 | 0.63 (0.45, 0.82) | 1.40E-11 |
| IFNL2 | Interferon lambda-2 | 0.0204 | 0.15 (-0.03, 0.34) | 0.0984 |
| IGDCC4 | Immunoglobulin superfamily DCC subclass member 4 | -0.013 | -1.99 (-2.18, -1.81) | 4.00E-98 |
| IGFBP2 | Insulin-like growth factor-binding protein 2 | -0.259 | -1.43 (-1.62, -1.24) | 4.80E-50 |
| IGFBP6 | Insulin-like growth factor-binding protein 6 | -0.0061 | -1.74 (-1.94, -1.55) | 1.20E-67 |
| IGFBP7 | Insulin-like growth factor-binding protein 7 | -0.1833 | -0.8 (-0.99, -0.6) | 9.10E-16 |
| IL12B | Interleukin-12 subunit beta | 0.0172 | 1.99 (1.8, 2.18) | 1.00E-94 |
| IL17C | Interleukin-17C | 0.0415 | -0.59 (-0.78, -0.4) | 9.30E-10 |
| IL19 | Interleukin-19 | -0.0424 | -0.42 (-0.61, -0.23) | 1.80E-05 |
| IL1RL1 | Interleukin-1 receptor-like 1 | -0.0416 | -2.01 (-2.19, -1.83) | 3.70E-103 |
| IL1RL2 | Interleukin-1 receptor-like 2 | 0.1683 | 1.73 (1.54, 1.91) | 2.10E-73 |
| IL2RA | Interleukin-2 receptor subunit alpha | -0.0119 | 0.52 (0.32, 0.71) | 1.80E-07 |
| IL5RA | Interleukin-5 receptor subunit alpha | 0.0108 | 0.31 (0.12, 0.5) | 0.0017 |
| IL6R | Interleukin-6 receptor subunit alpha | 0.0222 | 0.65 (0.46, 0.84) | 1.20E-11 |
| IMPG1 | Interphotoreceptor matrix proteoglycan 1 | 0.0213 | 0.21 (0.03, 0.4) | 0.0256 |
| INSL4 | Early placenta insulin-like peptide | 0.0047 | -0.28 (-0.46, -0.1) | 0.003 |
| IPCEF1 | Interactor protein for cytohesin exchange factors 1 | -0.0314 | 0.01 (-0.18, 0.19) | 0.9277 |
| ITGA5 | Integrin alpha-5 | 0.099 | 2.05 (1.86, 2.24) | 1.10E-98 |
| ITGAL | Integrin alpha-L | 0.0017 | 1.24 (1.05, 1.42) | 1.20E-37 |
| ITGB6 | Integrin beta-6 | -0.1259 | -2.77 (-2.95, -2.6) | 4.00E-194 |
| ITIH4 | Inter-alpha-trypsin inhibitor heavy chain H4 | 0.0574 | 2.98 (2.81, 3.16) | 2.80E-231 |
| JCHAIN | Immunoglobulin J chain | 0.024 | 0.66 (0.47, 0.85) | 4.40E-12 |

| | | | | |
|---|---|---|---|---|
| KCNC4 | Potassium voltage-gated channel subfamily C member 4 | 0.0462 | 0.31 (0.12, 0.49) | 0.0012 |
| KIAA0319 | Dyslexia-associated protein KIAA0319 | -0.0051 | -0.39 (-0.58, -0.2) | 0.0001 |
| KITLG | Kit ligand | -0.0786 | -0.55 (-0.74, -0.35) | 2.50E-08 |
| KLK1 | Kallikrein-1 | -0.0052 | -0.39 (-0.58, -0.2) | 0.0001 |
| KLK13 | Kallikrein-13 | -0.06 | -1.15 (-1.33, -0.96) | 1.50E-32 |
| KLK15 | Kallikrein-15 | -0.0065 | -0.04 (-0.23, 0.15) | 0.6749 |
| KLK3 | Prostate-specific antigen | 0.0671 | -4.38 (-4.56, -4.21) | 0.00E+00 |
| KLRD1 | Natural killer cells antigen CD94 | 0.0062 | 1.2 (1.01, 1.39) | 5.50E-36 |
| KRT5 | Keratin, type II cytoskeletal 5 | -0.0087 | -1.46 (-1.65, -1.28) | 2.20E-54 |
| LAIR1 | Leukocyte-associated immunoglobulin-like receptor 1 | 0.0058 | 2.1 (1.91, 2.3) | 5.40E-98 |
| LAMB1 | Laminin subunit beta-1 | 0.0336 | 0.73 (0.54, 0.92) | 4.20E-14 |
| LAMP2 | Lysosome-associated membrane glycoprotein 2 | 0.0633 | 2.13 (1.94, 2.32) | 2.50E-107 |
| LCN15 | Lipocalin-15 | -0.0119 | -0.71 (-0.9, -0.53) | 7.50E-14 |
| LEFTY2 | Left-right determination factor 2 | 0.123 | 1.65 (1.47, 1.83) | 2.30E-72 |
| LEP | Leptin | 3.0453 | 7.07 (6.95, 7.18) | 0.00E+00 |
| LEPR | Leptin receptor | -0.0097 | -0.59 (-0.78, -0.41) | 5.20E-10 |
| LGALS3 | Galectin-3 | 0.0099 | 2.04 (1.85, 2.23) | 5.10E-98 |
| LGALS3BP | Galectin-3-binding protein | 0.0416 | 2.42 (2.24, 2.6) | 1.50E-143 |
| LGALS7_LGALS7B | Galectin-7 | -0.218 | -1.09 (-1.28, -0.89) | 1.40E-28 |
| LGALS9 | Galectin-9 | 0.0972 | 2.52 (2.33, 2.71) | 3.20E-144 |
| LIFR | Leukemia inhibitory factor receptor | 0.0385 | 0.16 (-0.03, 0.35) | 0.1056 |
| LILRA2 | Leukocyte immunoglobulin-like receptor subfamily A member 2 | 0.0274 | 2.21 (2.02, 2.39) | 4.00E-116 |
| LILRA3 | Leukocyte immunoglobulin-like receptor subfamily A member 3 | 0.0064 | 1.18 (0.99, 1.37) | 8.50E-35 |
| LMOD1 | Leiomodin-1 | -0.0249 | -0.7 (-0.89, -0.51) | 3.60E-13 |
| LPCAT2 | Lysophosphatidylcholine acyltransferase 2 | 0.004 | 0.9 (0.72, 1.09) | 1.40E-21 |
| LRP1 | Prolow-density lipoprotein receptor-related protein 1 | 0.022 | 0.63 (0.44, 0.81) | 1.80E-11 |
| LRRC38 | Leucine-rich repeat-containing protein 38 | -0.0719 | -1.25 (-1.43, -1.06) | 1.50E-39 |
| LRRN1 | Leucine-rich repeat neuronal protein 1 | -0.0449 | -1.28 (-1.46, -1.09) | 1.10E-41 |
| LXN | Latexin | 0.0174 | -0.61 (-0.79, -0.42) | 1.80E-10 |
| LYAR | Cell growth-regulating nucleolar protein | -0.0128 | 0.06 (-0.13, 0.24) | 0.5524 |
| MAN2B2 | Epididymis-specific alpha-mannosidase | -0.0004 | -0.39 (-0.58, -0.19) | 0.0001 |
| MATN3 | Matrilin-3 | -0.0314 | -1.26 (-1.45, -1.07) | 1.80E-38 |
| MB | Myoglobin | -0.1065 | -1.96 (-2.14, -1.78) | 9.00E-96 |
| MBL2 | Mannose-binding protein C | -0.0028 | -0.81 (-1, -0.62) | 2.40E-17 |
| MCAM | Cell surface glycoprotein MUC18 | 0.0862 | 0.11 (-0.07, 0.3) | 0.2421 |
| MELTF | Melanotransferrin | 0.027 | 0.6 (0.41, 0.78) | 3.50E-10 |
| MEPE | Matrix extracellular phosphoglycoprotein | -0.1609 | -1.66 (-1.84, -1.48) | 4.00E-69 |
| MFGE8 | Lactadherin | -0.0303 | -0.54 (-0.73, -0.35) | 2.50E-08 |
| MIA | Melanoma-derived growth regulatory protein | -0.0328 | -0.59 (-0.77, -0.4) | 1.00E-09 |
| MLN | Promotilin | 0.0727 | 0.54 (0.35, 0.73) | 2.40E-08 |
| MMP1 | Interstitial collagenase | -0.0356 | 0.4 (0.21, 0.58) | 3.90E-05 |
| MMP3 | Stromelysin-1 | -0.33 | -3.96 (-4.13, -3.79) | 0.00E+00 |

| | | | | |
|---|---|---|---|---|
| MMP7 | Matrilysin | 0.0589 | 1.66 (1.46, 1.85) | 8.70E-59 |
| MOG | Myelin-oligodendrocyte glycoprotein | -0.1323 | -0.24 (-0.43, -0.05) | 0.014 |
| MPO | Myeloperoxidase | 0.082 | 1.01 (0.82, 1.19) | 1.50E-26 |
| MSMB | Beta-microseminoprotein | -0.0777 | -1.98 (-2.17, -1.8) | 5.50E-92 |
| MST1 | Hepatocyte growth factor-like protein | -0.0161 | 0.43 (0.23, 0.62) | 1.40E-05 |
| MUC13 | Mucin-13 | -0.0436 | 0.21 (0.02, 0.41) | 0.0308 |
| MUCL3 | Mucin-like protein 3 | 0.0158 | 0.3 (0.12, 0.49) | 0.0013 |
| NAAA | N-acylethanolamine-hydrolyzing acid amidase | -0.0224 | -0.55 (-0.74, -0.36) | 1.20E-08 |
| NCAM1 | Neural cell adhesion molecule 1 | -0.1196 | -2.32 (-2.5, -2.14) | 1.50E-133 |
| NCR3LG1 | Natural cytotoxicity triggering receptor 3 ligand 1 | -0.1467 | -1.84 (-2.03, -1.65) | 3.50E-81 |
| NCS1 | Neuronal calcium sensor 1 | -0.0767 | -1.27 (-1.46, -1.08) | 4.70E-39 |
| NECAP2 | Adaptin ear-binding coat-associated protein 2 | 0.0002 | 0.5 (0.32, 0.69) | 1.10E-07 |
| NECTIN4 | Nectin-4 | -0.0053 | -0.78 (-0.97, -0.58) | 1.10E-14 |
| NEFL | Neurofilament light polypeptide | -0.1199 | -0.15 (-0.35, 0.04) | 0.1252 |
| NOS2 | Nitric oxide synthase, inducible | 0.0312 | 0.05 (-0.14, 0.23) | 0.6084 |
| NPPC | C-type natriuretic peptide | -0.3373 | -2.35 (-2.53, -2.16) | 1.10E-129 |
| NPTX2 | Neuronal pentraxin-2 | -0.1173 | -3.25 (-3.42, -3.07) | 3.40E-271 |
| NPTXR | Neuronal pentraxin receptor | -0.0245 | -0.44 (-0.63, -0.25) | 5.40E-06 |
| NRP2 | Neuropilin-2 | 0.001 | 0.97 (0.78, 1.15) | 1.70E-23 |
| NTF3 | Neurotrophin-3 | -0.0755 | -0.58 (-0.77, -0.4) | 1.00E-09 |
| NTRK2 | BDNF/NT-3 growth factors receptor | 0.0757 | 1.25 (1.06, 1.44) | 7.20E-39 |
| OBP2B | Odorant-binding protein 2b | -0.2525 | -3.67 (-3.84, -3.5) | 0.00E+00 |
| OLFM4 | Olfactomedin-4 | -0.03 | 0.07 (-0.13, 0.26) | 0.4971 |
| OLR1 | Oxidized low-density lipoprotein receptor 1 | -0.1181 | 0.27 (0.08, 0.45) | 0.0049 |
| OMG | Oligodendrocyte-myelin glycoprotein | 0.0234 | -0.21 (-0.4, -0.02) | 0.0295 |
| OPTC | Opticin | -0.0097 | 0.56 (0.37, 0.75) | 5.40E-09 |
| PADI2 | Protein-arginine deiminase type-2 | -0.0088 | -0.27 (-0.45, -0.08) | 0.0045 |
| PALM | Paralemmin-1 | 0.0467 | 2.69 (2.51, 2.88) | 9.60E-169 |
| PBLD | Phenazine biosynthesis-like domain-containing protein | 0.0196 | 0.17 (-0.02, 0.36) | 0.0725 |
| PCDH17 | Protocadherin-17 | -0.0271 | -0.51 (-0.7, -0.32) | 1.80E-07 |
| PDGFRA | Platelet-derived growth factor receptor alpha | -0.1589 | -0.41 (-0.6, -0.22) | 1.90E-05 |
| PENK | Proenkephalin-A | -0.0498 | 0.37 (0.18, 0.57) | 0.0002 |
| PEPD | Xaa-Pro dipeptidase | -0.0164 | -0.03 (-0.22, 0.16) | 0.7788 |
| PGA4 | Pepsin A-4 | -0.0022 | -1.45 (-1.65, -1.25) | 1.90E-45 |
| PGF | Placenta growth factor | -0.0063 | -0.3 (-0.5, -0.1) | 0.0034 |
| PI3 | Elafin | -0.0369 | -0.83 (-1.02, -0.64) | 8.60E-18 |
| PLA2G1B | Phospholipase A2 | -0.0466 | 0.02 (-0.17, 0.21) | 0.8165 |
| PLB1 | Phospholipase B1, membrane-associated | -0.0195 | -1.94 (-2.12, -1.77) | 9.50E-101 |
| PLIN1 | Perilipin-1 | 0.019 | 2.4 (2.22, 2.58) | 6.00E-142 |
| PLXDC1 | Plexin domain-containing protein 1 | -0.012 | -0.71 (-0.89, -0.52) | 4.20E-14 |
| PODXL | Podocalyxin | 0.0049 | -0.16 (-0.34, 0.03) | 0.0966 |
| PODXL2 | Podocalyxin-like protein 2 | -0.0885 | -1.04 (-1.23, -0.85) | 5.60E-27 |
| POLR2A | DNA-directed RNA polymerase II subunit RPB1 | -0.0094 | -0.02 (-0.21, 0.16) | 0.8015 |

| | | | | |
|---|---|---|---|---|
| PRCP | Lysosomal Pro-X carboxypeptidase | 0.0158 | 1.02 (0.83, 1.21) | 5.10E-25 |
| PREB | Prolactin regulatory element-binding protein | -0.0484 | -0.07 (-0.26, 0.12) | 0.4879 |
| PRRT3 | Proline-rich transmembrane protein 3 | -0.1134 | -0.05 (-0.25, 0.14) | 0.5803 |
| PRSS27 | Serine protease 27 | -0.0005 | -0.23 (-0.41, -0.04) | 0.018 |
| PRSS53 | Serine protease 53 | -0.0112 | -0.42 (-0.6, -0.23) | 1.10E-05 |
| PRTG | Protogenin | -0.0314 | -1.07 (-1.26, -0.89) | 1.50E-29 |
| PSCA | Prostate stem cell antigen | 0.0107 | -0.1 (-0.29, 0.09) | 0.3137 |
| PTGDS | Prostaglandin-H2 D-isomerase | -0.168 | -0.86 (-1.05, -0.66) | 2.50E-17 |
| PTH1R | Parathyroid hormone/parathyroid hormone-related peptide receptor | 0.0546 | 0.21 (0.03, 0.4) | 0.0245 |
| PTPRC | Receptor-type tyrosine-protein phosphatase C | 0.0614 | 1.35 (1.17, 1.54) | 6.60E-46 |
| PTPRK | Receptor-type tyrosine-protein phosphatase kappa | 0.0632 | 0.91 (0.73, 1.1) | 1.20E-21 |
| PTS | 6-pyruvoyl tetrahydrobiopterin synthase | 0.0039 | 0.64 (0.45, 0.83) | 6.70E-11 |
| PTX3 | Pentraxin-related protein PTX3 | -0.0928 | -0.27 (-0.45, -0.08) | 0.0042 |
| PYDC1 | Pyrin domain-containing protein 1 | 0.0179 | -0.66 (-0.85, -0.48) | 4.00E-12 |
| PZP | Pregnancy zone protein | 0.1788 | 4.5 (4.34, 4.65) | 0.00E+00 |
| QPCT | Glutaminyl-peptide cyclotransferase | 0.0053 | 0.64 (0.45, 0.83) | 8.50E-11 |
| RARRES2 | Retinoic acid receptor responder protein 2 | 0.1433 | 2.93 (2.75, 3.11) | 2.10E-208 |
| RECK | Reversion-inducing cysteine-rich protein with Kazal motifs | 0.1162 | 1.42 (1.24, 1.6) | 3.40E-52 |
| REG1B | Lithostathine-1-beta | -0.0455 | -0.46 (-0.64, -0.27) | 1.40E-06 |
| REG3G | Regenerating islet-derived protein 3-gamma | -0.0003 | -0.72 (-0.91, -0.53) | 4.10E-14 |
| RELT | Tumor necrosis factor receptor superfamily member 19L | -0.2124 | -1.37 (-1.57, -1.18) | 8.80E-44 |
| REN | Renin | -0.1045 | -1.45 (-1.67, -1.22) | 1.40E-36 |
| RGMA | Repulsive guidance molecule A | -0.1097 | -1.89 (-2.08, -1.71) | 1.90E-87 |
| ROBO1 | Roundabout homolog 1 | 0.013 | 1.48 (1.29, 1.67) | 5.20E-51 |
| RPL14 | 60S ribosomal protein L14 | 0.0024 | 0.75 (0.57, 0.93) | 9.00E-16 |
| SCG2 | Secretogranin-2 | -0.0341 | -0.52 (-0.72, -0.33) | 1.30E-07 |
| SCGB3A1 | Secretoglobin family 3A member 1 | -0.0767 | 0.13 (-0.06, 0.32) | 0.176 |
| SCGB3A2 | Secretoglobin family 3A member 2 | -0.0432 | -0.92 (-1.1, -0.73) | 1.50E-21 |
| SCRG1 | Scrapie-responsive protein 1 | -0.145 | -0.66 (-0.85, -0.46) | 3.30E-11 |
| SERPINA1 | Alpha-1-antitrypsin | -0.0145 | 0.27 (0.08, 0.46) | 0.0047 |
| SERPINA4 | Kallistatin | -0.0076 | -0.22 (-0.41, -0.03) | 0.0218 |
| SERPIND1 | Heparin cofactor 2 | 0.0386 | 2.63 (2.44, 2.81) | 8.50E-167 |
| SERPING1 | Plasma protease C1 inhibitor | 0.0297 | 1.31 (1.13, 1.5) | 2.80E-43 |
| SEZ6L2 | Seizure 6-like protein 2 | 0.2702 | 1.91 (1.73, 2.09) | 2.00E-94 |
| SF3B4 | Splicing factor 3B subunit 4 | 0.0659 | 0.32 (0.14, 0.51) | 0.0006 |
| SFRP1 | Secreted frizzled-related protein 1 | 0.308 | 2.15 (1.96, 2.34) | 2.00E-107 |
| SFTPD | Pulmonary surfactant-associated protein D | -0.0812 | -0.28 (-0.47, -0.09) | 0.0046 |
| SHPK | Sedoheptulokinase | -0.0141 | -0.18 (-0.36, 0.01) | 0.0577 |
| SIGLEC10 | Sialic acid-binding Ig-like lectin 10 | 0.0526 | 1.23 (1.04, 1.43) | 2.40E-35 |
| SIRPA | Tyrosine-protein phosphatase non-receptor type substrate 1 | -0.0091 | 0.5 (0.31, 0.69) | 2.40E-07 |
| SLC9A3R2 | Na(+)/H(+) exchange regulatory cofactor NHE-RF2 | -0.0003 | -0.58 (-0.77, -0.4) | 2.70E-10 |
| SLIT2 | Slit homolog 2 protein | 0.0032 | 0.47 (0.28, 0.66) | 7.70E-07 |
| SLITRK2 | SLIT and NTRK-like protein 2 | 0.1278 | 1.97 (1.78, 2.15) | 3.30E-93 |

| | | | | |
|---|---|---|---|---|
| SMOC1 | SPARC-related modular calcium-binding protein 1 | -0.0394 | 0.32 (0.13, 0.52) | 0.001 |
| SMOC2 | SPARC-related modular calcium-binding protein 2 | -0.1115 | -0.95 (-1.13, -0.76) | 6.70E-23 |
| SMPD1 | Sphingomyelin phosphodiesterase | -0.0642 | -0.75 (-0.94, -0.56) | 6.60E-15 |
| SNCG | Gamma-synuclein | 0.0614 | 2.58 (2.39, 2.77) | 4.30E-148 |
| SOD2 | Superoxide dismutase [Mn], mitochondrial | -0.1096 | -1.48 (-1.66, -1.29) | 3.80E-53 |
| SPESP1 | Sperm equatorial segment protein 1 | -0.0239 | -2.09 (-2.27, -1.9) | 6.20E-109 |
| SPINK6 | Serine protease inhibitor Kazal-type 6 | -0.1922 | -0.54 (-0.73, -0.35) | 1.40E-08 |
| SPON1 | Spondin-1 | 0.0196 | 1.04 (0.85, 1.23) | 5.90E-26 |
| SPON2 | Spondin-2 | -0.0285 | 1.54 (1.34, 1.73) | 2.10E-53 |
| SSC4D | Scavenger receptor cysteine-rich domain-containing group B protein | -0.0092 | 0.02 (-0.17, 0.21) | 0.8352 |
| ST6GAL1 | Beta-galactoside alpha-2,6-sialyltransferase 1 | 0.0622 | 1.83 (1.64, 2.01) | 2.90E-82 |
| STX1B | Syntaxin-1B | -0.008 | 0.03 (-0.16, 0.21) | 0.7549 |
| SUSD5 | Sushi domain-containing protein 5 | -0.0097 | -1.63 (-1.82, -1.44) | 2.00E-60 |
| TBC1D17 | TBC1 domain family member 17 | 0.0115 | -0.18 (-0.36, 0.01) | 0.0572 |
| TEX101 | Testis-expressed protein 101 | -0.0898 | -3.73 (-3.9, -3.56) | 0.00E+00 |
| TFF2 | Trefoil factor 2 | -0.0082 | 0.52 (0.34, 0.71) | 4.10E-08 |
| TFRC | Transferrin receptor protein 1 | 0.0251 | 0.64 (0.46, 0.83) | 8.10E-12 |
| TG | Thyroglobulin | 0.065 | 1.05 (0.87, 1.23) | 1.00E-28 |
| TGFBR2 | TGF-beta receptor type-2 | 0.1827 | 1.85 (1.65, 2.04) | 6.40E-76 |
| TGM2 | Protein-glutamine gamma-glutamyltransferase 2 | -0.0259 | -0.55 (-0.74, -0.37) | 5.40E-09 |
| THBS2 | Thrombospondin-2 | -0.0224 | 0.88 (0.68, 1.07) | 5.40E-19 |
| THBS4 | Thrombospondin-4 | 0.1579 | 1.86 (1.68, 2.05) | 2.00E-83 |
| THOP1 | Thimet oligopeptidase | -0.0952 | -0.48 (-0.67, -0.29) | 5.60E-07 |
| THY1 | Thy-1 membrane glycoprotein | 0.2221 | 2.72 (2.53, 2.91) | 1.50E-170 |
| TIMP4 | Metalloproteinase inhibitor 4 | 0.151 | 2.29 (2.1, 2.47) | 7.50E-127 |
| TNC | Tenascin | -0.118 | -0.07 (-0.26, 0.12) | 0.4549 |
| TNFRSF19 | Tumor necrosis factor receptor superfamily member 19 | -0.0508 | -0.62 (-0.82, -0.43) | 5.10E-10 |
| TNFRSF6B | Tumor necrosis factor receptor superfamily member 6B | -0.0724 | 0.46 (0.26, 0.65) | 4.90E-06 |
| TNFSF10 | Tumor necrosis factor ligand superfamily member 10 | 0.0665 | 0.76 (0.58, 0.95) | 9.10E-16 |
| TNFSF12 | Tumor necrosis factor ligand superfamily member 12 | 0.011 | 0.35 (0.17, 0.54) | 0.0002 |
| TNNI3 | Troponin I, cardiac muscle | 0.0012 | -0.41 (-0.6, -0.22) | 2.00E-05 |
| TOP1 | DNA topoisomerase 1 | 0.0046 | -0.88 (-1.06, -0.69) | 1.20E-20 |
| TPSD1 | Tryptase delta | 0.0589 | 0.26 (0.07, 0.44) | 0.0067 |
| TPSG1 | Tryptase gamma | 0.0129 | 0.2 (0.01, 0.38) | 0.0371 |
| TREM2 | Triggering receptor expressed on myeloid cells 2 | 0.0341 | 1.8 (1.61, 1.99) | 7.10E-73 |
| TREML2 | Trem-like transcript 2 protein | 0.0107 | 1.26 (1.07, 1.45) | 8.20E-39 |
| TSPAN1 | Tetraspanin-1 | -0.0162 | -0.48 (-0.68, -0.28) | 2.30E-06 |
| TYRP1 | 5,6-dihydroxyindole-2-carboxylic acid oxidase | -0.0194 | 0.07 (-0.12, 0.26) | 0.4751 |
| UMOD | Uromodulin | -0.0314 | 0.48 (0.29, 0.66) | 3.60E-07 |
| UNG | Uracil-DNA glycosylase | -0.0273 | 0 (-0.19, 0.18) | 0.9663 |
| UXS1 | UDP-glucuronic acid decarboxylase 1 | -0.0605 | -1.12 (-1.3, -0.93) | 8.40E-32 |
| VAMP5 | Vesicle-associated membrane protein 5 | 0.009 | 0.12 (-0.06, 0.31) | 0.1891 |
| VEGFB | Vascular endothelial growth factor B | 0.0535 | 1.83 (1.64, 2.02) | 1.80E-78 |

| | | | | |
|---|---|---|---|---|
| VIT | Vitrin | -0.073 | -1.78 (-1.97, -1.6) | 1.20E-80 |
| VSIR | V-type immunoglobulin domain-containing suppressor of T-cell activation | -0.0175 | 0.2 (0.01, 0.38) | 0.0357 |
| VWA1 | von Willebrand factor A domain-containing protein 1 | -0.0498 | 0.97 (0.79, 1.16) | 2.60E-24 |
| VWC2L | von Willebrand factor C domain-containing protein 2-like | -0.0068 | 0.23 (0.04, 0.42) | 0.019 |
| WFIKKN2 | WAP, Kazal, immunoglobulin, Kunitz and NTR domain-containing protein 2 | -0.06 | -1.18 (-1.37, -0.99) | 2.50E-34 |
| XG | Glycoprotein Xg | 0.3044 | 4.68 (4.52, 4.84) | 0.00E+00 |

CI: confidence interval.
*Linear regressions of the measured body fat percentage regressed on each of the proteins (per standard deviation).

**Supplemental Table 2C. Association between the 176 LASSO Selected Proteins and Waist-hip Ratio in the Training Set of the Healthy Cohort.**

| Protein | Name | LASSO | Linear Regression* | |
|---|---|---|---|---|
| | | Beta | Beta (95% CI) | P |
| ACE2 | Angiotensin-converting enzyme 2 | 0.0012 | 0.033 (0.031, 0.035) | 2.70E-248 |
| ACRV1 | Acrosomal protein SP-10 | 0.0023 | 0.041 (0.039, 0.043) | 0.00E+00 |
| ACY1 | Aminoacylase-1 | 0.0007 | 0.037 (0.035, 0.039) | 0.00E+00 |
| ADGRG2 | Adhesion G-protein coupled receptor G2 | -0.003 | -0.029 (-0.031, -0.027) | 9.20E-211 |
| ADH4 | All-trans-retinol dehydrogenase [NAD(+)] ADH4 | 0.0005 | 0.033 (0.031, 0.035) | 3.00E-274 |
| ADIPOQ | Adiponectin | -0.0035 | -0.043 (-0.044, -0.041) | 0.00E+00 |
| AGER | Advanced glycosylation end product-specific receptor | -0.0004 | -0.014 (-0.016, -0.012) | 8.90E-50 |
| ANGPT2 | Angiopoietin-2 | -0.0003 | -0.012 (-0.014, -0.01) | 1.60E-34 |
| ANGPTL3 | Angiopoietin-related protein 3 | -0.0012 | -0.016 (-0.018, -0.014) | 2.70E-63 |
| ANPEP | Aminopeptidase N | -0.0003 | 0.007 (0.005, 0.009) | 2.70E-14 |
| APCS | Serum amyloid P-component | 0.0037 | 0.042 (0.04, 0.043) | 0.00E+00 |
| APOA1 | Apolipoprotein A-I | -0.0023 | -0.024 (-0.026, -0.022) | 1.60E-144 |
| BAG3 | BAG family molecular chaperone regulator 3 | 0.0001 | 0.017 (0.015, 0.018) | 4.80E-65 |
| BCAN | Brevican core protein | -8.24E-06 | -0.017 (-0.019, -0.015) | 4.40E-69 |
| BCHE | Cholinesterase | 0.0003 | 0.026 (0.024, 0.028) | 4.00E-164 |
| BMP10 | Bone morphogenetic protein 10 | -0.0004 | -0.024 (-0.026, -0.022) | 1.70E-149 |
| C1QL2 | Complement C1q-like protein 2 | -0.0006 | -0.015 (-0.017, -0.013) | 1.50E-53 |
| CCL11 | Eotaxin | 0.0002 | 0.009 (0.007, 0.011) | 1.10E-22 |
| CCN5 | CCN family member 5 | 0.0002 | 0.022 (0.02, 0.024) | 8.40E-108 |
| CD300LG | CMRF35-like molecule 9 | -0.0056 | -0.023 (-0.025, -0.021) | 5.20E-130 |
| CD59 | CD59 glycoprotein | 0.001 | 0.028 (0.026, 0.03) | 2.50E-163 |
| CD99 | CD99 antigen | 0.0045 | 0.03 (0.028, 0.031) | 1.20E-214 |
| CDH15 | Cadherin-15 | 0.0004 | 0.03 (0.028, 0.032) | 3.50E-205 |
| CDHR2 | Cadherin-related family member 2 | 0.0026 | 0.045 (0.043, 0.046) | 0.00E+00 |
| CDHR5 | Cadherin-related family member 5 | 1.31E-05 | 0.017 (0.015, 0.019) | 3.50E-67 |
| CEACAM5 | Carcinoembryonic antigen-related cell adhesion molecule 5 | 0.0003 | 0.005 (0.003, 0.007) | 6.00E-08 |
| CFD | Complement factor D | 0.0024 | 0.02 (0.019, 0.022) | 6.70E-95 |
| CFH | Complement factor H | 0.0005 | 0.026 (0.024, 0.028) | 7.40E-154 |
| CFP | Properdin | 0.0005 | 0.026 (0.024, 0.028) | 3.90E-174 |
| CHGB | Secretogranin-1 | -0.001 | -0.009 (-0.011, -0.007) | 7.70E-21 |
| CLEC4A | C-type lectin domain family 4 member A | -0.0002 | -0.015 (-0.016, -0.013) | 2.40E-54 |
| CLEC5A | C-type lectin domain family 5 member A | -0.001 | -0.016 (-0.018, -0.014) | 1.10E-58 |
| CNTN1 | Contactin-1 | -0.0019 | -0.028 (-0.03, -0.026) | 5.30E-198 |
| CNTN3 | Contactin-3 | 0.0019 | 0.035 (0.033, 0.036) | 6.90E-301 |
| CNTN5 | Contactin-5 | -0.0006 | -0.022 (-0.024, -0.021) | 8.40E-123 |
| CNTNAP2 | Contactin-associated protein-like 2 | -0.0004 | -0.017 (-0.018, -0.015) | 2.20E-66 |
| COL4A1 | Collagen alpha-1(IV) chain | -0.002 | -0.02 (-0.022, -0.019) | 3.30E-100 |
| COMP | Cartilage oligomeric matrix protein | 0.0016 | 0.015 (0.013, 0.017) | 2.70E-51 |
| CPA1 | Carboxypeptidase A1 | 9.38E-08 | 0.006 (0.004, 0.008) | 6.90E-09 |

| | | | | |
|---|---|---|---|---|
| CPM | Carboxypeptidase M | 0.0002 | 0.032 (0.03, 0.034) | 1.50E-241 |
| CRISP2 | Cysteine-rich secretory protein 2 | 0.0016 | 0.025 (0.023, 0.027) | 5.90E-157 |
| CTBS | Di-N-acetylchitobiase | 0.0007 | 0.021 (0.02, 0.023) | 1.80E-109 |
| CTHRC1 | Collagen triple helix repeat-containing protein 1 | 2.11E-06 | 0.028 (0.026, 0.03) | 6.80E-182 |
| CTSF | Cathepsin F | -0.0003 | -0.01 (-0.012, -0.009) | 8.10E-28 |
| CTSL | Cathepsin L1 | 0.0003 | 0.014 (0.012, 0.016) | 2.70E-48 |
| CTSV | Cathepsin L2 | -0.0004 | -0.008 (-0.01, -0.006) | 1.60E-15 |
| CXCL17 | C-X-C motif chemokine 17 | 0.0006 | 0.008 (0.006, 0.01) | 1.10E-15 |
| DCC | Netrin receptor DCC | -1.78E-05 | -0.006 (-0.008, -0.005) | 1.20E-11 |
| DEFB4A_DEFB4B | Beta-defensin 4A | 1.71E-06 | 0.025 (0.024, 0.027) | 6.90E-160 |
| DIPK2B | Divergent protein kinase domain 2B | -0.0011 | -0.018 (-0.02, -0.016) | 2.30E-81 |
| DLK1 | Protein delta homolog 1 | 0.0006 | 0.013 (0.011, 0.015) | 6.80E-40 |
| DMP1 | Dentin matrix acidic phosphoprotein 1 | -0.0004 | -0.017 (-0.019, -0.015) | 2.80E-67 |
| DPT | Dermatopontin | 0.0043 | 0.016 (0.014, 0.018) | 2.30E-61 |
| DSG2 | Desmoglein-2 | -0.0018 | -0.029 (-0.031, -0.027) | 1.60E-204 |
| DSG4 | Desmoglein-4 | -0.0009 | -0.007 (-0.009, -0.005) | 2.50E-14 |
| DTX2 | Probable E3 ubiquitin-protein ligase DTX2 | -0.0005 | -0.006 (-0.008, -0.005) | 8.80E-12 |
| EGFLAM | Pikachurin | -0.0001 | -0.009 (-0.011, -0.008) | 2.70E-23 |
| ENPP2 | Ectonucleotide pyrophosphatase/phosphodiesterase family member 2 | -0.0015 | -0.034 (-0.035, -0.032) | 1.80E-276 |
| ENPP6 | Glycerophosphocholine cholinephosphodiesterase ENPP6 | -0.0002 | -0.019 (-0.021, -0.018) | 4.40E-95 |
| EPHA1 | Ephrin type-A receptor 1 | 0.0004 | 0.032 (0.03, 0.034) | 1.30E-245 |
| ERBB2 | Receptor tyrosine-protein kinase erbB-2 | 0.0031 | 0.033 (0.032, 0.035) | 8.00E-280 |
| ERI1 | 3'-5' exoribonuclease 1 | 6.89E-06 | 0.013 (0.011, 0.015) | 9.20E-44 |
| FASLG | Tumor necrosis factor ligand superfamily member 6 | -0.0009 | -0.017 (-0.019, -0.015) | 3.40E-68 |
| FBN2 | Fibrillin-2 | -0.0004 | -0.004 (-0.006, -0.002) | 0.0001 |
| FCGR3B | Low affinity immunoglobulin gamma Fc region receptor III-B | -0.0005 | -0.008 (-0.01, -0.006) | 1.00E-18 |
| FGF19 | Fibroblast growth factor 19 | -0.0003 | -0.007 (-0.009, -0.005) | 3.00E-12 |
| FGF21 | Fibroblast growth factor 21 | 0.0014 | 0.022 (0.021, 0.024) | 6.10E-123 |
| FGL1 | Fibrinogen-like protein 1 | -0.0006 | -0.025 (-0.027, -0.024) | 6.90E-159 |
| FN1 | Fibronectin | 0.0003 | 0.022 (0.02, 0.023) | 1.00E-117 |
| FOLR1 | Folate receptor alpha | -0.0013 | -0.02 (-0.022, -0.018) | 5.10E-90 |
| FSHB | Follitropin subunit beta | -0.0014 | -0.029 (-0.031, -0.028) | 8.40E-216 |
| GAGE2A | G antigen 2A | -0.0001 | -0.001 (-0.003, 0) | 0.1327 |
| GDF15 | Growth/differentiation factor 15 | 0.0013 | 0.013 (0.011, 0.015) | 7.70E-36 |
| GH1 | Somatotropin | -0.0007 | -0.03 (-0.031, -0.028) | 4.70E-244 |
| GHR | Growth hormone receptor | 0.0001 | 0.021 (0.019, 0.023) | 1.70E-104 |
| GHRL | Appetite-regulating hormone | -0.0008 | -0.022 (-0.023, -0.02) | 1.50E-112 |
| GLA | Alpha-galactosidase A | 0.0003 | 0.027 (0.025, 0.029) | 7.50E-178 |
| GNGT1 | Guanine nucleotide-binding protein G(T) subunit gamma-T1 | -0.0002 | -0.005 (-0.007, -0.003) | 2.10E-08 |
| GPD1 | Glycerol-3-phosphate dehydrogenase | 0.0004 | 0.022 (0.02, 0.024) | 8.40E-119 |
| GPR158 | Probable G-protein coupled receptor 158 | -0.0004 | -0.013 (-0.015, -0.011) | 9.90E-41 |
| GPR15L | Protein GPR15L | 0.0009 | 0.02 (0.018, 0.021) | 1.00E-94 |

| | | | | |
|---|---|---|---|---|
| GUSB | Beta-glucuronidase | 0.0011 | 0.038 (0.036, 0.04) | 0.00E+00 |
| HGF | Hepatocyte growth factor | 0.0005 | 0.026 (0.025, 0.028) | 3.30E-162 |
| HLA-DRA | HLA class II histocompatibility antigen, DR alpha chain | -0.0003 | -0.002 (-0.003, 0) | 0.0965 |
| HSPB6 | Heat shock protein beta-6 | 0.0038 | 0.03 (0.028, 0.031) | 8.10E-211 |
| HTR1B | 5-hydroxytryptamine receptor 1B | 0.0004 | 0 (-0.002, 0.002) | 0.8244 |
| IGDCC4 | Immunoglobulin superfamily DCC subclass member 4 | -0.0008 | -0.001 (-0.002, 0.001) | 0.5862 |
| IGFBP1 | Insulin-like growth factor-binding protein 1 | -0.0025 | -0.03 (-0.032, -0.029) | 2.00E-236 |
| IGFBP3 | Insulin-like growth factor-binding protein 3 | -0.0016 | -0.007 (-0.009, -0.005) | 2.60E-13 |
| IGSF21 | Immunoglobulin superfamily member 21 | 0.0001 | 0.016 (0.014, 0.018) | 2.70E-63 |
| IL19 | Interleukin-19 | 0.0001 | 0.013 (0.011, 0.015) | 1.30E-37 |
| IL32 | Interleukin-32 | -0.0007 | -0.014 (-0.016, -0.012) | 1.70E-48 |
| IL3RA | Interleukin-3 receptor subunit alpha | 2.52E-05 | 0.015 (0.014, 0.017) | 9.80E-58 |
| INSL5 | Insulin-like peptide INSL5 | 0.0005 | 0.022 (0.02, 0.024) | 9.70E-125 |
| ITIH3 | Inter-alpha-trypsin inhibitor heavy chain H3 | -0.0007 | -0.025 (-0.026, -0.023) | 4.70E-157 |
| KIR3DL2 | Killer cell immunoglobulin-like receptor 3DL2 | -0.0004 | -0.004 (-0.006, -0.002) | 1.10E-05 |
| KLK14 | Kallikrein-14 | -0.0029 | -0.017 (-0.019, -0.015) | 1.60E-68 |
| KLK3 | Prostate-specific antigen | 0.0003 | 0.046 (0.044, 0.047) | 0.00E+00 |
| LACRT | Extracellular glycoprotein lacritin | -0.0001 | -0.012 (-0.014, -0.01) | 1.60E-38 |
| LCAT | Phosphatidylcholine-sterol acyltransferase | 0.0011 | 0.022 (0.02, 0.024) | 2.80E-117 |
| LDLR | Low-density lipoprotein receptor | 0.001 | 0.026 (0.024, 0.028) | 6.20E-175 |
| LEFTY2 | Left-right determination factor 2 | -0.0005 | -0.019 (-0.021, -0.017) | 3.10E-94 |
| LPL | Lipoprotein lipase | -0.0032 | -0.037 (-0.038, -0.035) | 0.00E+00 |
| LPO | Lactoperoxidase | -0.0001 | -0.003 (-0.005, -0.001) | 0.0013 |
| LRTM2 | Leucine-rich repeat and transmembrane domain-containing protein 2 | -0.0006 | -0.021 (-0.023, -0.019) | 1.30E-109 |
| LUZP2 | Leucine zipper protein 2 | -2.71E-05 | -0.012 (-0.013, -0.01) | 1.70E-33 |
| MAN2B2 | Epididymis-specific alpha-mannosidase | 0.0006 | 0.018 (0.017, 0.02) | 1.40E-79 |
| MDGA1 | MAM domain-containing glycosylphosphatidylinositol anchor protein 1 | -0.0001 | -0.013 (-0.015, -0.011) | 5.50E-39 |
| MEP1A | Meprin A subunit alpha | 0.0006 | 0.019 (0.017, 0.021) | 1.10E-88 |
| MET | Hepatocyte growth factor receptor | -0.0016 | -0.018 (-0.02, -0.016) | 8.00E-80 |
| MMP3 | Stromelysin-1 | 0.0034 | 0.035 (0.033, 0.037) | 2.40E-307 |
| MRC1 | Macrophage mannose receptor 1 | 3.56E-06 | 0.013 (0.011, 0.015) | 9.80E-40 |
| MSMB | Beta-microseminoprotein | 0.001 | 0.018 (0.017, 0.02) | 8.70E-79 |
| NCAN | Neurocan core protein | -0.0005 | -0.025 (-0.027, -0.023) | 1.80E-152 |
| NPC2 | NPC intracellular cholesterol transporter 2 | 0.0002 | 0.019 (0.017, 0.021) | 4.80E-74 |
| NPTX2 | Neuronal pentraxin-2 | 0.0016 | 0.025 (0.024, 0.027) | 5.30E-160 |
| NTF3 | Neurotrophin-3 | -0.0013 | -0.01 (-0.012, -0.008) | 1.70E-26 |
| NTRK2 | BDNF/NT-3 growth factors receptor | 0.0007 | 0.002 (0, 0.004) | 0.0157 |
| OPTC | Opticin | -0.0015 | -0.02 (-0.022, -0.018) | 9.80E-100 |
| PCDH9 | Protocadherin-9 | -0.0003 | 0.002 (0, 0.004) | 0.0665 |
| PGA4 | Pepsin A-4 | 0.0003 | 0.007 (0.005, 0.009) | 8.00E-12 |
| PGF | Placenta growth factor | 0.0024 | 0.029 (0.028, 0.031) | 2.60E-193 |
| PI16 | Peptidase inhibitor 16 | -0.0001 | -0.012 (-0.014, -0.01) | 4.80E-33 |

| | | | | |
|---|---|---|---|---|
| PI3 | Elafin | 0.0003 | 0.022 (0.02, 0.024) | 1.60E-115 |
| PLA2G1B | Phospholipase A2 | -0.0021 | -0.022 (-0.024, -0.02) | 2.90E-114 |
| PLA2G7 | Platelet-activating factor acetylhydrolase | 0.0003 | 0.027 (0.026, 0.029) | 2.70E-182 |
| PLAT | Tissue-type plasminogen activator | 0.0023 | 0.033 (0.031, 0.035) | 9.90E-264 |
| POF1B | Protein POF1B | -0.0004 | -0.004 (-0.006, -0.003) | 4.40E-06 |
| PRAP1 | Proline-rich acidic protein 1 | 0.0008 | 0.04 (0.038, 0.042) | 0.00E+00 |
| PRL | Prolactin | -4.99E-05 | -0.014 (-0.015, -0.012) | 1.10E-46 |
| PROK1 | Prokineticin-1 | 0.001 | 0.031 (0.029, 0.033) | 1.80E-257 |
| PRSS8 | Prostasin | 0.0031 | 0.032 (0.03, 0.034) | 1.40E-223 |
| PSPN | Persephin | 0.0052 | 0.041 (0.039, 0.043) | 0.00E+00 |
| PTH | Parathyroid hormone | 0.0002 | 0.02 (0.018, 0.022) | 4.10E-93 |
| PTPRF | Receptor-type tyrosine-protein phosphatase F | 0.0001 | 0.02 (0.018, 0.022) | 2.10E-96 |
| PTPRR | Receptor-type tyrosine-protein phosphatase R | -0.0001 | -0.009 (-0.011, -0.007) | 1.90E-19 |
| PTX3 | Pentraxin-related protein PTX3 | -0.002 | -0.014 (-0.016, -0.012) | 5.80E-49 |
| PZP | Pregnancy zone protein | -0.0019 | -0.03 (-0.032, -0.029) | 3.10E-244 |
| RBP7 | Retinoid-binding protein 7 | -0.0004 | 0.001 (0, 0.003) | 0.1294 |
| REN | Renin | 0.0017 | 0.024 (0.021, 0.026) | 5.10E-94 |
| RTN4R | Reticulon-4 receptor | 1.96E-05 | 0.028 (0.026, 0.03) | 2.80E-177 |
| SCGB1A1 | Uteroglobin | -0.0002 | -0.002 (-0.004, 0) | 0.0703 |
| SCGB3A1 | Secretoglobin family 3A member 1 | -1.76E-05 | -0.016 (-0.017, -0.014) | 1.10E-59 |
| SCGB3A2 | Secretoglobin family 3A member 2 | -0.0009 | -0.018 (-0.02, -0.016) | 1.00E-80 |
| SCRG1 | Scrapie-responsive protein 1 | 0.0003 | 0.024 (0.023, 0.026) | 1.40E-136 |
| SELE | E-selectin | 0.0021 | 0.025 (0.023, 0.027) | 2.90E-143 |
| SEMA3F | Semaphorin-3F | 0.0012 | 0.023 (0.021, 0.024) | 1.70E-116 |
| SERPINA11 | Serpin A11 | -0.0001 | -0.028 (-0.03, -0.026) | 2.60E-201 |
| SERPINA6 | Corticosteroid-binding globulin | -0.0014 | -0.025 (-0.026, -0.023) | 2.40E-150 |
| SETMAR | Histone-lysine N-methyltransferase SETMAR | 0.0016 | 0.027 (0.025, 0.029) | 1.20E-180 |
| SEZ6 | Seizure protein 6 homolog | -0.0001 | -0.015 (-0.017, -0.013) | 2.10E-53 |
| SGSH | N-sulphoglucosamine sulphohydrolase | 0.0002 | 0.017 (0.015, 0.019) | 6.00E-68 |
| SIGLEC7 | Sialic acid-binding Ig-like lectin 7 | 0.0001 | 0.014 (0.012, 0.016) | 1.30E-43 |
| SLC39A14 | Zinc transporter ZIP14 | -0.0002 | -0.011 (-0.013, -0.009) | 5.70E-31 |
| SLITRK1 | SLIT and NTRK-like protein 1 | -0.0015 | -0.021 (-0.023, -0.02) | 5.50E-113 |
| SPESP1 | Sperm equatorial segment protein 1 | 0.0002 | 0.023 (0.022, 0.025) | 2.00E-136 |
| SPINK6 | Serine protease inhibitor Kazal-type 6 | 0.0002 | 0.022 (0.02, 0.024) | 2.30E-118 |
| SPRR3 | Small proline-rich protein 3 | -0.0001 | -0.016 (-0.018, -0.014) | 6.50E-63 |
| SRPX | Sushi repeat-containing protein SRPX | -0.0009 | -0.019 (-0.021, -0.017) | 8.20E-84 |
| SSC4D | Scavenger receptor cysteine-rich domain-containing group B protein | 0.0026 | 0.04 (0.038, 0.042) | 0.00E+00 |
| SSC5D | Soluble scavenger receptor cysteine-rich domain-containing protein SSC5D | 0.001 | 0.025 (0.023, 0.027) | 3.40E-147 |
| STAB2 | Stabilin-2 | 0.0002 | 0.025 (0.023, 0.027) | 2.10E-151 |
| TEX101 | Testis-expressed protein 101 | 0.0021 | 0.036 (0.035, 0.038) | 0.00E+00 |
| TFPI2 | Tissue factor pathway inhibitor 2 | -0.0004 | -0.012 (-0.014, -0.01) | 1.80E-35 |
| THBD | Thrombomodulin | 0.0019 | 0.018 (0.017, 0.02) | 4.80E-79 |

| | | | | |
|---|---|---|---|---|
| THBS2 | Thrombospondin-2 | -0.0003 | -0.011 (-0.013, -0.009) | 2.10E-30 |
| TNR | Tenascin-R | -0.0002 | -0.016 (-0.017, -0.014) | 4.20E-60 |
| TPT1 | Translationally-controlled tumor protein | -0.0001 | -0.002 (-0.003, 0) | 0.1116 |
| UPB1 | Beta-ureidopropionase | 0.0001 | 0.029 (0.027, 0.031) | 4.70E-211 |
| VEGFD | Vascular endothelial growth factor D | -0.0017 | -0.023 (-0.025, -0.021) | 1.60E-128 |
| VIT | Vitrin | 0.0004 | 0.031 (0.029, 0.032) | 9.20E-241 |
| VWC2L | von Willebrand factor C domain-containing protein 2-like | -0.0006 | -0.019 (-0.021, -0.018) | 2.70E-88 |
| WFDC12 | WAP four-disulfide core domain protein 12 | 0.0023 | 0.029 (0.027, 0.03) | 1.50E-201 |
| WIF1 | Wnt inhibitory factor 1 | -0.0027 | -0.017 (-0.019, -0.015) | 5.30E-70 |
| XG | Glycoprotein Xg | -0.0043 | -0.029 (-0.031, -0.027) | 7.40E-199 |
| ZHX2 | Zinc fingers and homeoboxes protein 2 | 2.77E-05 | 0.015 (0.013, 0.017) | 1.80E-59 |

CI: confidence interval.
*Linear regressions of the measured waist-hip ratio regressed on each of the proteins (per standard deviation).

**Supplemental Table 2D. The 25 LASSO Selected Proteins Shared across Obesity-related Phenotypes.**

| Protein | Name | BMI | | | Body Fat Percentage | | | Waist-hip Ratio | | |
|---|---|---|---|---|---|---|---|---|---|---|
| | | LASSO Beta | Linear Regression* | | LASSO Beta | Linear Regression* | | LASSO Beta | Linear Regression* | |
| | | | Beta (95% CI) | P | | Beta (95% CI) | P | | Beta (95% CI) | P |
| ADGRG2 | Adhesion G-protein coupled receptor G2 | -0.1181 | -0.97 (-1.06, -0.88) | 3.60E-92 | -0.0482 | 0.69 (0.5, 0.88) | 5.40E-13 | -0.003 | -0.029 (-0.031, -0.027) | 9.20E-211 |
| AGER | Advanced glycosylation end product-specific receptor | -0.1281 | -0.84 (-0.93, -0.74) | 1.60E-68 | -0.0398 | -0.24 (-0.43, -0.05) | 0.0136 | -0.0004 | -0.014 (-0.016, -0.012) | 8.90E-50 |
| BAG3 | BAG family molecular chaperone regulator 3 | 0.0124 | 1.1 (1, 1.19) | 1.40E-115 | -0.0041 | 0.49 (0.3, 0.68) | 5.60E-07 | 0.0001 | 0.017 (0.015, 0.018) | 4.80E-65 |
| CD300LG | CMRF35-like molecule 9 | 0.1124 | -0.01 (-0.11, 0.08) | 0.7624 | 0.1262 | 1.46 (1.27, 1.65) | 2.30E-51 | -0.0056 | -0.023 (-0.025, -0.021) | 5.20E-130 |
| CD99 | CD99 antigen | 0.0552 | 0.66 (0.56, 0.75) | 8.90E-42 | -0.1061 | -2 (-2.19, -1.82) | 4.90E-97 | 0.0045 | 0.03 (0.028, 0.031) | 1.20E-214 |
| CFH | Complement factor H | 0.1082 | 1.84 (1.75, 1.93) | 9.9e-324 | 0.0384 | 2.4 (2.22, 2.59) | 2.30E-132 | 0.0005 | 0.026 (0.024, 0.028) | 7.40E-154 |
| CHGB | Secretogranin-1 | -0.0551 | -0.97 (-1.06, -0.87) | 2.00E-87 | -0.0457 | -0.98 (-1.17, -0.79) | 2.90E-23 | -0.001 | -0.009 (-0.011, -0.007) | 7.70E-21 |
| COL4A1 | Collagen alpha-1(IV) chain | -0.1903 | -1.27 (-1.37, -1.18) | 1.10E-157 | -0.2023 | -1.46 (-1.65, -1.27) | 1.30E-51 | -0.002 | -0.02 (-0.022, -0.019) | 3.30E-100 |
| CTBS | Di-N-acetylchitobiase | -0.002 | 0.68 (0.58, 0.77) | 2.10E-44 | -0.083 | -0.05 (-0.25, 0.14) | 0.5753 | 0.0007 | 0.021 (0.02, 0.023) | 1.80E-109 |
| CTHRC1 | Collagen triple helix repeat-containing protein 1 | 0.1693 | 1.2 (1.11, 1.29) | 7.00E-134 | -0.0993 | -0.83 (-1.02, -0.64) | 4.40E-17 | 2.11E-06 | 0.028 (0.026, 0.03) | 6.80E-182 |
| DMP1 | Dentin matrix acidic phosphoprotein 1 | -0.0295 | -0.66 (-0.75, -0.56) | 6.20E-42 | 0.0545 | -0.03 (-0.22, 0.16) | 0.7496 | -0.0004 | -0.017 (-0.019, -0.015) | 2.80E-67 |
| DPT | Dermatopontin | 0.0935 | 1.35 (1.26, 1.44) | 1.40E-172 | 0.1813 | 2.08 (1.89, 2.27) | 5.50E-102 | 0.0043 | 0.016 (0.014, 0.018) | 2.30E-61 |
| ENPP6 | Glycerophosphocholine cholinephosphodiesterase ENPP6 | -0.047 | -1.15 (-1.24, -1.07) | 1.40E-136 | -0.1154 | -0.74 (-0.92, -0.55) | 5.60E-15 | -0.0002 | -0.019 (-0.021, -0.018) | 4.40E-95 |
| FGL1 | Fibrinogen-like protein 1 | -0.0523 | -0.63 (-0.73, -0.54) | 3.00E-40 | -0.0181 | 1.53 (1.35, 1.72) | 2.20E-58 | -0.0006 | -0.025 (-0.027, -0.024) | 6.90E-159 |
| GHRL | Appetite-regulating hormone | -0.0668 | -1.02 (-1.11, -0.93) | 6.90E-101 | -0.0548 | 0.34 (0.15, 0.53) | 0.0004 | -0.0008 | -0.022 (-0.023, -0.02) | 1.50E-112 |
| GPD1 | Glycerol-3-phosphate dehydrogenase | 0.0752 | 1.67 (1.58, 1.75) | 4.70E-284 | 0.0394 | 2.27 (2.08, 2.45) | 2.90E-127 | 0.0004 | 0.022 (0.02, 0.024) | 8.40E-119 |
| NPTX2 | Neuronal pentraxin-2 | -0.0024 | -0.2 (-0.29, -0.1) | 3.40E-05 | -0.1173 | -3.25 (-3.42, -3.07) | 3.40E-271 | 0.0016 | 0.025 (0.024, 0.027) | 5.30E-160 |
| NTRK2 | BDNF/NT-3 growth factors receptor | 0.1347 | 0.57 (0.48, 0.66) | 2.50E-32 | 0.0757 | 1.25 (1.06, 1.44) | 7.20E-39 | 0.0007 | 0.002 (0, 0.004) | 0.0157 |
| OPTC | Opticin | -0.2481 | -0.91 (-1, -0.82) | 2.00E-81 | -0.0097 | 0.56 (0.37, 0.75) | 5.40E-09 | -0.0015 | -0.02 (-0.022, -0.018) | 9.80E-100 |
| PLA2G1B | Phospholipase A2 | -0.0326 | -0.98 (-1.07, -0.89) | 1.20E-90 | -0.0466 | 0.02 (-0.17, 0.21) | 0.8165 | -0.0021 | -0.022 (-0.024, -0.02) | 2.90E-114 |
| SCGB3A1 | Secretoglobin family 3A member 1 | -0.0528 | -0.78 (-0.87, -0.69) | 2.70E-60 | -0.0767 | 0.13 (-0.06, 0.32) | 0.176 | -1.76E-05 | -0.016 (-0.017, -0.014) | 1.10E-59 |
| SCGB3A2 | Secretoglobin family 3A member 2 | -0.073 | -1.22 (-1.31, -1.13) | 1.80E-146 | -0.0432 | -0.92 (-1.1, -0.73) | 1.50E-21 | -0.0009 | -0.018 (-0.02, -0.016) | 1.00E-80 |
| SPINK6 | Serine protease inhibitor Kazal-type 6 | 0.006 | 0.79 (0.7, 0.88) | 3.90E-62 | -0.1922 | -0.54 (-0.73, -0.35) | 1.40E-08 | 0.0002 | 0.022 (0.02, 0.024) | 2.30E-118 |

| | | | | | | | | | | |
|---|---|---|---|---|---|---|---|---|---|---|
| SSC4D | Scavenger receptor cysteine-rich domain-containing group B protein | 0.0317 | 1.75 (1.66, 1.83) | 5.9e-312 | -0.0092 | 0.02 (-0.17, 0.21) | 0.8352 | 0.0026 | 0.04 (0.038, 0.042) | 0.00E+00 |
| VWC2L | von Willebrand factor C domain-containing protein 2-like | -0.1231 | -1.03 (-1.13, -0.94) | 4.90E-100 | -0.0068 | 0.23 (0.04, 0.42) | 0.019 | -0.0006 | -0.019 (-0.021, -0.018) | 2.70E-88 |

CI: confidence interval.

*Linear regressions of the measured obesity-related phenotype regressed on each of the proteins (per standard deviation).

**Supplemental Table 3A. Pathway Enrichment for Gene Ontology Using the 389 LASSO-selected Proteins for BMI. Top 10 Pathways out of Total 86 Pathways with $p < 0.05$ are shown.**

| GO.ID | Term | Annotated | Significant | Expected | P | FDR |
|---|---|---|---|---|---|---|
| GO:0007155 | cell adhesion | 581 | 103 | 77.16 | 0.0005 | 1 |
| GO:0006508 | proteolysis | 400 | 58 | 53.12 | 0.0005 | 1 |
| GO:0051965 | positive regulation of synapse assembly | 22 | 9 | 2.92 | 0.0012 | 1 |
| GO:0032696 | negative regulation of interleukin-13 production | 5 | 4 | 0.66 | 0.0014 | 1 |
| GO:0051838 | cytolysis by host of symbiont cells | 5 | 4 | 0.66 | 0.0014 | 1 |
| GO:0014009 | glial cell proliferation | 23 | 5 | 3.05 | 0.0023 | 1 |
| GO:0099560 | synaptic membrane adhesion | 9 | 5 | 1.2 | 0.0032 | 1 |
| GO:0031214 | biomineral tissue development | 47 | 11 | 6.24 | 0.0036 | 1 |
| GO:0010460 | positive regulation of heart rate | 6 | 4 | 0.8 | 0.0037 | 1 |
| GO:0071310 | cellular response to organic substance | 541 | 75 | 71.85 | 0.0083 | 1 |

**Supplemental Table 3B. Pathway Enrichment for Gene Ontology Using the 385 LASSO-selected Proteins for Body Fat Percentage. Top 10 Pathways out of Total 75 Pathways with p < 0.05 are shown.**

| GO.ID | Term | Annotated | Significant | Expected | P | FDR |
|---|---|---|---|---|---|---|
| GO:0007155 | cell adhesion | 581 | 100 | 76.95 | 3.50E-07 | 0.0039 |
| GO:0030198 | extracellular matrix organization | 123 | 37 | 16.29 | 9.00E-06 | 0.0507 |
| GO:0006508 | proteolysis | 400 | 64 | 52.98 | 6.20E-05 | 0.2328 |
| GO:0032526 | response to retinoic acid | 27 | 9 | 3.58 | 0.0007 | 1 |
| GO:0038063 | collagen-activated tyrosine kinase receptor signaling pathway | 3 | 3 | 0.4 | 0.0023 | 1 |
| GO:0042104 | positive regulation of activated T cell proliferation | 12 | 6 | 1.59 | 0.0024 | 1 |
| GO:0006954 | inflammatory response | 329 | 57 | 43.57 | 0.0040 | 1 |
| GO:0051603 | proteolysis involved in protein catabolic process | 117 | 10 | 15.5 | 0.0070 | 1 |
| GO:0043129 | surfactant homeostasis | 7 | 4 | 0.93 | 0.0076 | 1 |
| GO:0014012 | peripheral nervous system axon regeneration | 4 | 3 | 0.53 | 0.0083 | 1 |

**Supplemental Table 3C. Pathway Enrichment for Gene Ontology Using the 176 LASSO-selected Proteins for Waist-hip Ratio. Top 10 Pathways out of Total 57 Pathways with p < 0.05 are shown.**

| GO.ID | Term | Annotated | Significant | Expected | P | FDR |
|---|---|---|---|---|---|---|
| GO:0007155 | cell adhesion | 581 | 49 | 35.22 | 0.0003 | 1 |
| GO:0046326 | positive regulation of glucose import | 8 | 4 | 0.49 | 0.0008 | 1 |
| GO:0042756 | drinking behavior | 3 | 3 | 0.18 | 0.0036 | 1 |
| GO:0044871 | negative regulation by host of viral glycoprotein metabolic process | 2 | 2 | 0.12 | 0.0037 | 1 |
| GO:1903016 | negative regulation of exo-alpha-sialidase activity | 2 | 2 | 0.12 | 0.0037 | 1 |
| GO:0019695 | choline metabolic process | 2 | 2 | 0.12 | 0.0037 | 1 |
| GO:0090675 | intermicrovillar adhesion | 2 | 2 | 0.12 | 0.0037 | 1 |
| GO:0044869 | negative regulation by host of viral exo-alpha-sialidase activity | 2 | 2 | 0.12 | 0.0037 | 1 |
| GO:0050805 | negative regulation of synaptic transmission | 16 | 5 | 0.97 | 0.0037 | 1 |
| GO:0019886 | antigen processing and presentation of exogenous peptide antigen via MHC class II | 12 | 4 | 0.73 | 0.0044 | 1 |

**Supplemental Table 4. Associations Between Protein Predicted Scores and Outcomes. Sensitivity Analysis Results after Excluding Cancer at Baseline.**

| Outcome | Model | $PPS_{BMI}$ (per SD) | | | | $PPS_{BFP}$ (per SD) | | | | $PPS_{WHR}$ (per SD) | | | |
|---|---|---|---|---|---|---|---|---|---|---|---|---|---|
| | | N | N Event | HR (95% CI) | P | N | N Event | HR (95% CI) | P | N | N Event | HR (95% CI) | P |
| MACE | Model 1 | 29032 | 3586 | 1.22 (1.18, 1.26) | **<.0001** | 28604 | 3501 | 1.29 (1.23, 1.35) | **<.0001** | 29081 | 3597 | 1.46 (1.39, 1.54) | **<.0001** |
| | Model 2 | 29032 | 3586 | 1.15 (1.07, 1.23) | **0.0002** | 28604 | 3501 | 1.33 (1.22, 1.46) | **<.0001** | 29081 | 3597 | 1.33 (1.25, 1.42) | **<.0001** |
| | Model 3 | 23476 | 2897 | 1.05 (0.97, 1.14) | 0.229 | 23148 | 2837 | 1.21 (1.09, 1.33) | **0.0003** | 23513 | 2906 | 1.12 (1.03, 1.22) | **0.0099** |
| Ischemic Stroke | Model 1 | 29031 | 691 | 1.14 (1.05, 1.23) | **0.0016** | 28604 | 683 | 1.18 (1.06, 1.32) | **0.0027** | 29080 | 695 | 1.32 (1.16, 1.49) | **<.0001** |
| | Model 2 | 29031 | 691 | 1.07 (0.92, 1.26) | 0.3819 | 28604 | 683 | 1.05 (0.87, 1.28) | 0.5963 | 29080 | 695 | 1.26 (1.08, 1.47) | **0.0027** |
| | Model 3 | 23476 | 572 | 1.07 (0.89, 1.28) | 0.4926 | 23148 | 568 | 0.94 (0.75, 1.17) | 0.5474 | 23513 | 574 | 1.19 (0.97, 1.46) | 0.0935 |
| MI | Model 1 | 29032 | 2743 | 1.23 (1.19, 1.28) | **<.0001** | 28604 | 2679 | 1.31 (1.24, 1.38) | **<.0001** | 29081 | 2745 | 1.52 (1.43, 1.62) | **<.0001** |
| | Model 2 | 29032 | 2743 | 1.21 (1.12, 1.31) | **<.0001** | 28604 | 2679 | 1.37 (1.24, 1.52) | **<.0001** | 29081 | 2745 | 1.4 (1.29, 1.51) | **<.0001** |
| | Model 3 | 23476 | 2215 | 1.09 (1, 1.2) | 0.0649 | 23148 | 2167 | 1.24 (1.1, 1.39) | **0.0003** | 23513 | 2217 | 1.14 (1.03, 1.26) | **0.0086** |
| CV Death | Model 1 | 29032 | 828 | 1.3 (1.2, 1.41) | **<.0001** | 28604 | 789 | 1.44 (1.29, 1.62) | **<.0001** | 29081 | 834 | 1.43 (1.27, 1.62) | **<.0001** |
| | Model 2 | 29032 | 828 | 1.01 (0.86, 1.18) | 0.8971 | 28604 | 789 | 1.56 (1.27, 1.9) | **<.0001** | 29081 | 834 | 1.19 (1.03, 1.37) | **0.0192** |
| | Model 3 | 23476 | 662 | 0.97 (0.82, 1.16) | 0.7492 | 23148 | 633 | 1.39 (1.11, 1.75) | **0.0046** | 23513 | 667 | 1.09 (0.91, 1.31) | 0.3312 |

$PPS_{BMI}$, protein predicted score of BMI; $PPS_{BFP}$: protein predicted score of body fat percentage; $PPS_{WHR}$: protein predicted score of waist-hip ratio; MACE: major adverse cardiovascular events (ischemic stroke, myocardial infarction, and cardiovascular death); MI: myocardial infarction; CV: cardiovascular; HR: hazard ratio; CI: confidence interval.
Model 1: adjusted for age, sex, race (white vs. other);
Model 2: adjusted for the measured obesity-related phenotype (BMI, body fat percentage, or waist-hip ratio) in addition to Model 1;
Model 3: adjusted for total cholesterol, high density lipoprotein cholesterol, systolic blood pressure, estimated glomerular filtration rate calculated using the 2021 CKD-EPI equation, diabetes, current smoking, blood pressure lowering medication use, cholesterol lowering medication use in addition to Model 2.

**Supplemental Table 5. Sex-Specific Associations Between Protein Predicted Scores and Outcomes.**

| Protein Predicted Score | Outcome | Model | Male | | | | Female | | | | Sex Interaction P |
|---|---|---|---|---|---|---|---|---|---|---|---|
| | | | N | N Event | HR (95% CI) | P | N | N Event | HR (95% CI) | P | |
| PPS$_{BMI}$ (per SD) | MACE | Model 1 | 14927 | 2489 | 1.26 (1.21, 1.32) | <.0001 | 17830 | 1568 | 1.17 (1.11, 1.23) | <.0001 | 0.018 |
| | | Model 2 | 14927 | 2489 | 1.21 (1.11, 1.32) | <.0001 | 17830 | 1568 | 1.08 (0.97, 1.19) | 0.1721 | 0.0127 |
| | | Model 3 | 12141 | 2016 | 1.13 (1.02, 1.25) | 0.0159 | 14309 | 1255 | 1.01 (0.89, 1.14) | 0.8875 | 0.3764 |
| | Ischemic Stroke | Model 1 | 14927 | 427 | 1.13 (1.01, 1.26) | 0.0256 | 17828 | 349 | 1.14 (1.03, 1.27) | 0.0103 | 0.9263 |
| | | Model 2 | 14927 | 427 | 1.04 (0.84, 1.27) | 0.7402 | 17828 | 349 | 1.18 (0.94, 1.47) | 0.1469 | 0.9494 |
| | | Model 3 | 12141 | 351 | 1.04 (0.82, 1.32) | 0.7535 | 14309 | 289 | 1.17 (0.92, 1.5) | 0.2054 | 0.3422 |
| | MI | Model 1 | 14927 | 1984 | 1.27 (1.21, 1.33) | <.0001 | 17830 | 1109 | 1.19 (1.12, 1.26) | <.0001 | 0.0681 |
| | | Model 2 | 14927 | 1984 | 1.28 (1.16, 1.41) | <.0001 | 17830 | 1109 | 1.1 (0.97, 1.25) | 0.1322 | 0.0612 |
| | | Model 3 | 12141 | 1610 | 1.17 (1.04, 1.31) | 0.007 | 14309 | 886 | 1.03 (0.89, 1.19) | 0.7113 | 0.4282 |
| | CV Death | Model 1 | 14927 | 622 | 1.33 (1.2, 1.46) | <.0001 | 17830 | 348 | 1.21 (1.08, 1.36) | 0.0011 | 0.2363 |
| | | Model 2 | 14927 | 622 | 1.17 (0.97, 1.41) | 0.1047 | 17830 | 348 | 0.86 (0.68, 1.09) | 0.222 | 0.1578 |
| | | Model 3 | 12141 | 481 | 1.17 (0.95, 1.43) | 0.1421 | 14309 | 286 | 0.84 (0.64, 1.09) | 0.1836 | 0.7397 |
| PPS$_{BFP}$ (per SD) | MACE | Model 1 | 14692 | 2428 | 1.35 (1.27, 1.43) | <.0001 | 17576 | 1524 | 1.21 (1.13, 1.3) | <.0001 | 0.0457 |
| | | Model 2 | 14692 | 2428 | 1.36 (1.22, 1.51) | <.0001 | 17576 | 1524 | 1.33 (1.15, 1.53) | <.0001 | 0.0548 |
| | | Model 3 | 11954 | 1971 | 1.26 (1.12, 1.42) | 0.0001 | 14120 | 1225 | 1.25 (1.06, 1.46) | 0.0069 | 0.3887 |
| | Ischemic Stroke | Model 1 | 14692 | 423 | 1.24 (1.07, 1.43) | 0.0038 | 17575 | 344 | 1.14 (0.98, 1.33) | 0.0836 | 0.5298 |
| | | Model 2 | 14692 | 423 | 1.13 (0.89, 1.45) | 0.319 | 17575 | 344 | 1.1 (0.83, 1.46) | 0.515 | 0.5023 |
| | | Model 3 | 11954 | 349 | 1.05 (0.79, 1.38) | 0.7474 | 14120 | 287 | 0.99 (0.73, 1.34) | 0.9405 | 0.8778 |
| | MI | Model 1 | 14692 | 1937 | 1.34 (1.26, 1.43) | <.0001 | 17576 | 1078 | 1.26 (1.16, 1.37) | <.0001 | 0.3371 |
| | | Model 2 | 14692 | 1937 | 1.4 (1.24, 1.57) | <.0001 | 17576 | 1078 | 1.35 (1.15, 1.59) | 0.0003 | 0.3805 |
| | | Model 3 | 11954 | 1574 | 1.28 (1.12, 1.46) | 0.0003 | 14120 | 864 | 1.28 (1.07, 1.55) | 0.0087 | 0.7546 |
| | CV Death | Model 1 | 14692 | 591 | 1.53 (1.34, 1.75) | <.0001 | 17576 | 333 | 1.19 (1, 1.42) | 0.0462 | 0.0344 |
| | | Model 2 | 14692 | 591 | 1.69 (1.37, 2.1) | <.0001 | 17576 | 333 | 1.24 (0.88, 1.74) | 0.2285 | 0.04 |
| | | Model 3 | 11954 | 457 | 1.58 (1.23, 2.02) | 0.0003 | 14120 | 275 | 1.16 (0.79, 1.71) | 0.4391 | 0.324 |
| PPS$_{WHR}$ (per SD) | MACE | Model 1 | 14966 | 2500 | 1.52 (1.42, 1.63) | <.0001 | 17850 | 1570 | 1.4 (1.29, 1.51) | <.0001 | 0.1787 |
| | | Model 2 | 14966 | 2500 | 1.34 (1.24, 1.46) | <.0001 | 17850 | 1570 | 1.32 (1.2, 1.46) | <.0001 | 0.146 |
| | | Model 3 | 12168 | 2026 | 1.15 (1.03, 1.27) | 0.0139 | 14327 | 1256 | 1.14 (1, 1.29) | 0.0476 | 0.4153 |
| | | Model 1 | 14966 | 430 | 1.32 (1.12, 1.55) | 0.0009 | 17848 | 351 | 1.31 (1.11, 1.55) | 0.0013 | 0.9464 |

| | | | | | | | | | | | |
|---|---|---|---|---|---|---|---|---|---|---|---|
| | **Ischemic Stroke** | **Model 2** | 14966 | 430 | 1.14 (0.94, 1.39) | 0.1805 | 17848 | 351 | 1.42 (1.15, 1.74) | **0.001** | 0.9644 |
| | | **Model 3** | 12168 | 353 | 1.08 (0.82, 1.41) | 0.5821 | 14327 | 290 | 1.34 (1.02, 1.75) | **0.0345** | 0.6546 |
| | **MI** | **Model 1** | 14966 | 1986 | 1.55 (1.43, 1.67) | **<.0001** | 17850 | 1109 | 1.48 (1.36, 1.62) | **<.0001** | 0.6109 |
| | | **Model 2** | 14966 | 1986 | 1.42 (1.29, 1.56) | **<.0001** | 17850 | 1109 | 1.38 (1.23, 1.54) | **<.0001** | 0.5559 |
| | | **Model 3** | 12168 | 1612 | 1.16 (1.03, 1.3) | **0.0165** | 14327 | 886 | 1.17 (1.01, 1.35) | **0.0413** | 0.8301 |
| | **CV Death** | **Model 1** | 14966 | 630 | 1.4 (1.21, 1.61) | **<.0001** | 17850 | 348 | 1.45 (1.21, 1.74) | **<.0001** | 0.6127 |
| | | **Model 2** | 14966 | 630 | 1.08 (0.91, 1.27) | 0.3814 | 17850 | 348 | 1.31 (1.06, 1.63) | **0.0137** | 0.6991 |
| | | **Model 3** | 12168 | 488 | 1.06 (0.85, 1.31) | 0.6094 | 14327 | 286 | 1.12 (0.86, 1.48) | 0.4018 | 0.6826 |

$PPS_{BMI}$, protein predicted score of BMI; $PPS_{BFP}$: protein predicted score of body fat percentage; $PPS_{WHR}$: protein predicted score of waist-hip ratio; MACE: major adverse cardiovascular events (ischemic stroke, myocardial infarction, and cardiovascular death); MI: myocardial infarction; CV: cardiovascular; HR: hazard ratio; CI: confidence interval.

Model 1: adjusted for age, race (white vs. other);

Model 2: adjusted for the measured obesity-related phenotype (BMI, body fat percentage, or waist-hip ratio) in addition to Model 1;

Model 3: adjusted for total cholesterol, high density lipoprotein cholesterol, systolic blood pressure, estimated glomerular filtration rate calculated using the 2021 CKD-EPI equation, diabetes, current smoking, blood pressure lowering medication use, cholesterol lowering medication use in addition to Model 2.

**Supplemental Table 6. Sex-Specific Associations Between Protein Predicted Scores and Outcomes. Sensitivity Analysis Results after Excluding Cancer at Baseline.**

| Protein Predicted Score | Outcome | Model | Male | | | | Female | | | | Sex Interaction P |
|---|---|---|---|---|---|---|---|---|---|---|---|
| | | | N | N Event | HR (95% CI) | P | N | N Event | HR (95% CI) | P | |
| $PPS_{BMI}$ (per SD) | MACE | **Model 1** | 13738 | 2250 | 1.26 (1.2, 1.32) | **<.0001** | 15294 | 1336 | 1.17 (1.11, 1.23) | **<.0001** | **0.03** |
| | | **Model 2** | 13738 | 2250 | 1.19 (1.09, 1.31) | **0.0002** | 15294 | 1336 | 1.07 (0.96, 1.19) | 0.243 | **0.0208** |
| | | **Model 3** | 11175 | 1825 | 1.1 (0.99, 1.23) | 0.0754 | 12301 | 1072 | 0.98 (0.86, 1.12) | 0.7712 | 0.3409 |
| | Ischemic Stroke | **Model 1** | 13738 | 388 | 1.11 (0.99, 1.25) | 0.0662 | 15293 | 303 | 1.15 (1.03, 1.29) | **0.01** | 0.6829 |
| | | **Model 2** | 13738 | 388 | 0.96 (0.77, 1.19) | 0.7029 | 15293 | 303 | 1.21 (0.95, 1.54) | 0.1169 | 0.7228 |
| | | **Model 3** | 11175 | 323 | 0.95 (0.74, 1.22) | 0.691 | 12301 | 249 | 1.21 (0.92, 1.58) | 0.1669 | 0.2 |
| | MI | **Model 1** | 13738 | 1797 | 1.27 (1.21, 1.33) | **<.0001** | 15294 | 946 | 1.19 (1.12, 1.26) | **<.0001** | 0.0816 |
| | | **Model 2** | 13738 | 1797 | 1.28 (1.16, 1.42) | **<.0001** | 15294 | 946 | 1.09 (0.96, 1.25) | 0.1828 | 0.0743 |
| | | **Model 3** | 11175 | 1456 | 1.16 (1.03, 1.3) | **0.0146** | 12301 | 759 | 0.99 (0.85, 1.16) | 0.8958 | 0.3378 |
| | CV Death | **Model 1** | 13738 | 549 | 1.31 (1.18, 1.46) | **<.0001** | 15294 | 279 | 1.28 (1.13, 1.45) | **0.0001** | 0.7621 |
| | | **Model 2** | 13738 | 549 | 1.1 (0.9, 1.35) | 0.3272 | 15294 | 279 | 0.87 (0.68, 1.11) | 0.2641 | 0.5688 |
| | | **Model 3** | 11175 | 424 | 1.09 (0.88, 1.36) | 0.4196 | 12301 | 238 | 0.81 (0.61, 1.08) | 0.1507 | 0.9832 |
| $PPS_{BFP}$ (per SD) | MACE | **Model 1** | 13522 | 2199 | 1.35 (1.27, 1.43) | **<.0001** | 15082 | 1302 | 1.21 (1.13, 1.31) | **<.0001** | 0.0567 |
| | | **Model 2** | 13522 | 2199 | 1.34 (1.2, 1.5) | **<.0001** | 15082 | 1302 | 1.3 (1.12, 1.51) | **0.0006** | 0.0629 |
| | | **Model 3** | 11004 | 1788 | 1.24 (1.09, 1.41) | **0.0008** | 12144 | 1049 | 1.16 (0.98, 1.37) | 0.0825 | 0.2368 |
| | Ischemic Stroke | **Model 1** | 13522 | 384 | 1.22 (1.05, 1.42) | **0.0101** | 15082 | 299 | 1.15 (0.98, 1.34) | 0.0965 | 0.6012 |
| | | **Model 2** | 13522 | 384 | 1.06 (0.82, 1.38) | 0.6345 | 15082 | 299 | 1.03 (0.76, 1.4) | 0.8309 | 0.5513 |
| | | **Model 3** | 11004 | 321 | 0.98 (0.73, 1.31) | 0.8725 | 12144 | 247 | 0.9 (0.65, 1.26) | 0.5547 | 0.8359 |
| | MI | **Model 1** | 13522 | 1756 | 1.34 (1.25, 1.44) | **<.0001** | 15082 | 923 | 1.26 (1.16, 1.38) | **<.0001** | 0.3777 |
| | | **Model 2** | 13522 | 1756 | 1.4 (1.24, 1.59) | **<.0001** | 15082 | 923 | 1.3 (1.1, 1.55) | **0.0026** | 0.4093 |
| | | **Model 3** | 11004 | 1424 | 1.27 (1.1, 1.46) | **0.001** | 12144 | 743 | 1.19 (0.98, 1.45) | 0.0818 | 0.5597 |
| | CV Death | **Model 1** | 13522 | 523 | 1.55 (1.34, 1.78) | **<.0001** | 15082 | 266 | 1.28 (1.05, 1.56) | **0.0127** | 0.1531 |
| | | **Model 2** | 13522 | 523 | 1.72 (1.37, 2.15) | **<.0001** | 15082 | 266 | 1.26 (0.85, 1.87) | 0.2469 | 0.1656 |
| | | **Model 3** | 11004 | 405 | 1.6 (1.23, 2.08) | **0.0004** | 12144 | 228 | 1.06 (0.69, 1.63) | 0.7772 | 0.3781 |
| $PPS_{WHR}$ (per SD) | MACE | **Model 1** | 13770 | 2259 | 1.51 (1.4, 1.62) | **<.0001** | 15311 | 1338 | 1.4 (1.29, 1.52) | **<.0001** | 0.2402 |
| | | **Model 2** | 13770 | 2259 | 1.32 (1.21, 1.45) | **<.0001** | 15311 | 1338 | 1.34 (1.21, 1.49) | **<.0001** | 0.2034 |
| | | **Model 3** | 11197 | 1833 | 1.13 (1.01, 1.26) | **0.0375** | 12316 | 1073 | 1.1 (0.96, 1.26) | 0.1674 | 0.406 |

| | | | | | | | | | | | |
|---|---|---|---|---|---|---|---|---|---|---|---|
| | **Ischemic Stroke** | **Model 1** | 13770 | 390 | 1.29 (1.08, 1.53) | **0.0044** | 15310 | 305 | 1.35 (1.13, 1.61) | **0.0011** | 0.6877 |
| | | **Model 2** | 13770 | 390 | 1.08 (0.87, 1.33) | 0.4876 | 15310 | 305 | 1.52 (1.22, 1.9) | **0.0002** | 0.7021 |
| | | **Model 3** | 11197 | 324 | 1.03 (0.77, 1.38) | 0.8201 | 12316 | 250 | 1.37 (1.03, 1.82) | **0.0321** | 0.5148 |
| | **MI** | **Model 1** | 13770 | 1799 | 1.54 (1.42, 1.68) | **<.0001** | 15311 | 946 | 1.48 (1.34, 1.63) | **<.0001** | 0.5954 |
| | | **Model 2** | 13770 | 1799 | 1.41 (1.28, 1.55) | **<.0001** | 15311 | 946 | 1.36 (1.21, 1.54) | **<.0001** | 0.5448 |
| | | **Model 3** | 11197 | 1458 | 1.15 (1.01, 1.3) | **0.0315** | 12316 | 759 | 1.12 (0.95, 1.31) | 0.1733 | 0.7572 |
| | **CV Death** | **Model 1** | 13770 | 555 | 1.36 (1.17, 1.59) | **<.0001** | 15311 | 279 | 1.52 (1.25, 1.86) | **<.0001** | 0.2809 |
| | | **Model 2** | 13770 | 555 | 1.05 (0.88, 1.26) | 0.5901 | 15311 | 279 | 1.45 (1.14, 1.84) | **0.0023** | 0.3256 |
| | | **Model 3** | 11197 | 429 | 1.06 (0.85, 1.33) | 0.5969 | 12316 | 238 | 1.15 (0.86, 1.54) | 0.3532 | 0.4266 |

$PPS_{BMI}$, protein predicted score of BMI; $PPS_{BFP}$: protein predicted score of body fat percentage; $PPS_{WHR}$: protein predicted score of waist-hip ratio; MACE: major adverse cardiovascular events (ischemic stroke, myocardial infarction, and cardiovascular death); MI: myocardial infarction; CV: cardiovascular; HR: hazard ratio; CI: confidence interval.

Model 1: adjusted for age, race (white vs. other);

Model 2: adjusted for the measured obesity-related phenotype (BMI, body fat percentage, or waist-hip ratio) in addition to Model 1;

Model 3: adjusted for total cholesterol, high density lipoprotein cholesterol, systolic blood pressure, estimated glomerular filtration rate calculated using the 2021 CKD-EPI equation, diabetes, current smoking, blood pressure lowering medication use, cholesterol lowering medication use in addition to Model 2.

**Supplemental Figure 1. Description of the sample selection workflow from the UK Biobank cohort.**

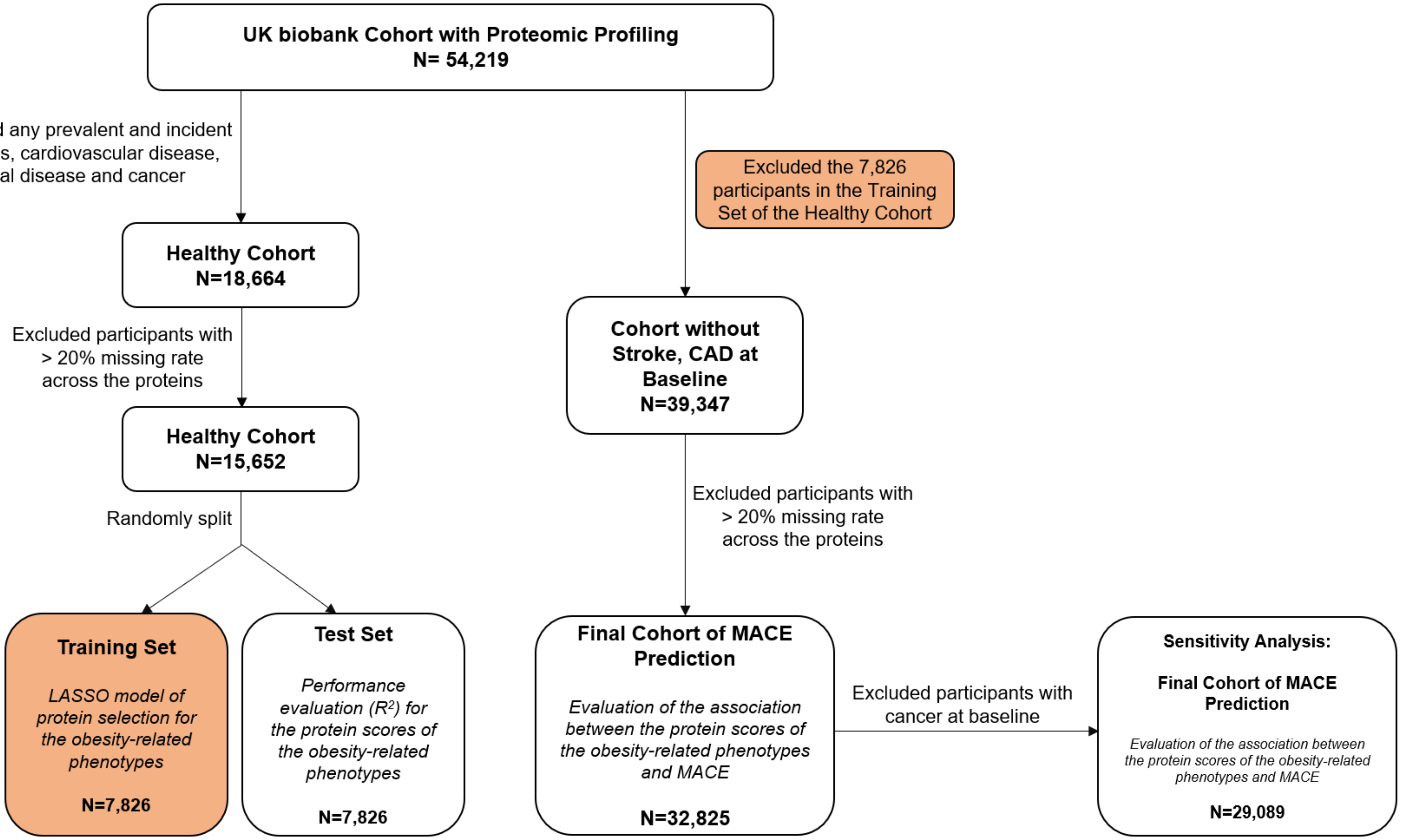

**Supplemental Figure 2. $R^2$ values assessing the prediction performance of protein-predicted scores of BMI ($PPS_{BMI}$), BFP ($PPS_{BFP}$), and WHR ($PPS_{WHR}$) across various sample sizes. The median $R^2$ from the LASSO models with 2.5% and 97.5% percentiles of the 100 iterations are shown.**

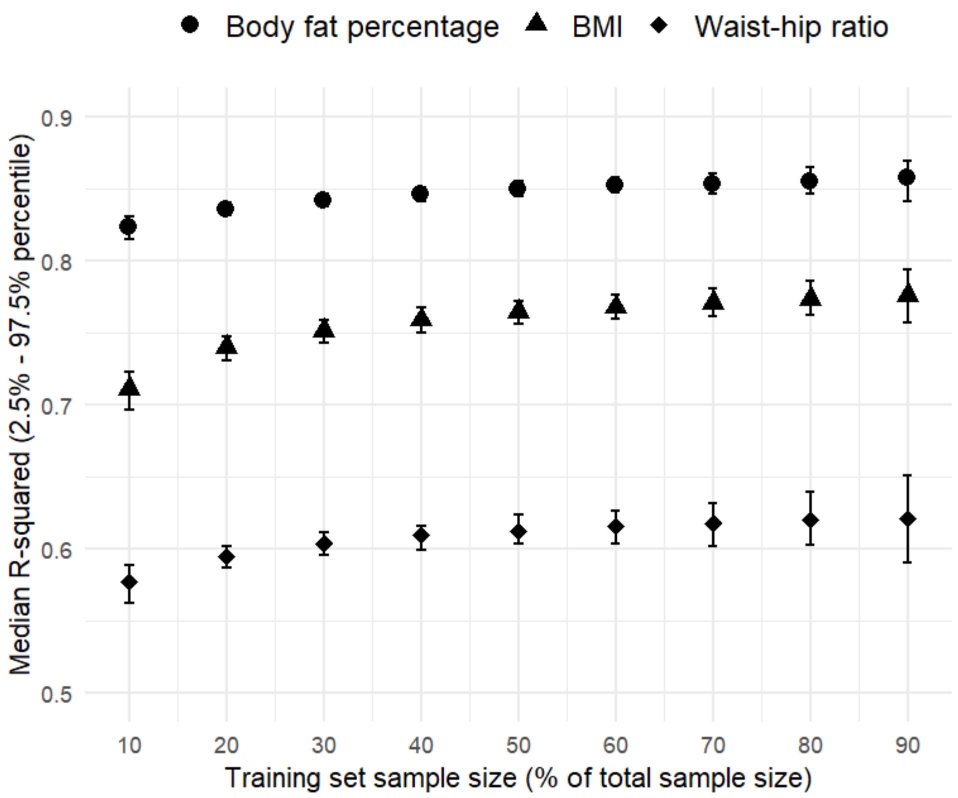

**Supplemental Figure 3. The LASSO Selected Proteins Shared across Obesity-related Phenotypes.**

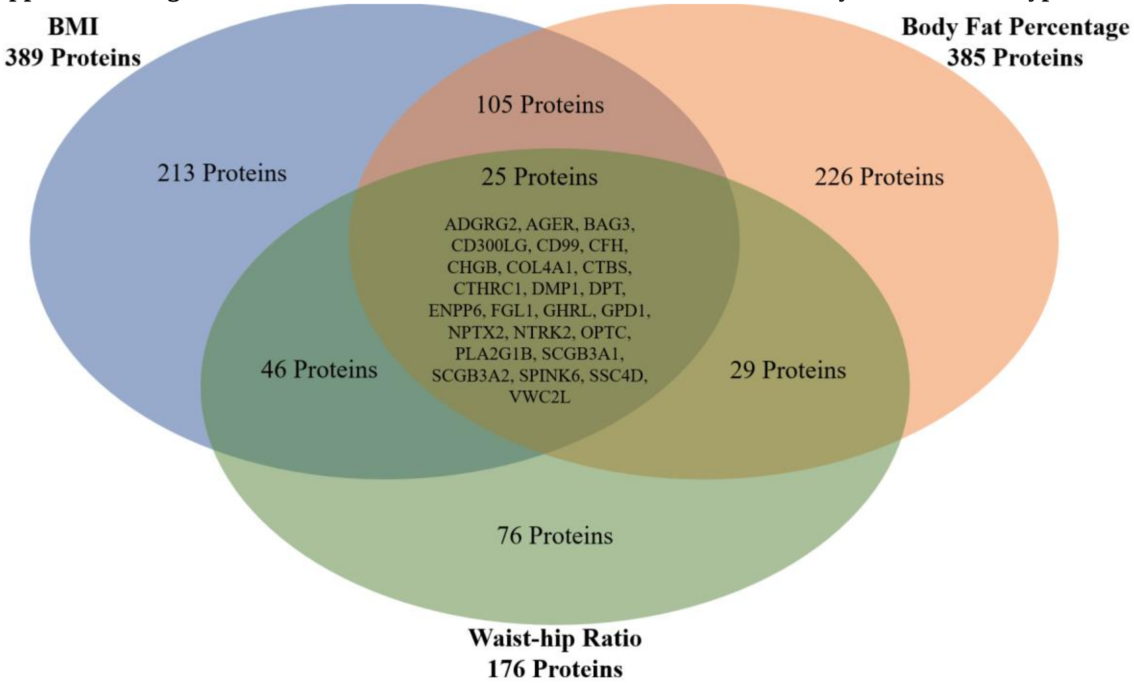

**Supplemental Figure 4. Linear Associations Between Predicted Protein Scores of Obesity-related Phenotypes and Measured Phenotypes.**

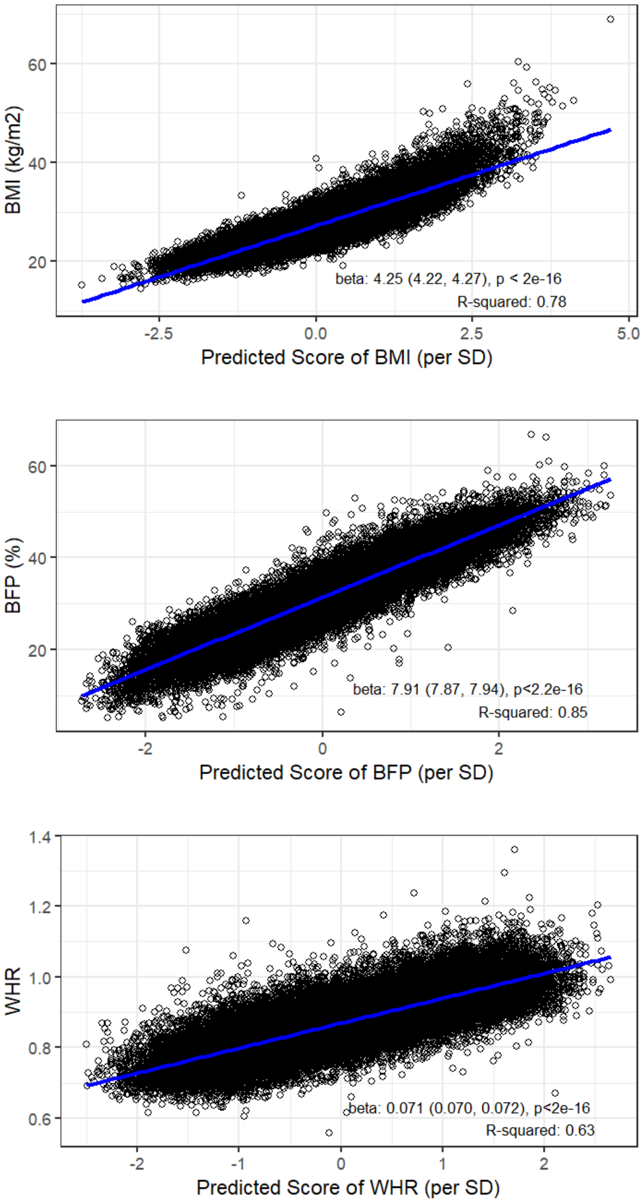

**Supplemental Figure 5. Forest Plot of the Associations Between Protein Predicted Scores of Obesity-related Phenotypes and MACE Individual Components. Model 1: adjusted for age, sex and race (white vs. other); Model 2: adjusted for the measured obesity-related phenotype (BMI, body fat percentage, or waist-hip ratio) in addition to Model 1; Model 3: adjusted for total cholesterol, high density lipoprotein cholesterol, systolic blood pressure, estimated glomerular filtration rate calculated using the 2021 CKD-EPI equation, diabetes, current smoking, blood pressure lowering medication use, cholesterol lowering medication use in addition to Model 2.**

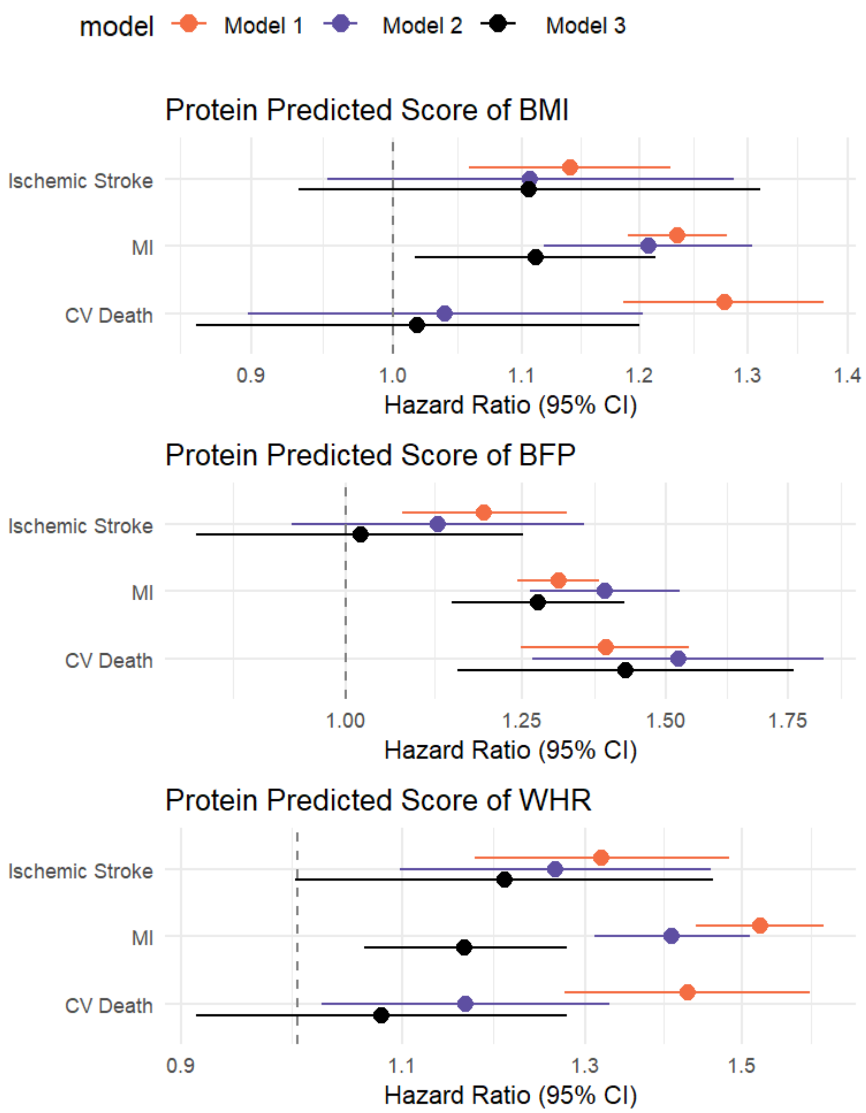
